# The Long-term Mediation Role of Cytokines on the Causal Pathway from Maternal Gestational Age to Offspring Visual System: Lifecourse-Network Mendelian Randomization

**DOI:** 10.1101/2022.09.30.22280560

**Authors:** Lei Hou, Yunxia Li, Lili Kang, Xiaoying Li, Hongkai Li, Fuzhong Xue

## Abstract

**Background:** Gestational duration has a non-negligible impact on eye diseases. However, the long-term role of cytokines on the causal relationship of maternal gestational age on offspring visual impairment remains unclear.

**Methods:** We perform a lifecourse-network Mendelian randomization (MR) to explore the causal relationships among maternal gestational duration (from EGG and iPSYCH, N=84,689), neonatal/adult cytokines (from the NHGRI-EBI Catalog, N=764/4,618) and adult eye diseases (from FinnGen consotium, N=309,154) using summary-level data from large genome-wide association studies. Multiplicative random effects inverse variance weighted (IVW) and multivariable-IVW method are the main analysis methods and the other 15 pleiotropy-robust methods, weak IV-robust methods and outliers-robust methods are performed as auxiliary methods.

**Results:** We find that maternal gestational age (early preterm birth, preterm birth, gestational duration and postterm birth) has causal relationships with 42 eye diseases. Specially, four neonatal cytokines: TNF-α, IL10, GROA and CTACK, as well as four adult cytokines: CTACK, IL10, IL12p70 and IL6.26 are mediators in the causal relationships between early preterm birth and preterm birth to 8 eye diseases. However, after adjusting for these mediators, null direct causal effect of early preterm birth and preterm birth on 8 eye diseases can be found. In addition, there is no mediator in the causal relationships from gestational duration and postterm birth to eye diseases.

**Conclusion:** The influences of maternal gestational duration on the offspring eye diseases through cytokines are long-term and lifecourse.

## 1 Introduction

For the last 30 years, at least 2.2 billion people have near or distance vision impairment all of the world. Almost half of these cases include those with moderate or severe distance vision impairment or blindness due to unaddressed refractive error, cataract, glaucoma, corneal opacities, diabetic retinopathy, and trachoma, as well as near vision impairment caused by unaddressed presbyopia [1]. Vision impairment severely affects the quality of life among adult populations, who tend to have lower rates of labor force participation and productivity and higher rates of depression and anxiety. In the case of older adults, vision impairment can lead to social isolation, difficulty walking, a higher risk of falls and fractures, and a greater likelihood of early entry into nursing or care homes [2]. A Study for Swedish Medical Birth Registry revealed an increase in the risk of visual impairment at whether small-for-gestational-age or large-for-gestational-age [3]. A previous literature demonstrated that children who were born very preterm (28–32 weeks) were at a significantly higher risk for abnormal visual and neurological development, when compared to children born at full term [4]. These abnormalities include retinopathy of prematurity (ROP), strabismus, color vision deficits, visual field defects, decreased visual acuity and refractive error [5]. In a retrospective cohort study from the Soroka Univer-sity Medical Center (SUMC), rate of visual disturbances and retinopathy of prematurity (ROP) was significantly higher among the 24–28 gestational weeks group [6]. Kozeis et al. [7] found preterm infants were associated with impairment of some aspects of visual function.

A study with a large prospective cohort in Indonesia from October 2007 till October 2009 found the associations between levels of 25 cytokines in newborn infants and gestational duration [8]. A prospective cohort study revealed that there was a relationship between high plasma levels of interleukin-6 (IL-6), interleukin-8 (IL-8), and Tumor Necrosis Factor-α (TNF-α) in the first day of life with the later development of ROP severe enough to treat in preterm infants with early-onset sepsis [9]. Masahiko et al. [10] investigated the causal relationships between vitreous levels of cytokines, including IL-6 and vascular endothelial growth factor (VEGF), and visual prognosis after pars plana vitrectomy (PPV) with arteriovenous sheathotomy in patients with branch retinal vein occlusion (BRVO) associated with macular oedema. Various cytokines involved in inflammation and angiogenesis including elevated interleukin-31(IL-31), LIF, SDF1-α, VEGF-A, VEGF-D might be involved in the pathogenesis of neovascular age-related macular degeneration (nAMD) and polypoidal choroidal vasculopathy (PCV) [11]. The role of cytokines on the causal relationship of maternal gestational age on offspring visual impairment is unclear.

Mendelian randomization (MR) analysis [12] is the exploitation of a natural experiment to obtain a causal association of an exposure on an outcome using genetic variants as instrumental variants (IVs). A valid instrumental variable must satisfy the following three assumptions: (1) Relevance – IV (G) is robustly associated with the exposure (X); (2) Exchangeability – IV (G) is not associated with any confounder (U) of the exposure–outcome relationship; (3) Exclusion restriction – IV (G) is independent of the outcome (Y) conditional on the exposure (X) and all confounders of the exposure-outcome relationship (i.e. the only path between the instrument and the outcome is via the exposure). Network MR can be used to estimate direct and indirect causal effects in a mediation setting [13]. Direct effect can be estimated using multivariable MR (MVMR) [14] by regressing genetic associations of outcome on the exposure and the mediator. Indirect effect can be estimated using two ways: difference method, that is, total effect of X on Y minus the direct effect; product method, the production of causal effects of X on the mediator and the mediator on outcome. Lifecourse MR is designed to evaluate the early life and adult causal relationship between two variables [15–16].

In this article, we aim to evaluate the long-term role of neonatal and adult cytokine levels on the causal pathway from maternal gestational age to offspring visual impairment using network MR and lifecourse MR.

## 2 Materials and methods

Study overview is illustrated in **Figure 1**. Firstly, UVMR is performed to calculate the total effects of gestational duration on eye diseases. Then a lifecourse MR approach is used to explore the long-term causal effects of gestational duration on cytokines using UVMR and MVMR. This process aims to disclose whether the effects of gestational duration on adult cytokines through neonatal cytokines or not. Similarly, a lifecourse MR approach is also used to explore whether the effects of neonatal cytokines on eye diseases through adult cytokines or not. Finally, we use MVMR to calculate the direct effects of gestational duration on eye diseases after adjusting for neonatal/adult cytokine levels.

**Figure 1.**
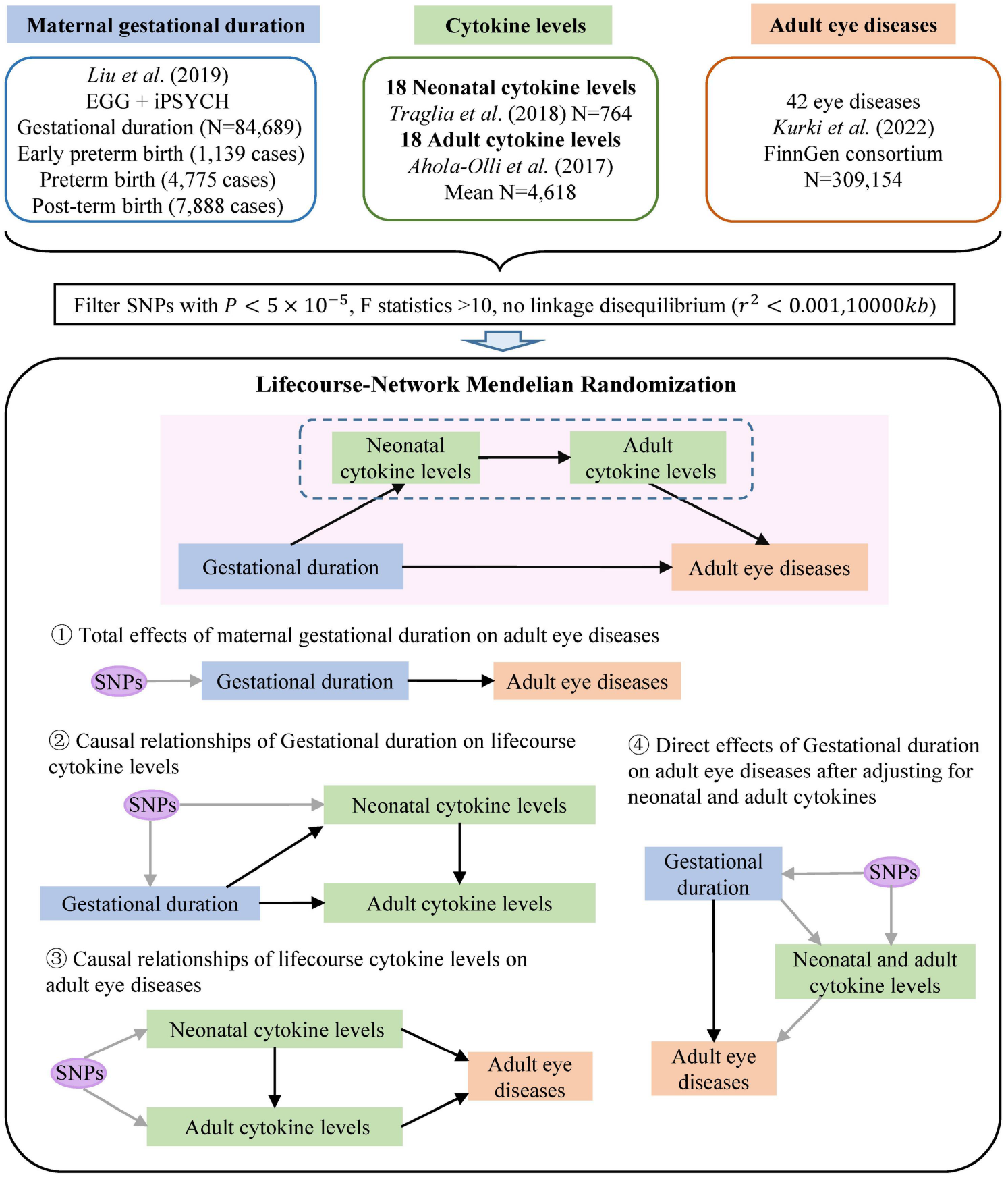
Study overview.

### 2.1 Data resources

GWAS summary statistics for maternal gestational duration are from fetal genome-wide meta-analyses of gestational duration (84,689 European), early preterm (<34 completed weeks of gestation, 1,139 cases), preterm (<37 weeks, 4,775 cases), and postterm birth (≥42 weeks, 7,888 cases), combining data from the Early Growth Genetics Consortium (EGG) consortium and the Lundbeck Foundation Initiative for Integrative Psychiatric Research (iPSYCH) study [17]. Gestational duration in days was regressed on infant sex and the resulting residuals were quantile transformed to a standard normal distribution before being tested for association with fetal single nucleotide polymorphism (SNP) genotypes using linear regression. The dichotomous outcomes early preterm birth, preterm birth, and postterm birth were analyzed using logistic regression. Details of four gestational duration variables are shown in the Supplementary Table S1.

GWAS summary statistics for neonatal cytokine levels are from a genetic study of 790 mother-infant pairs in a population-based nested case-control study (the Early Markers for Autism (EMA) study). This study analyzes 42 neonatal bloodspot-derived immune mediators (cytokines/chemokines) in the context of maternal and fetal genotype. Neonatal levels of peripheral blood immune markers were determined using a commercially available, slightly modified, Luminex multiplex magnetic bead assay [18]. GWAS summary statistics for adult cytokine levels are from a GWAS meta-analysis with up to 8,293 Finns in the combination of a multicenter follow-up study Young Finns Study (YFS) and a population-based cross-sectional study (FINRISK2002). In these two studies, a total of 48 cytokines were measured by using Bio-Rad’s premixed Bio-Plex Pro Human Cytokine 27-plex Assay and 21-plex Assay, and Bio-Plex 200 reader with Bio-Plex 6.0 software [19]. Finally, 18 duplicated cytokines in neonatal and adult cytokine GWAS datasets are included in our study. Details of 18 cytokine levels are shown in the Supplementary Table S2.

GWAS summary statistics for adult eye diseases are from FinnGen consortium [20]. FinnGen aims to collect and analyze genome and health data from 500,000 Finnish biobank participants. We are interested in the eye diseases whose IDs start with ‘H7’, including 163 eye-related diseases. We use the latest data release in 2022, and total sample size is 309,154, including 173,746 females and 135,408 males. We firstly filter the eye diseases which are affected by maternal gestational duration variables then 42 diseases remain. Details of 42 eye diseases are shown in the Supplementary Table S3.

### 2.2 Instrumental variants selection and harmonization

For four gestational duration variables, 18 neonatal and adult cytokine levels, we use the same filter criterion to select instrumental variants for UVMR and MVMR, respectively. For UVMR, we filter the SNPs with P<5×10^−5^ and identify independent SNPs based on a linkage disequilibrium (LD) cutoff of *r*^2^<0.001 (10000*kb*) as instrumental variants. For MVMR, we repeat LD clumping but use aggregated sets of genetic variants for all our exposures to ensure they were also independent. The F statistics and conditional F statistics [21] of all instrumental variants are above 10, which ensure the instrumental strength. Finally, we extract the GWAS summary statistics of these instrumental variants from the outcome. Genetic estimates for our exposures were harmonized with disease outcomes using the ‘TwoSampleMR’ R package.

### 2.3 Statistical analysis

#### UVMR

We apply the multiplicative random effects inverse variance weighted (IVW) method [22] for initial analyses, which takes the SNP-outcome estimates and regresses them on those for the SNP-exposure associations. 12 subsequently sensitivity analyses include fixed effects IVW [22], pleiotropy-robust methods (MR-Egger [23], simple median, weighted median, penalized weighted median [24], simple mode, weighted mode, simple mode (NOME), weighted mode (NOME) [25]), weak IV-robust method (MR-RAPS [26]), outliers-robust methods (MR Radial [27], MR PRESSO [28]). Egger’s test [23] is used for pleiotropy test. Heterogeneity tests [28] include two methods: IVW and MR-Egger.

#### MVMR

We choose MV-IVW [29] as the main method for MVMR, which takes the SNP-outcome estimates and regresses them on those for multiple SNP-exposure associations. Additional three methods: MV-Egger, MV-lasso and MV-median, also performed to evaluate the robustness of IVW estimates to horizontal pleiotropy. A modified form of Cochran’s Q statistic [30] is calculated to measure the pleiotropy.

#### Network MR

We use the network MR design [13] to explore the mediation roles of cytokine levels on the causal pathways from gestational duration to eye diseases. Firstly, UVMR is performed to calculate the total effects of gestational duration on eye diseases. Secondly, we use a lifecourse MR approach to calculate the causal effects of gestational duration on cytokine levels and the causal effects of cytokine levels on eye diseases. Thirdly, MVMR is performed to calculate the direct effects of gestational duration on eye diseases not through cytokines.

#### Lifecourse MR

In lifecourse MR, we are interested in the neonatal and adult cytokine levels. On the one hand, for the causal effects of gestational duration on cytokine levels, we firstly calculate the causal effects of gestational duration on neonatal and adult cytokine levels using UVMR, then MVMR is performed to calculate the direct effects of gestational duration on adult cytokines adjusting for neonatal cytokine levels, which are affected by gestational duration. On the another hand, for the causal effects of cytokine levels on eye diseases, we firstly calculate the causal effects of neonatal and adult cytokine levels on eye diseases using UVMR, then MVMR is performed to calculate the direct effects of neonatal cytokines on eye diseases adjusting for adult cytokine levels, which affect eye diseases.

Statistical analyses were performed using R version 3.6.1 with the R packages ‘TwosampleMR’, ‘MendelianRandomization’ and ‘MVMR’.

## 3 Results

### 3.1 Total effects of maternal gestational duration on offspring eye diseases

Firstly, we examine the causal effects of four gestational duration variables on the 163 eye diseases using UVMR. Results are shown in the Figure 2 (A). We find that four gestational duration variables have causal effects on 42 eye diseases. Both early preterm birth and preterm birth are risk factors of disorders of sclera, cornea, iris and ciliary body.

**Figure 2.**
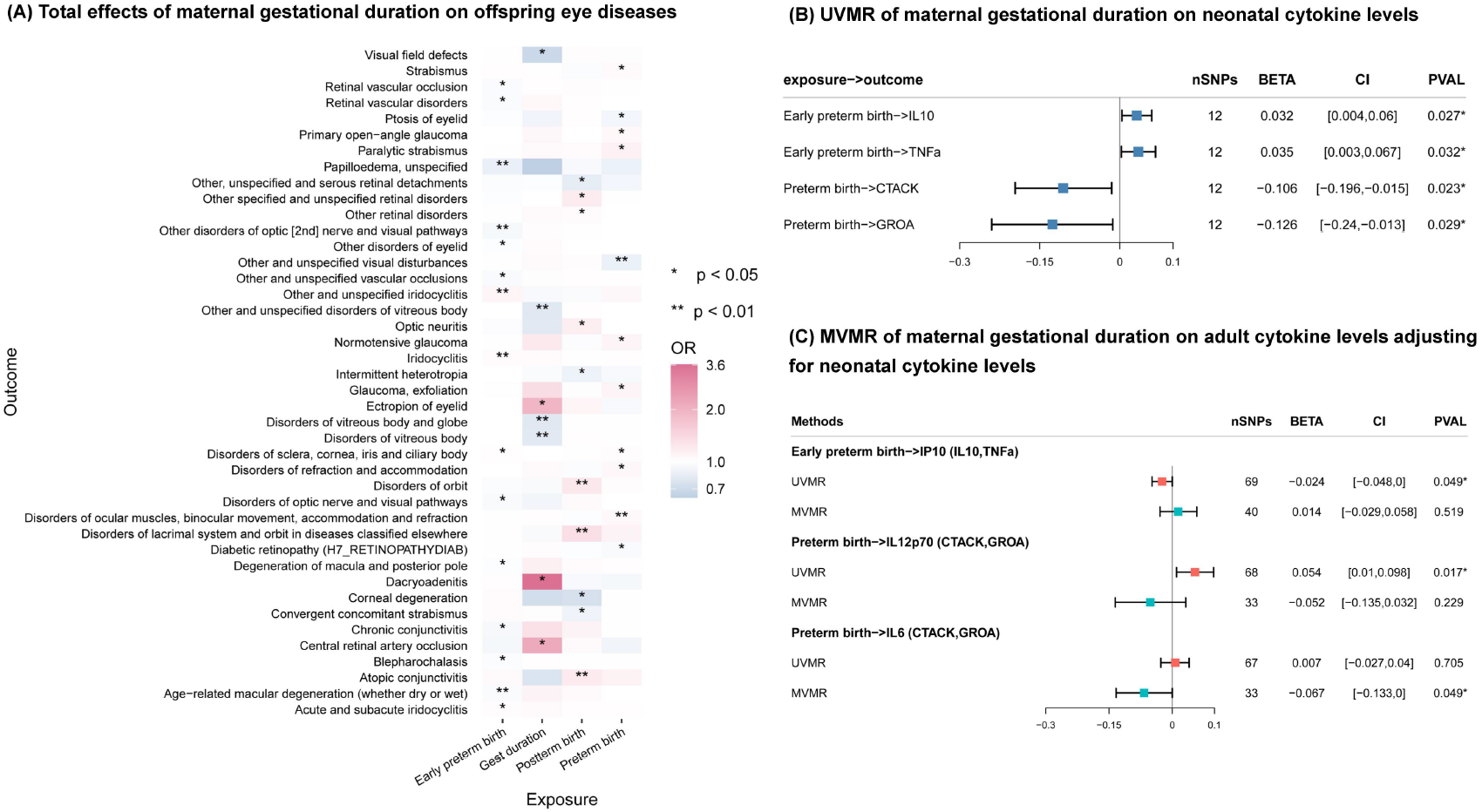
(A) UVMR results of maternal gestational duration on adult eye diseases; (B) UVMR results of maternal gestational duration on neonatal cytokine levels; (C) MVMR results of maternal gestational duration on adult cytokine levels adjusting for neonatal levels.

77 genetic variants determined early preterm birth causally affects the risk of 15 eye diseases. It increases the risk of iridocyclitis, acute and subacute iridocyclitis, but has protective effects on vascular disorders (e.g. retinal vascular occlusion, retinal vascular disorders), papilloedema, disorder of optic nerve and visual pathways, degeneration of macula and posterior pole, chronic conjunctivitis, blepharochalasis and age-related macular degeneration (whether dry or wet).

We find preterm birth has causal effects on 11 eye diseases using 73 instrumental variants. Preterm birth decreases the risk of ptosis of eyelid and diabetic retinopathy. On the contrary, preterm birth is a risk factor of strabismus, glaucoma, disorder of refraction and accommodation, disorder of ocular muscles, binocular movement, accommodation and refraction.

For postterm birth, we use 68 SNPs as instrumental variants and MR results reveal that postterm birth is causally associated with 10 eye-related diseases. It is a protective factor of intermittent heterotropia, corneal degeneration and convergent concomitant strabismus, but can increase the risk of optic neuritis, disorder of orbit, atopic conjunctivitis.

103 SNPs determined gestational duration has positive effects on ectropion of eyelid, dacryoadenitis and central retinal artery occlusion. On the contrary, it is a protective factor of visual field defects, disorder of vitreous body and globe, disorder of vitreous body.

Results of sensitivity analysis are shown in Supplementary Fig S1.1-S1.4. Results of pleiotropy test and heterogeneity tests are shown in Supplementary Table S4.1-S5.4. Results reveal that there is no pleiotropy in MR analysis, but there is heterogeneity exists in the Wald Ratio of 73 SNPs in the causal relationship of preterm birth and paralytic strabismus (Supplementary Table S5.2). After remove outliers, outliers-robust methods (MR Radial and MR PRESSO) also valid the existence of this causal relationship.

### 3.2 Lifecourse MR of maternal gestational duration on cytokine levels

Firstly, we perform UVMR of maternal gestational duration on neonatal cytokine levels (Figure 2 (B)). 12 SNPs are selected as IVs for all four gestational duration variables. We find that early preterm birth increases the levels of IL10 and TNF-α. On the contrary, preterm birth has negative causal relationships with CTACK and GROA levels. Besides, the other two gestational duration variables (postterm birth and gestational duration) have no causal effect on neonatal cytokine levels. Results of sensitivity analysis show that there is no pleiotropy and heterogeneity. Details are shown in Supplementary Fig S2 and Table S6-S7.

Then we perform UVMR and MVMR to explore the causal relationships from maternal gestational duration on adult cytokine levels (Figure 2 (C)). Results of UVMR reveal that 69 IVs determined early preterm birth decreases the levels of IP10, and 68 IVs determined preterm birth increase the levels of IL12p70. Postterm birth and gest duration also have no causal effect on adult cytokine levels. Results of sensitivity analysis show that there is no pleiotropy and heterogeneity. Details are shown in Supplementary Fig S3 and Table S8-S9. For MVMR, after adjusting for neonatal IL10 and TNF-α, early preterm birth has no direct effect on IP10. In addition, preterm birth also has no direct effect on IL12p70 after adjusting for neonatal CTACK and GROA, but a new causal relationship emerges, that is, preterm birth directly decreases the level of IL6 after adjusting for neonatal CTACK and GROA. Results of sensitivity analysis show that there is no pleiotropy and MVMR lasso provides the similar results. Details are shown in Supplementary Fig S4 and Table S10.

### 3.3 Lifecourse MR of cytokine levels on offspring eye diseases

In this section, we explore the above seven significant cytokine levels (IL10, TNF-α, CTACK, GROA, IP10, IL12p70 and IL6) on 42 eye diseases using lifecourse MR approach.

Firstly, UVMR of 7 adult cytokine levels on 42 eye diseases are performed and results find that 6 adult cytokine levels are causally associated with 12 eye-related diseases (Figure 3). IL6 and GROA negatively affects intermittent heterotropia, but TNF-α positively affects intermittent heterotropia. High level of CTACK decreases the risks of paralytic strabismus. GROA has positive causal relationships with dacryoadenitis. IL10 positively affects normotensive glaucoma, disorder of vitreous body and globe, vitreous body, optic nerve and visual pathways. High level of IL12p70 causally decreases the risk of corneal degeneration. IL6 has negatively association with disorders of orbit. TNF-α increases the risk of convergent concomitant strabismus. Results of sensitivity analysis show that there is no pleiotropy, but there exist heterogeneity in the causal relationships between IL10 and normotensive glaucoma, disorder of optic nerve and visual pathways. For the former, outliers-robust methods show that there is no causal relationship but other pleiotropy-robust methods provide the significant causal relationship. For the latter, outliers-robust methods still show the similar results after remove outliers. Details are shown in Supplementary Fig S5.1-S5.2 and Table S11-S12

**Figure 3.**
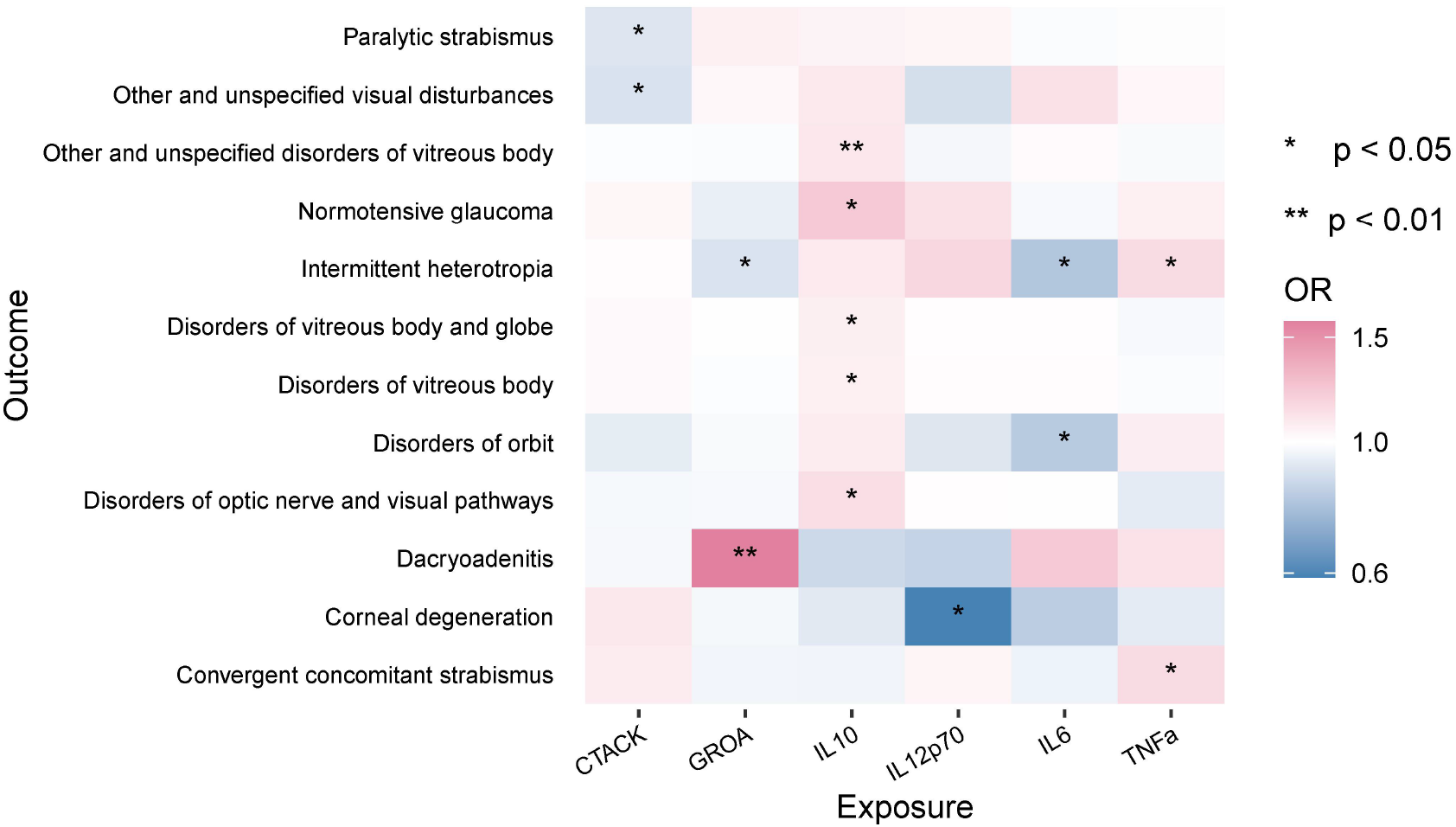
UVMR results of adult cytokine levels on adult eye diseases.

Then we perform UVMR and MVMR to explore the causal relationships from neonatal cytokine levels on 12 eye diseases (Figure 4). Results of UVMR show that seven neonatal cytokines are causally associated with six eye diseases. High level of GROA increases the risk of disorder of vitreous body and globe. IP10 positively affects corneal degeneration, TNF-α has positive causal relationships with normotensive glaucoma and paralytic strabismus. Results of sensitivity analysis show that there is no pleiotropy and heterogeneity. Details are shown in Supplementary Fig S6.1-S6.2 and Table S13-S14.2. For the results of MVMR, after adjusting corresponding adult cytokines, the causal relationships in UVMR disappear. Besides, several new direct causal relationships are found. For example, after adjusting for adult IL10, high level of CTACK decreases the risk of disorder of vitreous body and globe; GROA negatively affect normotensive glaucoma after; neonatal IL10 also has direct causal effect on normotensive glaucoma; IL12p70 positively affects disorder of vitreous body; high level of IL6 decreases the risk of normotensive glaucoma. Results of sensitivity analysis show that there is no pleiotropy and other MVMR methods provide the similar results. Details are shown in Supplementary Fig S7 and Table S15.

**Figure 4.**
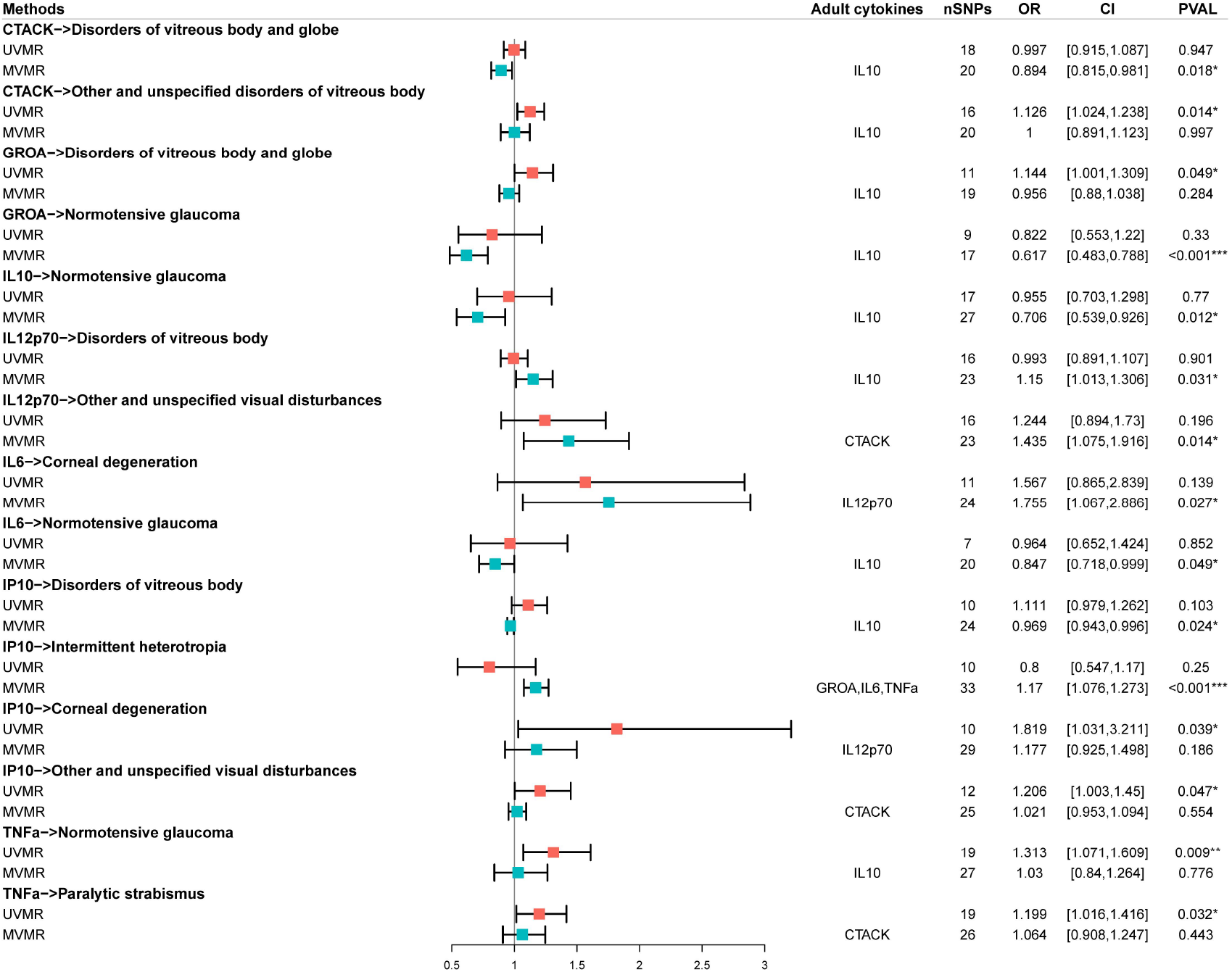
MVMR results of neonatal cytokine levels on eye diseases adjusting for adult cytokine levels. The column named ‘adult cytokines’ denotes the adjusting variables in MVMR.

### 3.4 Mediation pathways from maternal gestational duration to eye diseases through neonatal/adult cytokines

The causal pathways from maternal gestational duration to eye diseases through neonatal/adult cytokines are shown in Figure 5. There are 10 mediation pathways from early preterm birth on four eye diseases through two neonatal cytokines: TNF-α and IL10, and two adult cytokines: CTACK and IL10. Early preterm birth decreases the risk of paralytic strabismus through increasing the levels of neonatal TNF-α and adult CTACK. In addition, early preterm birth causally increases the risk of disorder of vitreous body, optic nerve and visual pathways as well as normotensive glaucoma by increasing neonatal TNF-α and IL10 levels, and adult IL10 cytokines. There is one pathway that early preterm birth has a positive effect on neonatal IL10 level, further decreases the risk of normotensive glaucoma not through the adult cytokines.

**Figure 5.**
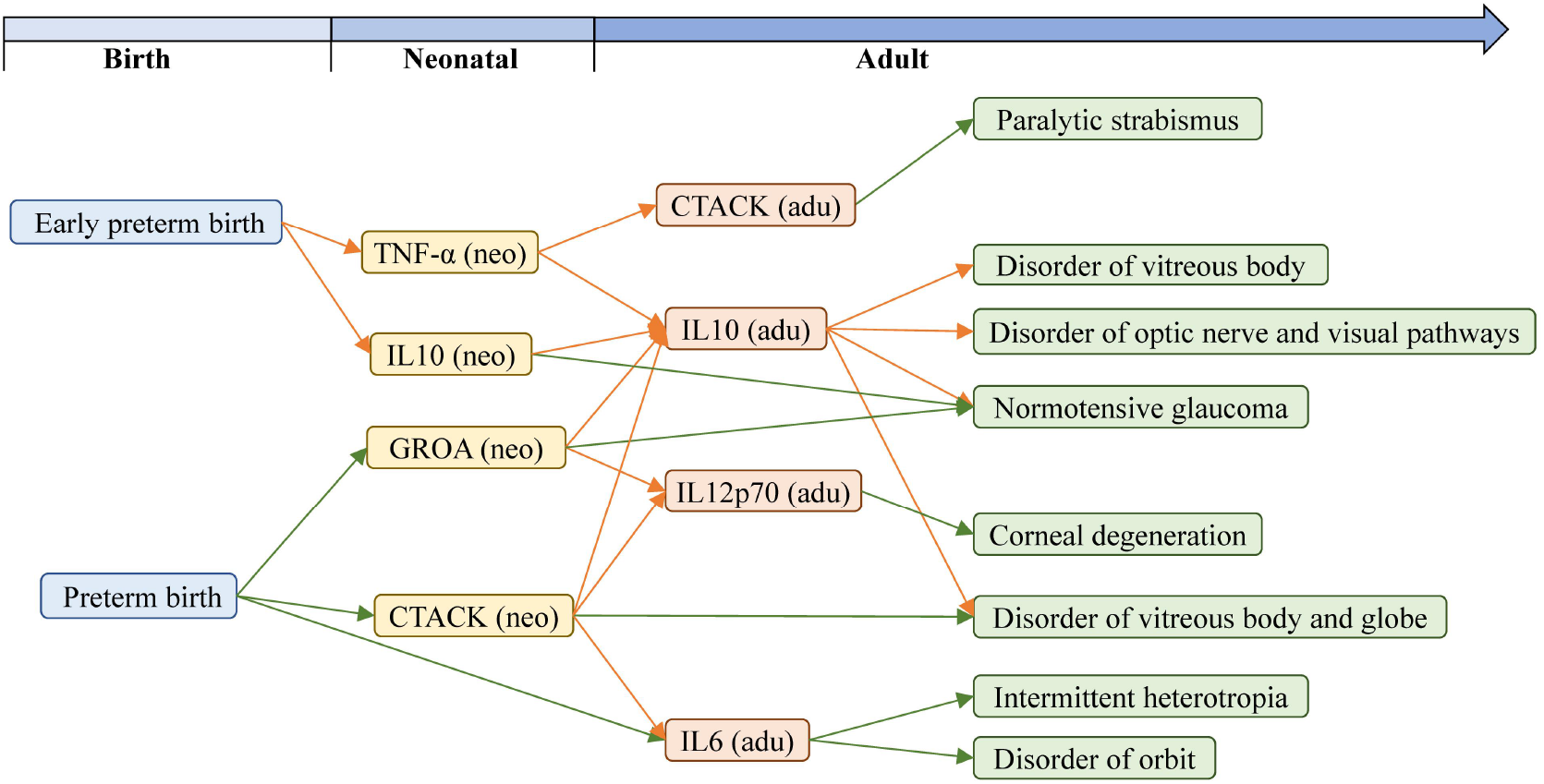
Causal mediation pathways from maternal gestational duration to adult eye diseases through neonatal/adult cytokine levels. The orange edges denote the positive causal effects. The green edges denote the negative causal effects. The blue boxes denote maternal gestational duration. The yellow boxes denote neonatal cytokines. The orange boxes denote adult cytokines. The green boxes denote eye diseases.

There are 16 mediation pathways from preterm birth on seven eye diseases through two neonatal cytokines: CTACK and GROA, and three adult cytokines: IL12p70, IL10 and IL6. Preterm birth has a negative causal relationship with the levels of neonatal GROA and CTACK, the increase the levels of adult IL10 and IL12p70. Further, adult IL12p70 decreases the risk of corneal degeneration, and adult IL10 increases the risk of disorder of vitreous body, globe, optic nerve and visual pathways as well as normotensive glaucoma. Preterm birth has a protective effect on the intermittent heterotropia through decreasing level of neonatal CTACK, with increasing level of adult IL6. Preterm birth also directly decreases the level of adult IL6. No mediation pathways from gestational duration and postterm birth to eye diseases is found through network and lifecourse MR analysis.

### 3.5 Direct effects maternal gestational duration on offspring eye diseases not through cytokines

MVMR is performed to calculate the direct effects of early preterm birth and preterm birth on above 12 eye diseases after adjusting for the neonatal and adult cytokines which are causally associated with exposure and outcome simultaneously, that is, adjusting for the mediators from gestational duration to eye diseases. The results of MVMR (Figure 6) show that nearly no direct causal relationship between early preterm birth/preterm birth and eye diseases after adjusting for neonatal and adult/neonatal cytokines can be found except one: MVMR lasso reveals that early preterm birth directly increases the risk of disorder of vitreous body and globe not through neonatal TNF-α and IL10 as well as adult IL10. Results of sensitivity analysis show that there is weak pleiotropy, but pleiotropy-robust methods also support the null direct effect. Details are shown in Supplementary Table S16.

**Figure 6.**
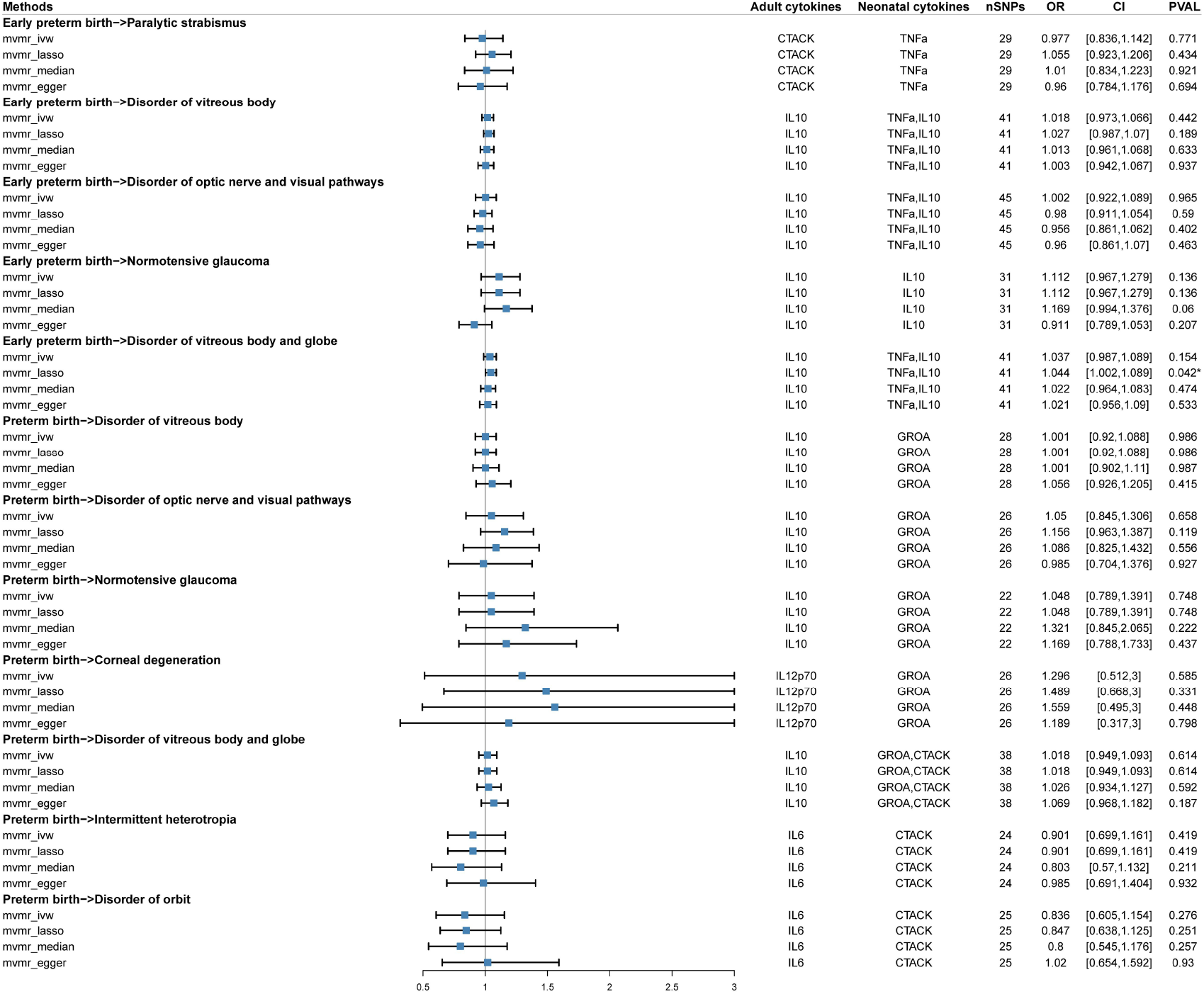
MVMR results of early preterm birth and preterm birth on eye diseases after adjusting for neonatal/adult cytokine levels. The columns named ‘adult cytokines’ and ‘neonatal cytokines’ denote the adjusting variables in MVMR.

## 4 Discussion

In this study, we conduct a network MR, embedding lifecourse MR approach, to explore the role of neonatal/adult cytokines in the causal pathway from maternal gestational duration on eye diseases. We find that maternal gestational duration has causal relationship with 42 eye diseases, and eight of these relationships are mediated by neonatal cytokines, three of these relationships are mediated by adult cytokines. Totally, 26 mediation pathways are founded for early preterm birth and preterm birth, but no mediation pathways from gestational duration and postterm birth to eye diseases is found.

For the mediation pathways, there are several literatures support these results. Serum IL-10 levels are increased in preterm infants and an inverse correlation between IL-10 cord blood levels and gestational age has been reported [31]. Increased levels of cytokines (IL-1β, IL-2, IL-5, IL-6, IL-8, IL-10, IL-13, IL-15, IL-17, MCP-1/CCL2, TNF-α) have been reported in tears of glaucoma patients on medical therapy [32–33]. Retinopathy of prematurity is an eye disease that affects many premature babies [34]. Our results provide a possible interpretable pathway that preterm birth affect the risk of retinopathy through neonatal CTACK level. TLR2 and TLR4-mediated TNF-α, IL-1β, IL-6, IL-12p70, and IL-10 production in vitro increased over the first month of life in preterm infants [35]. An early marker of the activated inflammatory cascade is IL6 levels in serum peak 3–4 hours after endotoxin stimulation and may be used as an early diagnostic marker of neonatal bacterial infection [36]. IL-6 plays an important role in the pathogenesis of Graves’ disease and its orbital component [37]. The levels of IL-12p70 is significantly elevated in bullous keratopathy eyes, compared with healthy control [38].

The causal direction is naturally determined by lifecourse, that is a person from birth to adulthood, thus there is no influence of bidirectional causal relationship. This is the advantage of our study. In addition, lifecourse MR reveals the change of neonatal effect and adult effect, provide the lifecourse causal association between cytokine and maternal gestational duration and eye diseases. Network MR dissects the direct and indirect effect from maternal gestational duration to eye diseases, and clarify the mediation role of neonatal/adult cytokines. Pleiotropy-robust methods, weak IV-robust method and outliers-robust methods jointly validate the causal relationships and provide strong evidences for this study. On of drawback in this study is that the dataset of neonatal cytokines contains a relatively small number of SNPs, thus the corresponding MR results have less power. Further study with huge datasets is needed to verify these results.

In conclusion, this study provides the lifecourse causal relationship from maternal gestational age to neonatal/adult cytokines then to adult eye diseases. The influences of maternal gestational duration on the offspring eye diseases are long term and lifecourse, and biological and pathological mechanisms need further studies.

## Data Availability

All data produced in the present study are available upon reasonable request to the authors

## Acknowledgement

We want to acknowledge the participants and investigators of the FinnGen study.

## Data availability statement

All the GWAS summary data are publically available. GWAS summary data for maternal gestational duration in EGG and iPSYCH can be download at http://egg-consortium.org/gestational-duration-2019.html. GWAS summary data for 18 neonatal/adult cytokines in the NHGRI-EBI Catalog can be download at https://www.ebi.ac.uk/gwas/summary-statistics. GWAS summary data for adult eye diseases in the FinnGen consotium can be download at https://www.finngen.fi/en/access_results.

## Ethics approval and patient consent statement

Ethical approval was not sought, because this study involved analysis of publicly available summary-level data from GWASs, and no individual-level data were used.

## Author contributions

HL, FX and XL conceived the study. YL contributed to data collection. LH contributed to the data analysis and sensitivity analyses. LH, YL and LK contributed to the application and wrote the manuscript. HL, FX and XL modified the manuscript. All authors reviewed and approved the final manuscript.

