## Supplementary material for "The Long-term Mediation Role of Cytokines on the Causal Pathway from Maternal Gestational Age to Offspring Visual System: Lifecourse-Network Mendelian Randomization": Supplomentary Notes: Supplementary Notes.pdf

### 1 Datasets

**Table S1. Datasets information of 4 pregnancy disorders**

| trait | year | Consortium | Author | Pmid | nsnp | build | category | ncase | N |
|---|---|---|---|---|---|---|---|---|---|
| Gestational duration | 2019 | EGG + iPSYCH | Xueping Liu | 31477735 | 7646297 | HG19/GRCh37 | Continious | - | 84689 |
| Early preterm birth (<34 weeks) | 2019 | EGG + iPSYCH | Xueping Liu | 31477735 | 7588467 | HG19/GRCh37 | Binary | 1139 | 84689 |
| Preterm birth (<37 weeks) | 2019 | EGG + iPSYCH | Xueping Liu | 31477735 | 7545601 | HG19/GRCh37 | Binary | 4775 | 84689 |
| Post-term birth (⩾42 weeks) | 2019 | EGG + iPSYCH | Xueping Liu | 31477735 | 7583965 | HG19/GRCh37 | Binary | 7888 | 84689 |

**Table S2. Datasets information of 18 adult and neonatal cytokine levels**

| Trait name | | Adult | | | | | Neonatal | | | | | Build |
|---|---|---|---|---|---|---|---|---|---|---|---|---|
| | | year | Author | Pmid | N | nsnp | year | author | Pmid | N | nsnp | |
| CTACK | CTACK levels | 2016 | Ahola-Olli AV | 27989323 | 3631 | 9568408 | 2018 | Michela Traglia | 30134952 | 764 | 652831 | HG19/GRCh37 |
| GROA | Growth-regulated protein alpha levels | 2016 | Ahola-Olli AV | 27989323 | 3505 | 9528505 | 2018 | Michela Traglia | 30134952 | 764 | 652831 | HG19/GRCh37 |
| IFNg | Interferon gamma levels | 2016 | Ahola-Olli AV | 27989323 | 7701 | 9785363 | 2018 | Michela Traglia | 30134952 | 764 | 652831 | HG19/GRCh37 |
| IL10 | Interferon gamma-induced protein 10 levels | 2016 | Ahola-Olli AV | 27989323 | 3685 | 9576881 | 2018 | Michela Traglia | 30134952 | 764 | 652831 | HG19/GRCh37 |
| IL12p70 | Interleukin-1-beta levels | 2016 | Ahola-Olli AV | 27989323 | 3309 | 9983642 | 2018 | Michela Traglia | 30134952 | 764 | 652831 | HG19/GRCh37 |
| IL13 | Interleukin-10 levels | 2016 | Ahola-Olli AV | 27989323 | 7681 | 9793415 | 2018 | Michela Traglia | 30134952 | 764 | 652831 | HG19/GRCh37 |
| IL16 | Interleukin-12p70 levels | 2016 | Ahola-Olli AV | 27989323 | 8270 | 9799886 | 2018 | Michela Traglia | 30134952 | 764 | 652831 | HG19/GRCh37 |
| IL1b | Interleukin-13 levels | 2016 | Ahola-Olli AV | 27989323 | 3557 | 9539073 | 2018 | Michela Traglia | 30134952 | 764 | 652831 | HG19/GRCh37 |
| IL2 | Interleukin-16 levels | 2016 | Ahola-Olli AV | 27989323 | 3483 | 9551485 | 2018 | Michela Traglia | 30134952 | 764 | 652831 | HG19/GRCh37 |
| IL4 | Interleukin-2 levels | 2016 | Ahola-Olli AV | 27989323 | 3475 | 9512914 | 2018 | Michela Traglia | 30134952 | 764 | 652831 | HG19/GRCh37 |
| IL6 | Interleukin-4 levels | 2016 | Ahola-Olli AV | 27989323 | 8124 | 9786064 | 2018 | Michela Traglia | 30134952 | 764 | 652831 | HG19/GRCh37 |
| IL8 | Interleukin-6 levels | 2016 | Ahola-Olli AV | 27989323 | 8189 | 9790590 | 2018 | Michela Traglia | 30134952 | 764 | 652831 | HG19/GRCh37 |
| IP10 | Interleukin-8 levels | 2016 | Ahola-Olli AV | 27989323 | 3526 | 9517348 | 2018 | Michela Traglia | 30134952 | 764 | 652831 | HG19/GRCh37 |
| MCP3 | Macrophage inflammatory protein 1a levels | 2016 | Ahola-Olli AV | 27989323 | 3522 | 9519267 | 2018 | Michela Traglia | 30134952 | 764 | 652831 | HG19/GRCh37 |
| MIF | Macrophage Migration Inhibitory Factor levels | 2016 | Ahola-Olli AV | 27989323 | 3494 | 9537573 | 2018 | Michela Traglia | 30134952 | 764 | 652831 | HG19/GRCh37 |
| MIG | Monocyte chemoattractant protein-3 levels | 2016 | Ahola-Olli AV | 27989323 | 843 | 7630881 | 2018 | Michela Traglia | 30134952 | 764 | 652831 | HG19/GRCh37 |
| MIP1a | Monokine induced by gamma interferon levels | 2016 | Ahola-Olli AV | 27989323 | 3685 | 9579894 | 2018 | Michela Traglia | 30134952 | 764 | 652831 | HG19/GRCh37 |
| TNFa | Tumor necrosis factor alpha levels | 2016 | Ahola-Olli AV | 27989323 | 3454 | 9500449 | 2018 | Michela Traglia | 30134952 | 764 | 652831 | HG19/GRCh37 |

**Table S3. Datasets information of 42 eye diseases**

| trait | note | year | population | nsnp | build | category | ncase | ncontrol |
|---|---|---|---|---|---|---|---|---|
| Glaucoma, exfoliation | H7_GLAUCOMA_XFG | 2021 | European | 16380448 | HG19/GRCh37 | Binary | 1515 | 210201 |
| Disorders of vitreous body | H7_VITRBODYGLOBE | 2021 | European | 16380466 | HG19/GRCh37 | Binary | 6782 | 211720 |
| Retinal vascular disorders | H7_RETINAVASC | 2021 | European | 16380386 | HG19/GRCh37 | Binary | 1642 | 203018 |
| Disorders of ocular muscles, binocular movement, accommodation and refraction | H7_OCUMUSCLE | 2021 | European | 16380466 | HG19/GRCh37 | Binary | 7861 | 210931 |
| Disorders of refraction and accommodation | H7_REFRAACCOMMODIS | 2021 | European | 16380456 | HG19/GRCh37 | Binary | 3453 | 210931 |
| Dacryoadenitis | H7_DACRYOADENITIS | 2021 | European | 16380434 | HG19/GRCh37 | Binary | 102 | 203231 |
| Age-related macular degeneration (whether dry or wet) | H7_AMD | 2021 | European | 16380424 | HG19/GRCh37 | Binary | 3763 | 205359 |
| Papilloedema, unspecified | H7_PAPILLOEDEMA | 2021 | European | 16380465 | HG19/GRCh37 | Binary | 251 | 217491 |
| Ptosis of eyelid | H7_PTOSIS | 2021 | European | 16380436 | HG19/GRCh37 | Binary | 1349 | 203231 |
| Normotensive glaucoma | H7_GLAUCOMA_NTG | 2021 | European | 16380447 | HG19/GRCh37 | Binary | 892 | 210201 |
| Optic neuritis | H7_OPTNEURITIS | 2021 | European | 16380463 | HG19/GRCh37 | Binary | 582 | 217491 |
| Other and unspecified visual disturbances | H7_VISDISTNAS | 2021 | European | 16380454 | HG19/GRCh37 | Binary | 902 | 210866 |
| Primary open-angle glaucoma | H7_GLAUCPRIMOPEN | 2021 | European | 16380455 | HG19/GRCh37 | Binary | 4433 | 210201 |
| Other and unspecified vascular occlusions | H7_RETVASCNAS | 2021 | European | 16380386 | HG19/GRCh37 | Binary | 1240 | 203018 |
| Paralytic strabismus | H7_PARASTRAB | 2021 | European | 16380456 | HG19/GRCh37 | Binary | 918 | 210931 |
| Atopic conjunctivitis | H7_CONJUNCTIVITISATOPIC | 2021 | European | 16380437 | HG19/GRCh37 | Binary | 582 | 203517 |
| Retinal vascular occlusion | H7_RETIVASCOCCLUSION | 2021 | European | 16380386 | HG19/GRCh37 | Binary | 1595 | 203018 |
| Other and unspecified iridocyclitis | H7_IRIDONAS | 2021 | European | 16380408 | HG19/GRCh37 | Binary | 413 | 209287 |
| Disorders of lacrimal system and orbit in diseases classified elsewhere | H7_LACRIMALORBITINOTH | 2021 | European | 16380435 | HG19/GRCh37 | Binary | 351 | 203231 |
| Iridocyclitis | H7_IRIDOCYCLITIS | 2021 | European | 16380395 | HG19/GRCh37 | Binary | 3622 | 209287 |
| Central retinal artery occlusion | H7_CENTRRETARTOCC | 2021 | European | 16380385 | HG19/GRCh37 | Binary | 251 | 203018 |
| Disorders of optic nerve and visual pathways | H7_OPTNERVE | 2021 | European | 16380466 | HG19/GRCh37 | Binary | 1301 | 217491 |
| Visual field defects | H7_VISFIELDDEF | 2021 | European | 16380453 | HG19/GRCh37 | Binary | 1178 | 210866 |
| Degeneration of macula and posterior pole | H7_MACULADEGEN | 2021 | European | 16380423 | HG19/GRCh37 | Binary | 6508 | 203018 |

| | | | | | | | | |
|---|---|---|---|---|---|---|---|---|
| Disorders of sclera, cornea, iris and ciliary body | H7_SCLERACORNEA | 2021 | European | 16380466 | HG19/GRCh37 | Binary | 9505 | 209287 |
| Other, unspecified and serous retinal detachments | H7_RETINALDETACHOTH | 2021 | European | 16380385 | HG19/GRCh37 | Binary | 374 | 203018 |
| Convergent concomitant strabismus | H7_CONVERSTRAB | 2021 | European | 16380461 | HG19/GRCh37 | Binary | 967 | 210931 |
| Other specified and unspecified retinal disorders | H7_RETINANAS | 2021 | European | 16380388 | HG19/GRCh37 | Binary | 376 | 203018 |
| Disorders of orbit | H7_ORBIT | 2021 | European | 16380436 | HG19/GRCh37 | Binary | 624 | 203231 |
| Intermittent heterotropia | H7_INTERHETEROTRO | 2021 | European | 16380456 | HG19/GRCh37 | Binary | 823 | 210931 |
| Other disorders of eyelid | H7_EYELIDDIS | 2021 | European | 16380445 | HG19/GRCh37 | Binary | 6844 | 203231 |
| Ectropion of eyelid | H7_ECTROPION | 2021 | European | 16380434 | HG19/GRCh37 | Binary | 473 | 203231 |
| Disorders of vitreous body and globe | H7_VITREOUS | 2021 | European | 16380466 | HG19/GRCh37 | Binary | 7072 | 211720 |
| Corneal degeneration | H7_CORNEALDYSTROPHY | 2021 | European | 16380407 | HG19/GRCh37 | Binary | 124 | 209287 |
| Other disorders of optic [2nd] nerve and visual pathways | H7_OPTNEUROTH | 2021 | European | 16380465 | HG19/GRCh37 | Binary | 785 | 217491 |
| Other and unspecified disorders of vitreous body | H7_VITROTH | 2021 | European | 16380466 | HG19/GRCh37 | Binary | 5304 | 211720 |
| Acute and subacute iridocyclitis | H7_IRIDOACUTE | 2021 | European | 16380353 | HG19/GRCh37 | Binary | 3126 | 209287 |
| Diabetic retinopathy (H7_RETINOPATHYDIAB) | H7_RETINOPATHYDIAB | 2021 | European | 16380430 | HG19/GRCh37 | Binary | 3646 | 203018 |
| Blepharochalasis | H7_BLEPHAROCHALASIS | 2021 | European | 16380443 | HG19/GRCh37 | Binary | 4135 | 203231 |
| Other retinal disorders | H7_RETINALDISOTH | 2021 | European | 16380459 | HG19/GRCh37 | Binary | 11096 | 203018 |
| Chronic conjunctivitis | H7_CONJUNCTIVITISCHRON | 2021 | European | 16380438 | HG19/GRCh37 | Binary | 407 | 203517 |
| Strabismus | H7_STRABISMUS | 2021 | European | 16380466 | HG19/GRCh37 | Binary | 4620 | 214172 |

### 2 Results

| Methods | nSNPs | OR | CI | PVAL |
|---|---|---|---|---|
| **Gest duration->Disorders of vitreous body** | | | | |
| MR Egger (bootstrap) | 103 | 0.812 | [0.599,1.102] | 0.095 |
| Simple median | 103 | 0.802 | [0.649,0.991] | 0.041* |
| Weighted median | 103 | 0.912 | [0.729,1.141] | 0.423 |
| Penalised weighted median | 103 | 0.912 | [0.732,1.136] | 0.412 |
| IVW radial | 103 | 0.808 | [0.703,0.929] | 0.003* |
| Inverse variance weighted (multiplicative random effects) | 103 | 0.808 | [0.703,0.929] | 0.003* |
| Inverse variance weighted (fixed effects) | 103 | 0.808 | [0.697,0.938] | 0.005* |
| Simple mode | 103 | 1.014 | [0.596,1.726] | 0.96 |
| Weighted mode | 103 | 1.025 | [0.657,1.599] | 0.911 |
| Weighted mode (NOME) | 103 | 1.025 | [0.648,1.621] | 0.914 |
| Simple mode (NOME) | 103 | 1.014 | [0.578,1.78] | 0.962 |
| Robust adjusted profile score (RAPS) | 103 | 0.802 | [0.682,0.943] | 0.007* |
| MRPRESSO | 103 | 0.808 | [0.703,0.929] | 0.003* |
| **Gest duration->Dacryoadenitis** | | | | |
| MR Egger (bootstrap) | 103 | 5.16 | [0.447,59.571] | 0.098 |
| Simple median | 103 | 2.406 | [0.408,14.184] | 0.332 |
| Weighted median | 103 | 2.678 | [0.478,15.009] | 0.263 |
| Penalised weighted median | 103 | 2.635 | [0.476,14.578] | 0.267 |
| IVW radial | 103 | 3.626 | [1.061,12.384] | 0.04* |
| Inverse variance weighted (multiplicative random effects) | 103 | 3.629 | [1.063,12.389] | 0.04* |
| Inverse variance weighted (fixed effects) | 103 | 3.629 | [1.134,11.619] | 0.03* |
| Simple mode | 103 | 1.496 | [0.026,86.143] | 0.846 |
| Weighted mode | 103 | 2.044 | [0.075,55.951] | 0.673 |
| Weighted mode (NOME) | 103 | 2.268 | [0.061,84.945] | 0.659 |
| Simple mode (NOME) | 103 | 1.496 | [0.021,105.821] | 0.853 |
| Robust adjusted profile score (RAPS) | 103 | 3.532 | [0.945,13.209] | 0.061 |
| MRPRESSO | 103 | 3.629 | [1.063,12.389] | 0.042* |
| **Gest duration->Central retinal artery occlusion** | | | | |
| MR Egger (bootstrap) | 103 | 3.047 | [0.631,14.705] | 0.077 |
| Simple median | 103 | 1.992 | [0.657,6.042] | 0.224 |
| Weighted median | 103 | 2.575 | [0.825,8.042] | 0.103 |
| Penalised weighted median | 103 | 2.575 | [0.837,7.927] | 0.099 |
| IVW radial | 103 | 2.171 | [1.069,4.408] | 0.032* |
| Inverse variance weighted (multiplicative random effects) | 103 | 2.171 | [1.069,4.408] | 0.032* |
| Inverse variance weighted (fixed effects) | 103 | 2.171 | [1.032,4.564] | 0.041* |
| Simple mode | 103 | 2.514 | [0.167,37.884] | 0.507 |
| Weighted mode | 103 | 4.315 | [0.426,43.691] | 0.219 |
| Weighted mode (NOME) | 103 | 4.554 | [0.41,50.616] | 0.22 |
| Simple mode (NOME) | 103 | 2.514 | [0.142,44.594] | 0.531 |
| Robust adjusted profile score (RAPS) | 103 | 2.184 | [0.97,4.914] | 0.059 |
| MRPRESSO | 103 | 2.171 | [1.069,4.408] | 0.034* |
| **Gest duration->Visual field defects** | | | | |
| MR Egger (bootstrap) | 103 | 0.649 | [0.321,1.313] | 0.109 |
| Simple median | 103 | 0.72 | [0.43,1.205] | 0.211 |
| Weighted median | 103 | 0.725 | [0.421,1.251] | 0.249 |
| Penalised weighted median | 103 | 0.743 | [0.439,1.257] | 0.268 |
| IVW radial | 103 | 0.684 | [0.481,0.973] | 0.034* |
| Inverse variance weighted (multiplicative random effects) | 103 | 0.683 | [0.48,0.971] | 0.034* |
| Inverse variance weighted (fixed effects) | 103 | 0.683 | [0.484,0.964] | 0.03* |
| Simple mode | 103 | 0.674 | [0.173,2.622] | 0.571 |
| Weighted mode | 103 | 0.91 | [0.27,3.074] | 0.88 |
| Weighted mode (NOME) | 103 | 0.845 | [0.249,2.859] | 0.787 |
| Simple mode (NOME) | 103 | 0.674 | [0.172,2.646] | 0.574 |
| Robust adjusted profile score (RAPS) | 103 | 0.702 | [0.471,1.045] | 0.082 |
| MRPRESSO | 103 | 0.683 | [0.48,0.971] | 0.036* |
| **Gest duration->Ectropion of eyelid** | | | | |
| MR Egger (bootstrap) | 103 | 1.313 | [0.459,3.754] | 0.306 |
| Simple median | 103 | 2.428 | [1.087,5.421] | 0.03* |
| Weighted median | 103 | 2.098 | [0.922,4.776] | 0.077 |
| Penalised weighted median | 103 | 2.098 | [0.945,4.656] | 0.068 |
| IVW radial | 103 | 1.944 | [1.147,3.297] | 0.013* |
| Inverse variance weighted (multiplicative random effects) | 103 | 1.944 | [1.147,3.295] | 0.014* |
| Inverse variance weighted (fixed effects) | 103 | 1.944 | [1.127,3.355] | 0.017* |
| Simple mode | 103 | 1.347 | [0.191,9.503] | 0.766 |
| Weighted mode | 103 | 1.422 | [0.327,6.192] | 0.641 |
| Weighted mode (NOME) | 103 | 1.422 | [0.269,7.524] | 0.68 |
| Simple mode (NOME) | 103 | 1.347 | [0.159,11.44] | 0.786 |
| Robust adjusted profile score (RAPS) | 103 | 2.018 | [1.114,3.656] | 0.021* |
| MRPRESSO | 103 | 1.944 | [1.147,3.295] | 0.015* |
| **Gest duration->Disorders of vitreous body and globe** | | | | |
| MR Egger (bootstrap) | 103 | 0.816 | [0.599,1.113] | 0.093 |
| Simple median | 103 | 0.812 | [0.655,1.007] | 0.058 |
| Weighted median | 103 | 0.876 | [0.707,1.087] | 0.229 |
| Penalised weighted median | 103 | 0.875 | [0.702,1.089] | 0.232 |
| IVW radial | 103 | 0.8 | [0.696,0.92] | 0.002* |
| Inverse variance weighted (multiplicative random effects) | 103 | 0.801 | [0.697,0.92] | 0.002* |
| Inverse variance weighted (fixed effects) | 103 | 0.801 | [0.693,0.926] | 0.003* |
| Simple mode | 103 | 0.884 | [0.518,1.511] | 0.653 |
| Weighted mode | 103 | 0.935 | [0.628,1.394] | 0.744 |
| Weighted mode (NOME) | 103 | 0.935 | [0.602,1.453] | 0.768 |
| Simple mode (NOME) | 103 | 0.884 | [0.538,1.454] | 0.628 |
| Robust adjusted profile score (RAPS) | 103 | 0.795 | [0.678,0.931] | 0.005* |
| MRPRESSO | 103 | 0.801 | [0.697,0.92] | 0.002* |
| **Gest duration->Other and unspecified disorders of vitreous body** | | | | |
| MR Egger (bootstrap) | 103 | 0.806 | [0.569,1.142] | 0.112 |
| Simple median | 103 | 0.79 | [0.615,1.013] | 0.064 |
| Weighted median | 103 | 0.803 | [0.627,1.028] | 0.082 |
| Penalised weighted median | 103 | 0.803 | [0.628,1.027] | 0.08 |
| IVW radial | 103 | 0.796 | [0.681,0.931] | 0.004* |
| Inverse variance weighted (multiplicative random effects) | 103 | 0.796 | [0.681,0.93] | 0.004* |
| Inverse variance weighted (fixed effects) | 103 | 0.796 | [0.673,0.942] | 0.008* |
| Simple mode | 103 | 0.807 | [0.45,1.45] | 0.476 |
| Weighted mode | 103 | 0.796 | [0.485,1.307] | 0.37 |
| Weighted mode (NOME) | 103 | 0.796 | [0.48,1.321] | 0.38 |
| Simple mode (NOME) | 103 | 0.807 | [0.454,1.435] | 0.469 |
| Robust adjusted profile score (RAPS) | 103 | 0.779 | [0.649,0.935] | 0.007* |
| MRPRESSO | 103 | 0.796 | [0.681,0.93] | 0.005* |

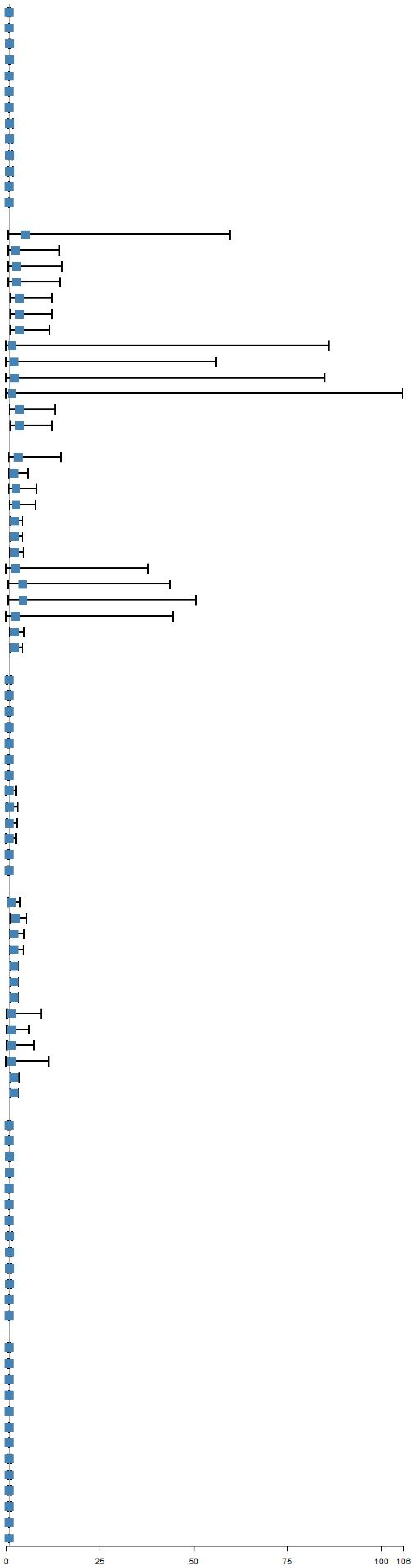

**Fig S1.1 UVMR results of gest duration on adult eye diseases.**

| Methods | nSNPs | OR | CI | PVAL |
|---|---|---|---|---|
| **Preterm birth->Glaucoma, exfoliation** | | | | |
| MR Egger (bootstrap) | 73 | 1.019 | [0.858,1.211] | 0.412 |
| Simple median | 73 | 1.126 | [0.99,1.281] | 0.069 |
| Weighted median | 73 | 1.125 | [0.997,1.27] | 0.056 |
| Penalised weighted median | 73 | 1.126 | [0.999,1.271] | 0.054 |
| IVW radial | 73 | 1.103 | [1.012,1.202] | 0.025* |
| Inverse variance weighted (multiplicative random effects) | 73 | 1.103 | [1.012,1.202] | 0.025* |
| Inverse variance weighted (fixed effects) | 73 | 1.103 | [1.017,1.196] | 0.017* |
| Simple mode | 73 | 1.135 | [0.84,1.535] | 0.414 |
| Weighted mode | 73 | 1.149 | [0.896,1.474] | 0.276 |
| Weighted mode (NOME) | 73 | 1.157 | [0.881,1.519] | 0.299 |
| Simple mode (NOME) | 73 | 1.135 | [0.846,1.524] | 0.402 |
| Robust adjusted profile score (RAPS) | 73 | 1.122 | [1.012,1.244] | 0.029* |
| MRPRESSO | 73 | 1.103 | [1.012,1.202] | 0.028* |
| **Preterm birth->Disorders of ocular muscles, binocular movement, accommodation and refraction** | | | | |
| MR Egger (bootstrap) | 73 | 1.042 | [0.964,1.127] | 0.151 |
| Simple median | 73 | 1.074 | [1.018,1.133] | 0.01* |
| Weighted median | 73 | 1.049 | [0.996,1.105] | 0.067 |
| Penalised weighted median | 73 | 1.049 | [0.992,1.109] | 0.09 |
| IVW radial | 73 | 1.061 | [1.022,1.101] | 0.002* |
| Inverse variance weighted (multiplicative random effects) | 73 | 1.061 | [1.022,1.101] | 0.002* |
| Inverse variance weighted (fixed effects) | 73 | 1.061 | [1.024,1.099] | 0.001* |
| Simple mode | 73 | 1.081 | [0.957,1.222] | 0.214 |
| Weighted mode | 73 | 1.064 | [0.969,1.168] | 0.196 |
| Weighted mode (NOME) | 73 | 1.067 | [0.969,1.175] | 0.191 |
| Simple mode (NOME) | 73 | 1.081 | [0.956,1.222] | 0.215 |
| Robust adjusted profile score (RAPS) | 73 | 1.06 | [1.017,1.104] | 0.005* |
| MRPRESSO | 73 | 1.061 | [1.022,1.101] | 0.002* |
| **Preterm birth->Disorders of refraction and accommodation** | | | | |
| MR Egger (bootstrap) | 73 | 1.009 | [0.907,1.122] | 0.444 |
| Simple median | 73 | 1.101 | [1.017,1.192] | 0.018* |
| Weighted median | 73 | 1.051 | [0.974,1.135] | 0.199 |
| Penalised weighted median | 73 | 1.05 | [0.97,1.137] | 0.226 |
| IVW radial | 73 | 1.074 | [1.016,1.134] | 0.012* |
| Inverse variance weighted (multiplicative random effects) | 73 | 1.073 | [1.015,1.133] | 0.012* |
| Inverse variance weighted (fixed effects) | 73 | 1.073 | [1.018,1.13] | 0.008* |
| Simple mode | 73 | 1.101 | [0.931,1.302] | 0.266 |
| Weighted mode | 73 | 1.021 | [0.885,1.178] | 0.778 |
| Weighted mode (NOME) | 73 | 1.024 | [0.882,1.189] | 0.752 |
| Simple mode (NOME) | 73 | 1.101 | [0.914,1.326] | 0.316 |
| Robust adjusted profile score (RAPS) | 73 | 1.071 | [1.008,1.139] | 0.027* |
| MRPRESSO | 73 | 1.073 | [1.015,1.133] | 0.014* |
| **Preterm birth->Ptosis of eyelid** | | | | |
| MR Egger (bootstrap) | 73 | 0.908 | [0.759,1.085] | 0.145 |
| Simple median | 73 | 0.952 | [0.838,1.081] | 0.451 |
| Weighted median | 73 | 0.892 | [0.789,1.01] | 0.071 |
| Penalised weighted median | 73 | 0.888 | [0.783,1.006] | 0.063 |
| IVW radial | 73 | 0.913 | [0.835,0.998] | 0.044* |
| Inverse variance weighted (multiplicative random effects) | 73 | 0.913 | [0.835,0.998] | 0.044* |
| Inverse variance weighted (fixed effects) | 73 | 0.913 | [0.84,0.992] | 0.031* |
| Simple mode | 73 | 0.942 | [0.706,1.256] | 0.685 |
| Weighted mode | 73 | 0.923 | [0.742,1.148] | 0.472 |
| Weighted mode (NOME) | 73 | 0.917 | [0.733,1.146] | 0.447 |
| Simple mode (NOME) | 73 | 0.942 | [0.709,1.25] | 0.68 |
| Robust adjusted profile score (RAPS) | 73 | 0.907 | [0.823,0.999] | 0.046* |
| MRPRESSO | 73 | 0.913 | [0.835,0.998] | 0.048* |
| **Preterm birth->Normotensive glaucoma** | | | | |
| MR Egger (bootstrap) | 73 | 1.093 | [0.883,1.353] | 0.215 |
| Simple median | 73 | 1.057 | [0.91,1.226] | 0.474 |
| Weighted median | 73 | 1.076 | [0.926,1.249] | 0.341 |
| Penalised weighted median | 73 | 1.075 | [0.924,1.25] | 0.349 |
| IVW radial | 73 | 1.104 | [1.001,1.218] | 0.048* |
| Inverse variance weighted (multiplicative random effects) | 73 | 1.104 | [1.001,1.218] | 0.048* |
| Inverse variance weighted (fixed effects) | 73 | 1.104 | [0.996,1.224] | 0.061 |
| Simple mode | 73 | 0.998 | [0.71,1.404] | 0.992 |
| Weighted mode | 73 | 1.039 | [0.782,1.38] | 0.795 |
| Weighted mode (NOME) | 73 | 1.039 | [0.782,1.38] | 0.795 |
| Simple mode (NOME) | 73 | 0.998 | [0.708,1.407] | 0.992 |
| Robust adjusted profile score (RAPS) | 73 | 1.101 | [0.984,1.232] | 0.095 |
| MRPRESSO | 73 | 1.104 | [1.001,1.218] | 0.052 |
| **Preterm birth->Other and unspecified visual disturbances** | | | | |
| MR Egger (bootstrap) | 73 | 0.798 | [0.64,0.995] | 0.03* |
| Simple median | 73 | 0.932 | [0.798,1.089] | 0.376 |
| Weighted median | 73 | 0.959 | [0.824,1.116] | 0.592 |
| Penalised weighted median | 73 | 0.97 | [0.832,1.131] | 0.706 |
| IVW radial | 73 | 0.87 | [0.786,0.964] | 0.007* |
| Inverse variance weighted (multiplicative random effects) | 73 | 0.869 | [0.785,0.963] | 0.007* |
| Inverse variance weighted (fixed effects) | 73 | 0.869 | [0.786,0.961] | 0.006* |
| Simple mode | 73 | 0.99 | [0.7,1.401] | 0.956 |
| Weighted mode | 73 | 0.998 | [0.736,1.353] | 0.991 |
| Weighted mode (NOME) | 73 | 0.998 | [0.713,1.396] | 0.991 |
| Simple mode (NOME) | 73 | 0.99 | [0.691,1.419] | 0.958 |
| Robust adjusted profile score (RAPS) | 73 | 0.888 | [0.793,0.994] | 0.04* |
| MRPRESSO | 73 | 0.869 | [0.785,0.963] | 0.009* |
| **Preterm birth->Primary open-angle glaucoma** | | | | |
| MR Egger (bootstrap) | 73 | 1.011 | [0.918,1.114] | 0.39 |
| Simple median | 73 | 1.095 | [1.016,1.181] | 0.018* |
| Weighted median | 73 | 1.051 | [0.976,1.132] | 0.189 |
| Penalised weighted median | 73 | 1.05 | [0.976,1.13] | 0.19 |
| IVW radial | 73 | 1.064 | [1.01,1.121] | 0.019* |
| Inverse variance weighted (multiplicative random effects) | 73 | 1.064 | [1.01,1.121] | 0.019* |
| Inverse variance weighted (fixed effects) | 73 | 1.064 | [1.013,1.117] | 0.012* |
| Simple mode | 73 | 1.078 | [0.908,1.28] | 0.394 |
| Weighted mode | 73 | 1.07 | [0.93,1.232] | 0.343 |
| Weighted mode (NOME) | 73 | 1.078 | [0.934,1.244] | 0.308 |
| Simple mode (NOME) | 73 | 1.078 | [0.905,1.284] | 0.402 |
| Robust adjusted profile score (RAPS) | 73 | 1.068 | [1.008,1.132] | 0.026* |
| MRPRESSO | 73 | 1.064 | [1.01,1.121] | 0.022* |
| **Preterm birth->Paralytic strabismus** | | | | |
| MR Egger (bootstrap) | 73 | 1.156 | [0.929,1.439] | 0.098 |
| Simple median | 73 | 1.133 | [0.969,1.325] | 0.116 |
| Weighted median | 73 | 1.168 | [0.998,1.366] | 0.052 |
| Penalised weighted median | 73 | 1.165 | [0.999,1.359] | 0.05 |
| IVW radial | 73 | 1.138 | [1.011,1.28] | 0.032* |
| Inverse variance weighted (multiplicative random effects) | 73 | 1.138 | [1.011,1.28] | 0.032* |
| Inverse variance weighted (fixed effects) | 73 | 1.138 | [1.029,1.257] | 0.011* |
| Simple mode | 73 | 1.184 | [0.83,1.689] | 0.354 |
| Weighted mode | 73 | 1.207 | [0.9,1.618] | 0.213 |
| Weighted mode (NOME) | 73 | 1.218 | [0.915,1.62] | 0.181 |
| Simple mode (NOME) | 73 | 1.184 | [0.811,1.729] | 0.384 |
| Robust adjusted profile score (RAPS) | 73 | 1.119 | [0.986,1.268] | 0.08 |
| MRPRESSO | 73 | 1.138 | [1.011,1.28] | 0.035* |
| **Preterm birth->Disorders of sclera, cornea, iris and ciliary body** | | | | |
| MR Egger (bootstrap) | 73 | 1.016 | [0.949,1.088] | 0.321 |
| Simple median | 73 | 1.035 | [0.987,1.084] | 0.159 |
| Weighted median | 73 | 1.022 | [0.975,1.072] | 0.369 |
| Penalised weighted median | 73 | 1.022 | [0.975,1.072] | 0.369 |
| IVW radial | 73 | 1.029 | [1.001,1.058] | 0.043* |
| Inverse variance weighted (multiplicative random effects) | 73 | 1.029 | [1.001,1.058] | 0.043* |
| Inverse variance weighted (fixed effects) | 73 | 1.029 | [0.997,1.063] | 0.081 |
| Simple mode | 73 | 1.02 | [0.923,1.128] | 0.691 |
| Weighted mode | 73 | 1.014 | [0.935,1.1] | 0.733 |
| Weighted mode (NOME) | 73 | 1.014 | [0.937,1.098] | 0.727 |
| Simple mode (NOME) | 73 | 1.02 | [0.924,1.126] | 0.686 |
| Robust adjusted profile score (RAPS) | 73 | 1.034 | [0.998,1.071] | 0.069 |
| MRPRESSO | 73 | 1.029 | [1.001,1.058] | 0.046* |
| **Preterm birth->Diabetic retinopathy (H7_RETINOPATHYDIAB)** | | | | |
| MR Egger (bootstrap) | 73 | 0.972 | [0.871,1.086] | 0.316 |
| Simple median | 73 | 0.947 | [0.873,1.028] | 0.191 |
| Weighted median | 73 | 0.95 | [0.88,1.026] | 0.194 |
| Penalised weighted median | 73 | 0.95 | [0.879,1.027] | 0.2 |
| IVW radial | 73 | 0.95 | [0.905,0.998] | 0.041* |
| Inverse variance weighted (multiplicative random effects) | 73 | 0.95 | [0.905,0.998] | 0.041* |
| Inverse variance weighted (fixed effects) | 73 | 0.95 | [0.901,1.002] | 0.059 |
| Simple mode | 73 | 0.932 | [0.777,1.119] | 0.456 |
| Weighted mode | 73 | 0.96 | [0.816,1.129] | 0.625 |
| Weighted mode (NOME) | 73 | 0.96 | [0.808,1.14] | 0.646 |
| Simple mode (NOME) | 73 | 0.932 | [0.773,1.124] | 0.466 |
| Robust adjusted profile score (RAPS) | 73 | 0.948 | [0.895,1.005] | 0.074 |
| MRPRESSO | 73 | 0.95 | [0.905,0.998] | 0.045* |
| **Preterm birth->Strabismus** | | | | |
| MR Egger (bootstrap) | 73 | 1.066 | [0.969,1.173] | 0.09 |
| Simple median | 73 | 1.026 | [0.959,1.099] | 0.458 |
| Weighted median | 73 | 1.042 | [0.977,1.111] | 0.216 |
| Penalised weighted median | 73 | 1.042 | [0.973,1.115] | 0.241 |
| IVW radial | 73 | 1.052 | [1.005,1.102] | 0.03* |
| Inverse variance weighted (multiplicative random effects) | 73 | 1.052 | [1.005,1.102] | 0.03* |
| Inverse variance weighted (fixed effects) | 73 | 1.052 | [1.006,1.101] | 0.027* |
| Simple mode | 73 | 1.015 | [0.892,1.156] | 0.815 |
| Weighted mode | 73 | 1.03 | [0.931,1.14] | 0.567 |
| Weighted mode (NOME) | 73 | 1.034 | [0.926,1.154] | 0.557 |
| Simple mode (NOME) | 73 | 1.015 | [0.89,1.158] | 0.819 |
| Robust adjusted profile score (RAPS) | 73 | 1.054 | [1.004,1.108] | 0.035* |
| MRPRESSO | 73 | 1.052 | [1.005,1.102] | 0.034* |

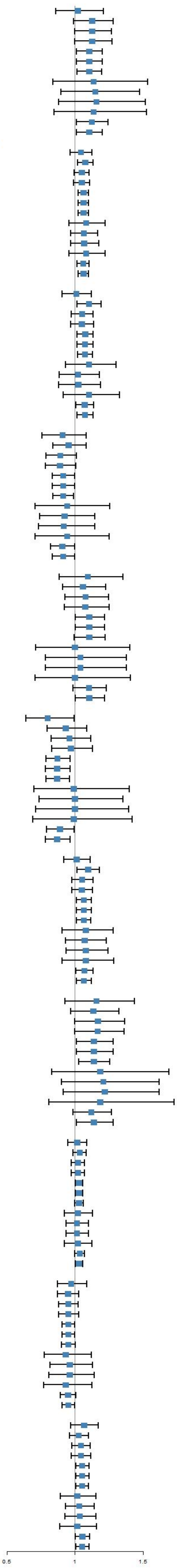

**Fig S1.2 UVMR results of preterm birth on adult eye diseases.**

| Methods | nSNPs | OR | CI | PVAL |
|---|---|---|---|---|
| **Postterm birth->Optic neuritis** | | | | |
| MR Egger (bootstrap) | 68 | 1.102 | [0.824,1.474] | 0.251 |
| Simple median | 68 | 1.151 | [0.923,1.436] | 0.212 |
| Weighted median | 68 | 1.234 | [0.996,1.528] | 0.055 |
| Penalised weighted median | 68 | 1.236 | [0.994,1.537] | 0.056 |
| IVW radial | 68 | 1.169 | [1.019,1.341] | 0.026* |
| Inverse variance weighted (multiplicative random effects) | 68 | 1.169 | [1.019,1.34] | 0.025* |
| Inverse variance weighted (fixed effects) | 68 | 1.169 | [1.013,1.349] | 0.033* |
| Simple mode | 68 | 1.398 | [0.902,2.168] | 0.139 |
| Weighted mode | 68 | 1.26 | [0.877,1.809] | 0.216 |
| Weighted mode (NOME) | 68 | 1.271 | [0.896,1.804] | 0.183 |
| Simple mode (NOME) | 68 | 1.398 | [0.893,2.189] | 0.148 |
| Robust adjusted profile score (RAPS) | 68 | 1.202 | [1.028,1.406] | 0.021* |
| MRPRESSO | 68 | 1.169 | [1.019,1.34] | 0.029* |
| **Postterm birth->Atopic conjunctivitis** | | | | |
| MR Egger (bootstrap) | 68 | 1.13 | [0.852,1.498] | 0.19 |
| Simple median | 68 | 1.158 | [0.935,1.436] | 0.181 |
| Weighted median | 68 | 1.097 | [0.886,1.36] | 0.396 |
| Penalised weighted median | 68 | 1.095 | [0.876,1.369] | 0.425 |
| IVW radial | 68 | 1.212 | [1.05,1.398] | 0.009* |
| Inverse variance weighted (multiplicative random effects) | 68 | 1.21 | [1.049,1.396] | 0.009* |
| Inverse variance weighted (fixed effects) | 68 | 1.21 | [1.049,1.396] | 0.009* |
| Simple mode | 68 | 1.039 | [0.686,1.573] | 0.86 |
| Weighted mode | 68 | 1.051 | [0.726,1.523] | 0.793 |
| Weighted mode (NOME) | 68 | 1.051 | [0.735,1.504] | 0.786 |
| Simple mode (NOME) | 68 | 1.039 | [0.669,1.613] | 0.868 |
| Robust adjusted profile score (RAPS) | 68 | 1.203 | [1.029,1.407] | 0.02* |
| MRPRESSO | 68 | 1.21 | [1.049,1.396] | 0.011* |
| **Postterm birth->Disorders of lacrimal system and orbit in diseases classified elsewhere** | | | | |
| MR Egger (bootstrap) | 68 | 1.388 | [0.954,2.021] | 0.043* |
| Simple median | 68 | 1.15 | [0.874,1.515] | 0.318 |
| Weighted median | 68 | 1.223 | [0.927,1.612] | 0.154 |
| Penalised weighted median | 68 | 1.21 | [0.91,1.61] | 0.188 |
| IVW radial | 68 | 1.334 | [1.113,1.598] | 0.002* |
| Inverse variance weighted (multiplicative random effects) | 68 | 1.335 | [1.115,1.599] | 0.002* |
| Inverse variance weighted (fixed effects) | 68 | 1.335 | [1.112,1.603] | 0.002* |
| Simple mode | 68 | 1.079 | [0.583,1.997] | 0.811 |
| Weighted mode | 68 | 1.194 | [0.747,1.908] | 0.462 |
| Weighted mode (NOME) | 68 | 1.209 | [0.751,1.946] | 0.438 |
| Simple mode (NOME) | 68 | 1.079 | [0.577,2.018] | 0.814 |
| Robust adjusted profile score (RAPS) | 68 | 1.315 | [1.073,1.612] | 0.008* |
| MRPRESSO | 68 | 1.335 | [1.115,1.599] | 0.003* |
| **Postterm birth->Other, unspecified and serous retinal detachments** | | | | |
| MR Egger (bootstrap) | 68 | 1.122 | [0.787,1.599] | 0.262 |
| Simple median | 68 | 0.855 | [0.655,1.115] | 0.248 |
| Weighted median | 68 | 0.859 | [0.664,1.111] | 0.248 |
| Penalised weighted median | 68 | 0.859 | [0.659,1.12] | 0.262 |
| IVW radial | 68 | 0.836 | [0.718,0.973] | 0.02* |
| Inverse variance weighted (multiplicative random effects) | 68 | 0.836 | [0.719,0.973] | 0.021* |
| Inverse variance weighted (fixed effects) | 68 | 0.836 | [0.7,0.999] | 0.048* |
| Simple mode | 68 | 0.931 | [0.539,1.609] | 0.799 |
| Weighted mode | 68 | 0.906 | [0.59,1.391] | 0.652 |
| Weighted mode (NOME) | 68 | 0.906 | [0.591,1.389] | 0.652 |
| Simple mode (NOME) | 68 | 0.931 | [0.551,1.575] | 0.791 |
| Robust adjusted profile score (RAPS) | 68 | 0.849 | [0.698,1.031] | 0.099 |
| MRPRESSO | 68 | 0.836 | [0.719,0.973] | 0.024* |
| **Postterm birth->Convergent concomitant strabismus** | | | | |
| MR Egger (bootstrap) | 68 | 0.815 | [0.662,1.003] | 0.03* |
| Simple median | 68 | 0.937 | [0.793,1.107] | 0.442 |
| Weighted median | 68 | 0.927 | [0.785,1.094] | 0.372 |
| Penalised weighted median | 68 | 0.927 | [0.788,1.09] | 0.362 |
| IVW radial | 68 | 0.894 | [0.816,0.979] | 0.016* |
| Inverse variance weighted (multiplicative random effects) | 68 | 0.894 | [0.816,0.979] | 0.016* |
| Inverse variance weighted (fixed effects) | 68 | 0.894 | [0.799,1] | 0.05 |
| Simple mode | 68 | 0.936 | [0.678,1.293] | 0.688 |
| Weighted mode | 68 | 0.947 | [0.729,1.231] | 0.69 |
| Weighted mode (NOME) | 68 | 0.947 | [0.722,1.243] | 0.701 |
| Simple mode (NOME) | 68 | 0.936 | [0.681,1.286] | 0.683 |
| Robust adjusted profile score (RAPS) | 68 | 0.882 | [0.779,0.997] | 0.045* |
| MRPRESSO | 68 | 0.894 | [0.816,0.979] | 0.018* |
| **Postterm birth->Other specified and unspecified retinal disorders** | | | | |
| MR Egger (bootstrap) | 68 | 1.399 | [0.989,1.98] | 0.034* |
| Simple median | 68 | 1.141 | [0.87,1.497] | 0.34 |
| Weighted median | 68 | 1.328 | [1.016,1.736] | 0.038* |
| Penalised weighted median | 68 | 1.33 | [1.021,1.732] | 0.035* |
| IVW radial | 68 | 1.201 | [1.026,1.405] | 0.022* |
| Inverse variance weighted (multiplicative random effects) | 68 | 1.201 | [1.026,1.405] | 0.022* |
| Inverse variance weighted (fixed effects) | 68 | 1.201 | [1.005,1.435] | 0.044* |
| Simple mode | 68 | 1.29 | [0.744,2.239] | 0.367 |
| Weighted mode | 68 | 1.446 | [0.918,2.278] | 0.117 |
| Weighted mode (NOME) | 68 | 1.464 | [0.946,2.264] | 0.091 |
| Simple mode (NOME) | 68 | 1.29 | [0.749,2.222] | 0.36 |
| Robust adjusted profile score (RAPS) | 68 | 1.226 | [1.009,1.49] | 0.04* |
| MRPRESSO | 68 | 1.201 | [1.026,1.405] | 0.025* |
| **Postterm birth->Disorders of orbit** | | | | |
| MR Egger (bootstrap) | 68 | 1.201 | [0.908,1.589] | 0.106 |
| Simple median | 68 | 1.259 | [1.015,1.561] | 0.036* |
| Weighted median | 68 | 1.256 | [1.019,1.549] | 0.033* |
| Penalised weighted median | 68 | 1.255 | [1.021,1.542] | 0.03* |
| IVW radial | 68 | 1.231 | [1.073,1.413] | 0.003* |
| Inverse variance weighted (multiplicative random effects) | 68 | 1.231 | [1.073,1.412] | 0.003* |
| Inverse variance weighted (fixed effects) | 68 | 1.231 | [1.073,1.412] | 0.003* |
| Simple mode | 68 | 1.478 | [0.972,2.248] | 0.072 |
| Weighted mode | 68 | 1.478 | [1.023,2.137] | 0.041* |
| Weighted mode (NOME) | 68 | 1.505 | [1.083,2.092] | 0.018* |
| Simple mode (NOME) | 68 | 1.478 | [0.966,2.262] | 0.076 |
| Robust adjusted profile score (RAPS) | 68 | 1.241 | [1.066,1.444] | 0.005* |
| MRPRESSO | 68 | 1.231 | [1.073,1.412] | 0.004* |
| **Postterm birth->Intermittent heterotropia** | | | | |
| MR Egger (bootstrap) | 68 | 0.78 | [0.615,0.991] | 0.03* |
| Simple median | 68 | 0.992 | [0.825,1.193] | 0.933 |
| Weighted median | 68 | 0.943 | [0.781,1.138] | 0.538 |
| Penalised weighted median | 68 | 0.943 | [0.785,1.133] | 0.528 |
| IVW radial | 68 | 0.884 | [0.786,0.995] | 0.041* |
| Inverse variance weighted (multiplicative random effects) | 68 | 0.884 | [0.786,0.995] | 0.041* |
| Inverse variance weighted (fixed effects) | 68 | 0.884 | [0.784,0.997] | 0.045* |
| Simple mode | 68 | 1.004 | [0.696,1.449] | 0.983 |
| Weighted mode | 68 | 0.969 | [0.728,1.288] | 0.825 |
| Weighted mode (NOME) | 68 | 0.96 | [0.686,1.343] | 0.81 |
| Simple mode (NOME) | 68 | 1.004 | [0.686,1.469] | 0.984 |
| Robust adjusted profile score (RAPS) | 68 | 0.899 | [0.789,1.025] | 0.112 |
| MRPRESSO | 68 | 0.884 | [0.786,0.995] | 0.044* |
| **Postterm birth->Corneal degeneration** | | | | |
| MR Egger (bootstrap) | 68 | 0.818 | [0.444,1.508] | 0.252 |
| Simple median | 68 | 0.6 | [0.376,0.959] | 0.033* |
| Weighted median | 68 | 0.966 | [0.608,1.534] | 0.884 |
| Penalised weighted median | 68 | 0.966 | [0.609,1.532] | 0.881 |
| IVW radial | 68 | 0.736 | [0.562,0.966] | 0.027* |
| Inverse variance weighted (multiplicative random effects) | 68 | 0.737 | [0.562,0.966] | 0.027* |
| Inverse variance weighted (fixed effects) | 68 | 0.737 | [0.542,1.003] | 0.052 |
| Simple mode | 68 | 0.906 | [0.356,2.305] | 0.836 |
| Weighted mode | 68 | 1.008 | [0.48,2.118] | 0.983 |
| Weighted mode (NOME) | 68 | 1.008 | [0.453,2.245] | 0.985 |
| Simple mode (NOME) | 68 | 0.906 | [0.359,2.282] | 0.834 |
| Robust adjusted profile score (RAPS) | 68 | 0.698 | [0.498,0.978] | 0.036* |
| MRPRESSO | 68 | 0.737 | [0.562,0.966] | 0.031* |
| **Postterm birth->Other retinal disorders** | | | | |
| MR Egger (bootstrap) | 68 | 1.067 | [0.993,1.147] | 0.035* |
| Simple median | 68 | 1.024 | [0.97,1.082] | 0.395 |
| Weighted median | 68 | 1.026 | [0.972,1.083] | 0.35 |
| Penalised weighted median | 68 | 1.026 | [0.973,1.082] | 0.344 |
| IVW radial | 68 | 1.04 | [1.002,1.079] | 0.04* |
| Inverse variance weighted (multiplicative random effects) | 68 | 1.04 | [1.002,1.079] | 0.04* |
| Inverse variance weighted (fixed effects) | 68 | 1.04 | [1.004,1.077] | 0.031* |
| Simple mode | 68 | 1.028 | [0.915,1.156] | 0.64 |
| Weighted mode | 68 | 1.028 | [0.926,1.142] | 0.603 |
| Weighted mode (NOME) | 68 | 1.028 | [0.92,1.149] | 0.623 |
| Simple mode (NOME) | 68 | 1.028 | [0.917,1.154] | 0.634 |
| Robust adjusted profile score (RAPS) | 68 | 1.039 | [0.995,1.085] | 0.083 |
| MRPRESSO | 68 | 1.04 | [1.002,1.079] | 0.044* |

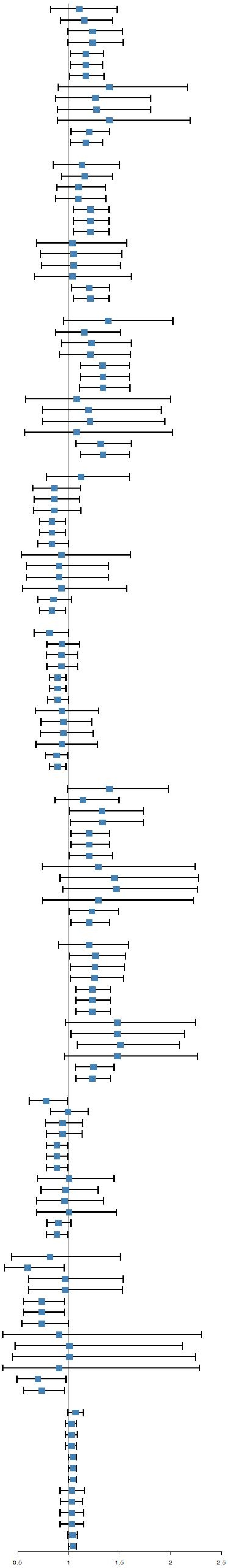

**Fig S1.3 UVMR results of postterm birth on adult eye diseases.**

| Methods | nSNPs | OR | CI | PVAL |
|---|---|---|---|---|
| **Early preterm birth->Retinal vascular disorders** | | | | |
| MR Egger (bootstrap) | 77 | 0.915 | [0.848,0.987] | 0.011* |
| Simple median | 77 | 0.937 | [0.888,0.989] | 0.018* |
| Weighted median | 77 | 0.969 | [0.918,1.021] | 0.231 |
| Penalised weighted median | 77 | 0.969 | [0.918,1.022] | 0.239 |
| IVW radial | 77 | 0.958 | [0.927,0.99] | 0.011* |
| Inverse variance weighted (multiplicative random effects) | 77 | 0.958 | [0.927,0.99] | 0.011* |
| Inverse variance weighted (fixed effects) | 77 | 0.958 | [0.925,0.993] | 0.018* |
| Simple mode | 77 | 0.908 | [0.812,1.014] | 0.091 |
| Weighted mode | 77 | 0.95 | [0.868,1.041] | 0.272 |
| Weighted mode (NOME) | 77 | 0.947 | [0.87,1.032] | 0.216 |
| Simple mode (NOME) | 77 | 0.908 | [0.81,1.017] | 0.098 |
| Robust adjusted profile score (RAPS) | 77 | 0.953 | [0.917,0.991] | 0.015* |
| MRPRESSO | 77 | 0.958 | [0.927,0.99] | 0.013* |
| **Early preterm birth->Age-related macular degeneration (whether dry or wet)** | | | | |
| MR Egger (bootstrap) | 77 | 0.984 | [0.931,1.04] | 0.281 |
| Simple median | 77 | 0.966 | [0.93,1.003] | 0.073 |
| Weighted median | 77 | 0.969 | [0.932,1.006] | 0.097 |
| Penalised weighted median | 77 | 0.969 | [0.932,1.006] | 0.099 |
| IVW radial | 77 | 0.964 | [0.939,0.989] | 0.004* |
| Inverse variance weighted (multiplicative random effects) | 77 | 0.963 | [0.938,0.988] | 0.004* |
| Inverse variance weighted (fixed effects) | 77 | 0.963 | [0.939,0.987] | 0.003* |
| Simple mode | 77 | 0.967 | [0.894,1.046] | 0.402 |
| Weighted mode | 77 | 0.954 | [0.885,1.028] | 0.221 |
| Weighted mode (NOME) | 77 | 0.949 | [0.884,1.02] | 0.155 |
| Simple mode (NOME) | 77 | 0.967 | [0.88,1.062] | 0.483 |
| Robust adjusted profile score (RAPS) | 77 | 0.961 | [0.934,0.988] | 0.005* |
| MRPRESSO | 77 | 0.963 | [0.938,0.988] | 0.005* |
| **Early preterm birth->Papilloedema, unspecified** | | | | |
| MR Egger (bootstrap) | 77 | 0.815 | [0.664,1.002] | 0.02* |
| Simple median | 77 | 0.889 | [0.772,1.023] | 0.1 |
| Weighted median | 77 | 0.887 | [0.77,1.022] | 0.097 |
| Penalised weighted median | 77 | 0.887 | [0.77,1.021] | 0.095 |
| IVW radial | 77 | 0.867 | [0.793,0.947] | 0.001* |
| Inverse variance weighted (multiplicative random effects) | 77 | 0.867 | [0.793,0.947] | 0.001* |
| Inverse variance weighted (fixed effects) | 77 | 0.867 | [0.792,0.948] | 0.002* |
| Simple mode | 77 | 0.828 | [0.607,1.129] | 0.236 |
| Weighted mode | 77 | 0.833 | [0.652,1.064] | 0.147 |
| Weighted mode (NOME) | 77 | 0.822 | [0.644,1.049] | 0.12 |
| Simple mode (NOME) | 77 | 0.828 | [0.593,1.156] | 0.27 |
| Robust adjusted profile score (RAPS) | 77 | 0.858 | [0.774,0.952] | 0.004* |
| MRPRESSO | 77 | 0.867 | [0.793,0.947] | 0.002* |
| **Early preterm birth->Other and unspecified vascular occlusions** | | | | |
| MR Egger (bootstrap) | 77 | 0.901 | [0.825,0.984] | 0.008* |
| Simple median | 77 | 0.928 | [0.868,0.991] | 0.027* |
| Weighted median | 77 | 0.956 | [0.895,1.021] | 0.18 |
| Penalised weighted median | 77 | 0.956 | [0.896,1.02] | 0.179 |
| IVW radial | 77 | 0.953 | [0.915,0.993] | 0.02* |
| Inverse variance weighted (multiplicative random effects) | 77 | 0.953 | [0.915,0.993] | 0.02* |
| Inverse variance weighted (fixed effects) | 77 | 0.953 | [0.915,0.993] | 0.02* |
| Simple mode | 77 | 0.919 | [0.793,1.067] | 0.271 |
| Weighted mode | 77 | 0.959 | [0.853,1.078] | 0.487 |
| Weighted mode (NOME) | 77 | 0.956 | [0.855,1.069] | 0.435 |
| Simple mode (NOME) | 77 | 0.919 | [0.79,1.07] | 0.281 |
| Robust adjusted profile score (RAPS) | 77 | 0.949 | [0.907,0.993] | 0.024* |
| MRPRESSO | 77 | 0.953 | [0.915,0.993] | 0.022* |
| **Early preterm birth->Retinal vascular occlusion** | | | | |
| MR Egger (bootstrap) | 77 | 0.919 | [0.848,0.995] | 0.014* |
| Simple median | 77 | 0.958 | [0.91,1.009] | 0.099 |
| Weighted median | 77 | 0.974 | [0.922,1.03] | 0.361 |
| Penalised weighted median | 77 | 0.974 | [0.925,1.027] | 0.335 |
| IVW radial | 77 | 0.96 | [0.928,0.992] | 0.016* |
| Inverse variance weighted (multiplicative random effects) | 77 | 0.96 | [0.928,0.992] | 0.016* |
| Inverse variance weighted (fixed effects) | 77 | 0.96 | [0.926,0.995] | 0.027* |
| Simple mode | 77 | 0.911 | [0.818,1.015] | 0.096 |
| Weighted mode | 77 | 0.96 | [0.881,1.046] | 0.348 |
| Weighted mode (NOME) | 77 | 0.957 | [0.875,1.046] | 0.332 |
| Simple mode (NOME) | 77 | 0.911 | [0.814,1.02] | 0.111 |
| Robust adjusted profile score (RAPS) | 77 | 0.955 | [0.918,0.993] | 0.024* |
| MRPRESSO | 77 | 0.96 | [0.928,0.992] | 0.019* |
| **Early preterm birth->Other and unspecified iridocyclitis** | | | | |
| MR Egger (bootstrap) | 77 | 1.001 | [0.858,1.168] | 0.504 |
| Simple median | 77 | 1.092 | [0.983,1.213] | 0.098 |
| Weighted median | 77 | 1.076 | [0.969,1.194] | 0.171 |
| Penalised weighted median | 77 | 1.076 | [0.972,1.19] | 0.16 |
| IVW radial | 77 | 1.093 | [1.032,1.158] | 0.002* |
| Inverse variance weighted (multiplicative random effects) | 77 | 1.093 | [1.032,1.158] | 0.002* |
| Inverse variance weighted (fixed effects) | 77 | 1.093 | [1.019,1.173] | 0.013* |
| Simple mode | 77 | 1.133 | [0.907,1.416] | 0.275 |
| Weighted mode | 77 | 1.108 | [0.928,1.324] | 0.259 |
| Weighted mode (NOME) | 77 | 1.115 | [0.926,1.343] | 0.255 |
| Simple mode (NOME) | 77 | 1.133 | [0.912,1.408] | 0.262 |
| Robust adjusted profile score (RAPS) | 77 | 1.094 | [1.013,1.182] | 0.022* |
| MRPRESSO | 77 | 1.093 | [1.032,1.158] | 0.003* |
| **Early preterm birth->Iridocyclitis** | | | | |
| MR Egger (bootstrap) | 77 | 1.022 | [0.97,1.077] | 0.201 |
| Simple median | 77 | 1.036 | [0.999,1.074] | 0.062 |
| Weighted median | 77 | 1.038 | [1.002,1.074] | 0.036* |
| Penalised weighted median | 77 | 1.038 | [1.002,1.075] | 0.042* |
| IVW radial | 77 | 1.034 | [1.012,1.056] | 0.003* |
| Inverse variance weighted (multiplicative random effects) | 77 | 1.034 | [1.012,1.056] | 0.003* |
| Inverse variance weighted (fixed effects) | 77 | 1.034 | [1.008,1.059] | 0.009* |
| Simple mode | 77 | 1.071 | [0.99,1.159] | 0.09 |
| Weighted mode | 77 | 1.071 | [1.003,1.145] | 0.045* |
| Weighted mode (NOME) | 77 | 1.074 | [1.004,1.148] | 0.043* |
| Simple mode (NOME) | 77 | 1.071 | [0.993,1.156] | 0.079 |
| Robust adjusted profile score (RAPS) | 77 | 1.031 | [1.004,1.06] | 0.023* |
| MRPRESSO | 77 | 1.034 | [1.012,1.056] | 0.003* |
| **Early preterm birth->Disorders of optic nerve and visual pathways** | | | | |
| MR Egger (bootstrap) | 77 | 0.893 | [0.817,0.976] | 0.005* |
| Simple median | 77 | 0.962 | [0.905,1.022] | 0.205 |
| Weighted median | 77 | 0.946 | [0.89,1.007] | 0.081 |
| Penalised weighted median | 77 | 0.946 | [0.89,1.007] | 0.081 |
| IVW radial | 77 | 0.954 | [0.916,0.994] | 0.025* |
| Inverse variance weighted (multiplicative random effects) | 77 | 0.954 | [0.916,0.994] | 0.025* |
| Inverse variance weighted (fixed effects) | 77 | 0.954 | [0.917,0.993] | 0.023* |
| Simple mode | 77 | 0.969 | [0.841,1.116] | 0.659 |
| Weighted mode | 77 | 0.948 | [0.855,1.052] | 0.316 |
| Weighted mode (NOME) | 77 | 0.948 | [0.857,1.05] | 0.308 |
| Simple mode (NOME) | 77 | 0.969 | [0.849,1.105] | 0.636 |
| Robust adjusted profile score (RAPS) | 77 | 0.952 | [0.912,0.994] | 0.025* |
| MRPRESSO | 77 | 0.954 | [0.916,0.994] | 0.027* |
| **Early preterm birth->Degeneration of macula and posterior pole** | | | | |
| MR Egger (bootstrap) | 77 | 0.987 | [0.942,1.034] | 0.278 |
| Simple median | 77 | 0.969 | [0.942,0.996] | 0.024* |
| Weighted median | 77 | 0.971 | [0.944,1] | 0.049* |
| Penalised weighted median | 77 | 0.971 | [0.945,0.999] | 0.044* |
| IVW radial | 77 | 0.977 | [0.959,0.996] | 0.014* |
| Inverse variance weighted (multiplicative random effects) | 77 | 0.977 | [0.959,0.996] | 0.014* |
| Inverse variance weighted (fixed effects) | 77 | 0.977 | [0.959,0.996] | 0.016* |
| Simple mode | 77 | 0.962 | [0.9,1.028] | 0.254 |
| Weighted mode | 77 | 0.956 | [0.9,1.015] | 0.15 |
| Weighted mode (NOME) | 77 | 0.953 | [0.899,1.011] | 0.11 |
| Simple mode (NOME) | 77 | 0.962 | [0.897,1.031] | 0.277 |
| Robust adjusted profile score (RAPS) | 77 | 0.977 | [0.957,0.998] | 0.032* |
| MRPRESSO | 77 | 0.977 | [0.959,0.996] | 0.016* |
| **Early preterm birth->Disorders of sclera, cornea, iris and ciliary body** | | | | |
| MR Egger (bootstrap) | 77 | 1.016 | [0.982,1.052] | 0.193 |
| Simple median | 77 | 1.018 | [0.996,1.041] | 0.125 |
| Weighted median | 77 | 1.006 | [0.984,1.029] | 0.588 |
| Penalised weighted median | 77 | 1.006 | [0.983,1.03] | 0.602 |
| IVW radial | 77 | 1.014 | [1.001,1.028] | 0.033* |
| Inverse variance weighted (multiplicative random effects) | 77 | 1.014 | [1.001,1.028] | 0.033* |
| Inverse variance weighted (fixed effects) | 77 | 1.014 | [0.999,1.03] | 0.064 |
| Simple mode | 77 | 0.988 | [0.939,1.04] | 0.636 |
| Weighted mode | 77 | 1.003 | [0.965,1.043] | 0.866 |
| Weighted mode (NOME) | 77 | 1.003 | [0.962,1.045] | 0.875 |
| Simple mode (NOME) | 77 | 0.988 | [0.938,1.041] | 0.641 |
| Robust adjusted profile score (RAPS) | 77 | 1.016 | [0.999,1.033] | 0.067 |
| MRPRESSO | 77 | 1.014 | [1.001,1.028] | 0.036* |
| **Early preterm birth->Other disorders of eyelid** | | | | |
| MR Egger (bootstrap) | 77 | 0.97 | [0.932,1.01] | 0.067 |
| Simple median | 77 | 0.978 | [0.952,1.005] | 0.105 |
| Weighted median | 77 | 0.969 | [0.943,0.995] | 0.02* |
| Penalised weighted median | 77 | 0.969 | [0.944,0.994] | 0.017* |
| IVW radial | 77 | 0.981 | [0.965,0.997] | 0.023* |
| Inverse variance weighted (multiplicative random effects) | 77 | 0.981 | [0.965,0.997] | 0.022* |
| Inverse variance weighted (fixed effects) | 77 | 0.981 | [0.963,0.999] | 0.04* |
| Simple mode | 77 | 0.965 | [0.91,1.022] | 0.229 |
| Weighted mode | 77 | 0.964 | [0.923,1.006] | 0.098 |
| Weighted mode (NOME) | 77 | 0.964 | [0.925,1.004] | 0.081 |
| Simple mode (NOME) | 77 | 0.965 | [0.915,1.017] | 0.188 |
| Robust adjusted profile score (RAPS) | 77 | 0.979 | [0.96,0.999] | 0.04* |
| MRPRESSO | 77 | 0.981 | [0.965,0.997] | 0.025* |
| **Early preterm birth->Other disorders of optic [2nd] nerve and visual pathways** | | | | |
| MR Egger (bootstrap) | 77 | 0.863 | [0.768,0.97] | 0.005* |
| Simple median | 77 | 0.941 | [0.871,1.016] | 0.121 |
| Weighted median | 77 | 0.905 | [0.839,0.976] | 0.01* |
| Penalised weighted median | 77 | 0.902 | [0.837,0.973] | 0.007* |
| IVW radial | 77 | 0.932 | [0.887,0.98] | 0.005* |
| Inverse variance weighted (multiplicative random effects) | 77 | 0.932 | [0.887,0.98] | 0.005* |
| Inverse variance weighted (fixed effects) | 77 | 0.932 | [0.886,0.981] | 0.007* |
| Simple mode | 77 | 1.005 | [0.849,1.189] | 0.958 |
| Weighted mode | 77 | 0.899 | [0.793,1.018] | 0.098 |
| Weighted mode (NOME) | 77 | 0.892 | [0.787,1.012] | 0.079 |
| Simple mode (NOME) | 77 | 1.005 | [0.848,1.191] | 0.958 |
| Robust adjusted profile score (RAPS) | 77 | 0.926 | [0.876,0.979] | 0.007* |
| MRPRESSO | 77 | 0.932 | [0.887,0.98] | 0.007* |
| **Early preterm birth->Acute and subacute iridocyclitis** | | | | |
| MR Egger (bootstrap) | 77 | 1.013 | [0.955,1.074] | 0.338 |
| Simple median | 77 | 1.026 | [0.987,1.067] | 0.194 |
| Weighted median | 77 | 1.025 | [0.986,1.066] | 0.199 |
| Penalised weighted median | 77 | 1.025 | [0.987,1.065] | 0.192 |
| IVW radial | 77 | 1.031 | [1.006,1.058] | 0.016* |
| Inverse variance weighted (multiplicative random effects) | 77 | 1.031 | [1.006,1.058] | 0.016* |
| Inverse variance weighted (fixed effects) | 77 | 1.031 | [1.005,1.059] | 0.02* |
| Simple mode | 77 | 1.03 | [0.944,1.125] | 0.499 |
| Weighted mode | 77 | 1.039 | [0.956,1.128] | 0.372 |
| Weighted mode (NOME) | 77 | 1.048 | [0.966,1.137] | 0.259 |
| Simple mode (NOME) | 77 | 1.03 | [0.938,1.133] | 0.53 |
| Robust adjusted profile score (RAPS) | 77 | 1.03 | [1.001,1.06] | 0.04* |
| MRPRESSO | 77 | 1.031 | [1.006,1.058] | 0.018* |
| **Early preterm birth->Blepharochalasis** | | | | |
| MR Egger (bootstrap) | 77 | 0.958 | [0.91,1.009] | 0.057 |
| Simple median | 77 | 0.977 | [0.944,1.012] | 0.189 |
| Weighted median | 77 | 0.978 | [0.945,1.013] | 0.21 |
| Penalised weighted median | 77 | 0.978 | [0.944,1.014] | 0.216 |
| IVW radial | 77 | 0.974 | [0.954,0.995] | 0.013* |
| Inverse variance weighted (multiplicative random effects) | 77 | 0.974 | [0.954,0.995] | 0.013* |
| Inverse variance weighted (fixed effects) | 77 | 0.974 | [0.952,0.998] | 0.03* |
| Simple mode | 77 | 0.98 | [0.906,1.061] | 0.622 |
| Weighted mode | 77 | 0.98 | [0.922,1.042] | 0.525 |
| Weighted mode (NOME) | 77 | 0.978 | [0.917,1.044] | 0.513 |
| Simple mode (NOME) | 77 | 0.98 | [0.909,1.057] | 0.607 |
| Robust adjusted profile score (RAPS) | 77 | 0.972 | [0.948,0.998] | 0.034* |
| MRPRESSO | 77 | 0.974 | [0.954,0.995] | 0.015* |
| **Early preterm birth->Chronic conjunctivitis** | | | | |
| MR Egger (bootstrap) | 77 | 1.026 | [0.873,1.207] | 0.374 |
| Simple median | 77 | 0.914 | [0.82,1.019] | 0.106 |
| Weighted median | 77 | 0.921 | [0.829,1.023] | 0.127 |
| Penalised weighted median | 77 | 0.921 | [0.827,1.026] | 0.136 |
| IVW radial | 77 | 0.937 | [0.878,1] | 0.048* |
| Inverse variance weighted (multiplicative random effects) | 77 | 0.937 | [0.878,1] | 0.049* |
| Inverse variance weighted (fixed effects) | 77 | 0.937 | [0.873,1.006] | 0.072 |
| Simple mode | 77 | 1.126 | [0.879,1.443] | 0.349 |
| Weighted mode | 77 | 1.113 | [0.908,1.364] | 0.305 |
| Weighted mode (NOME) | 77 | 1.12 | [0.914,1.372] | 0.278 |
| Simple mode (NOME) | 77 | 1.126 | [0.885,1.433] | 0.335 |
| Robust adjusted profile score (RAPS) | 77 | 0.929 | [0.86,1.003] | 0.062 |
| MRPRESSO | 77 | 0.937 | [0.878,1] | 0.053 |

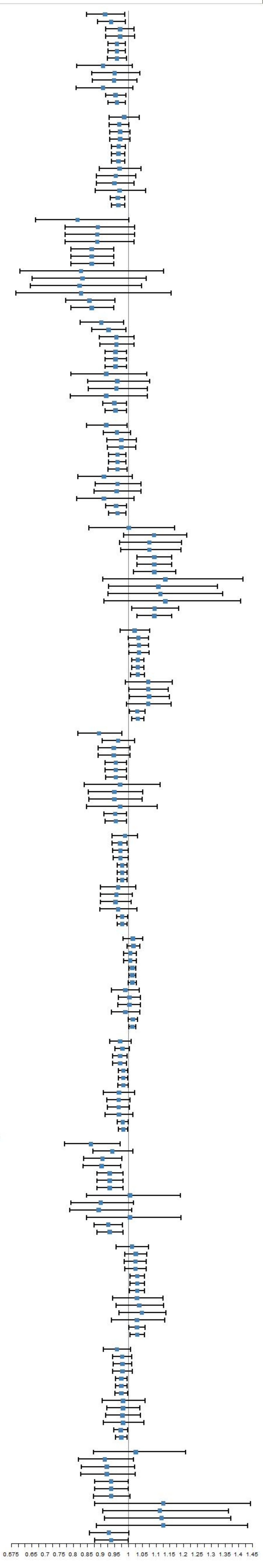

**Fig S1.4 UVMR results of early preterm birth on adult eye diseases.**

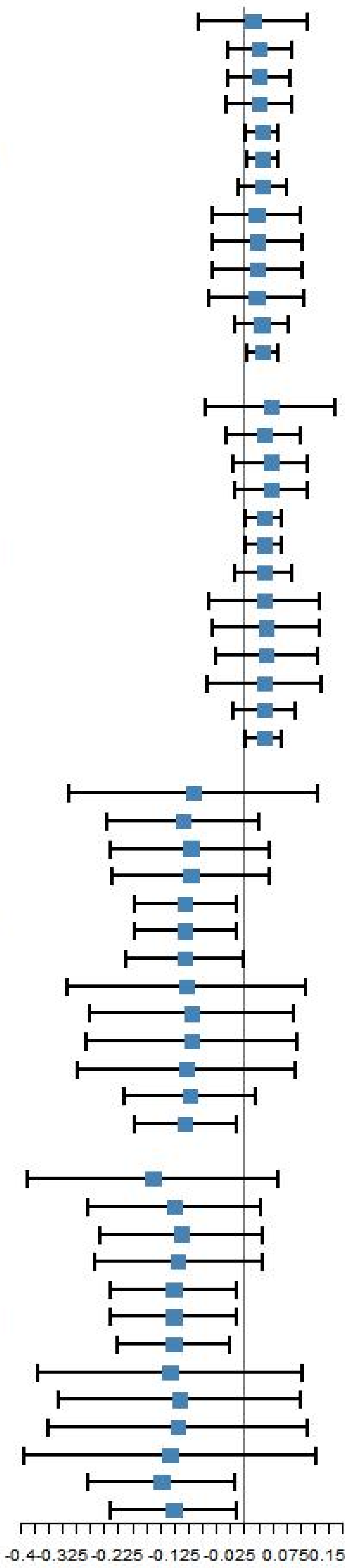

| Methods | nSNPs | BETA | CI | PVAL |
|---|---|---|---|---|
| **Early preterm birth->IL10** | | | | |
| MR Egger (bootstrap) | 12 | 0.015 | [-0.083,0.113] | 0.382 |
| Simple median | 12 | 0.027 | [-0.029,0.084] | 0.342 |
| Weighted median | 12 | 0.027 | [-0.029,0.083] | 0.345 |
| Penalised weighted median | 12 | 0.027 | [-0.032,0.086] | 0.365 |
| IVW radial | 12 | 0.032 | [0.003,0.060] | 0.028* |
| Inverse variance weighted (multiplicative random effects) | 12 | 0.032 | [0.004,0.060] | 0.027* |
| Inverse variance weighted (fixed effects) | 12 | 0.032 | [-0.012,0.075] | 0.154 |
| Simple mode | 12 | 0.022 | [-0.056,0.101] | 0.587 |
| Weighted mode | 12 | 0.023 | [-0.057,0.103] | 0.583 |
| Weighted mode (NOME) | 12 | 0.023 | [-0.057,0.103] | 0.581 |
| Simple mode (NOME) | 12 | 0.022 | [-0.062,0.107] | 0.614 |
| Robust adjusted profile score (RAPS) | 12 | 0.031 | [-0.017,0.079] | 0.206 |
| MRPRESSO | 12 | 0.032 | [0.004,0.060] | 0.05 |
| **Early preterm birth->TNFa** | | | | |
| MR Egger (bootstrap) | 12 | 0.047 | [-0.069,0.162] | 0.22 |
| Simple median | 12 | 0.035 | [-0.031,0.101] | 0.302 |
| Weighted median | 12 | 0.047 | [-0.020,0.114] | 0.169 |
| Penalised weighted median | 12 | 0.047 | [-0.017,0.111] | 0.151 |
| IVW radial | 12 | 0.035 | [0.003,0.067] | 0.033* |
| Inverse variance weighted (multiplicative random effects) | 12 | 0.035 | [0.003,0.067] | 0.032* |
| Inverse variance weighted (fixed effects) | 12 | 0.035 | [-0.016,0.086] | 0.177 |
| Simple mode | 12 | 0.035 | [-0.062,0.133] | 0.489 |
| Weighted mode | 12 | 0.039 | [-0.057,0.135] | 0.448 |
| Weighted mode (NOME) | 12 | 0.039 | [-0.051,0.130] | 0.413 |
| Simple mode (NOME) | 12 | 0.035 | [-0.065,0.136] | 0.503 |
| Robust adjusted profile score (RAPS) | 12 | 0.035 | [-0.021,0.091] | 0.215 |
| MRPRESSO | 12 | 0.035 | [0.003,0.067] | 0.055 |
| **Preterm birth->CTACK** | | | | |
| MR Egger (bootstrap) | 12 | -0.091 | [-0.312,0.131] | 0.205 |
| Simple median | 12 | -0.109 | [-0.245,0.027] | 0.117 |
| Weighted median | 12 | -0.096 | [-0.239,0.046] | 0.186 |
| Penalised weighted median | 12 | -0.096 | [-0.237,0.045] | 0.181 |
| IVW radial | 12 | -0.106 | [-0.197,-0.015] | 0.023* |
| Inverse variance weighted (multiplicative random effects) | 12 | -0.106 | [-0.196,-0.015] | 0.023* |
| Inverse variance weighted (fixed effects) | 12 | -0.106 | [-0.211,-0.000] | 0.05 |
| Simple mode | 12 | -0.103 | [-0.315,0.109] | 0.359 |
| Weighted mode | 12 | -0.095 | [-0.276,0.087] | 0.329 |
| Weighted mode (NOME) | 12 | -0.095 | [-0.282,0.093] | 0.345 |
| Simple mode (NOME) | 12 | -0.103 | [-0.298,0.091] | 0.32 |
| Robust adjusted profile score (RAPS) | 12 | -0.097 | [-0.214,0.019] | 0.101 |
| MRPRESSO | 12 | -0.106 | [-0.196,-0.015] | 0.044* |
| **Preterm birth->GROA** | | | | |
| MR Egger (bootstrap) | 12 | -0.163 | [-0.388,0.061] | 0.077 |
| Simple median | 12 | -0.124 | [-0.278,0.030] | 0.115 |
| Weighted median | 12 | -0.112 | [-0.256,0.032] | 0.126 |
| Penalised weighted median | 12 | -0.118 | [-0.268,0.032] | 0.122 |
| IVW radial | 12 | -0.126 | [-0.239,-0.013] | 0.029* |
| Inverse variance weighted (multiplicative random effects) | 12 | -0.126 | [-0.240,-0.013] | 0.029* |
| Inverse variance weighted (fixed effects) | 12 | -0.126 | [-0.228,-0.025] | 0.015* |
| Simple mode | 12 | -0.132 | [-0.368,0.104] | 0.295 |
| Weighted mode | 12 | -0.115 | [-0.330,0.099] | 0.314 |
| Weighted mode (NOME) | 12 | -0.118 | [-0.349,0.113] | 0.338 |
| Simple mode (NOME) | 12 | -0.132 | [-0.394,0.129] | 0.343 |
| Robust adjusted profile score (RAPS) | 12 | -0.148 | [-0.280,-0.016] | 0.027* |
| MRPRESSO | 12 | -0.126 | [-0.240,-0.013] | 0.052 |

**Fig S2 UVMR results of maternal pregnancy disorders on neonatal cytokine levels.**

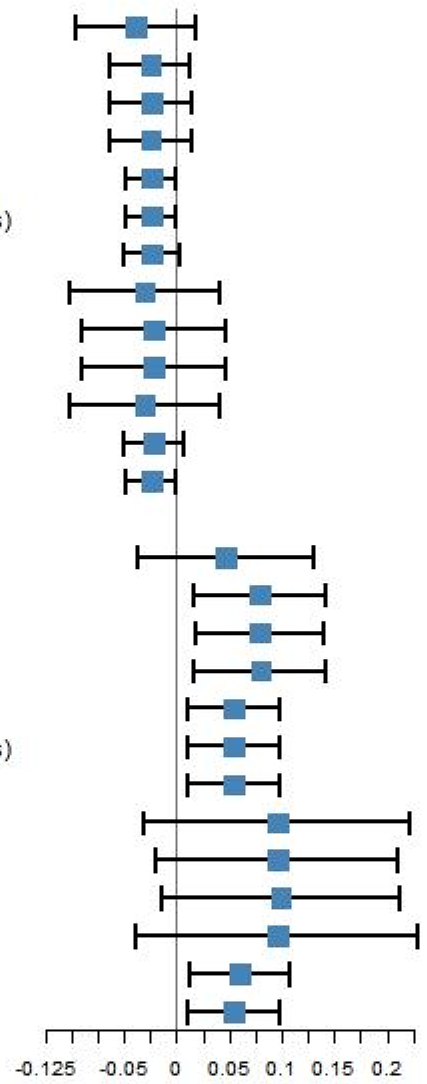

| Methods | nSNPs | BETA | CI | PVAL |
|---|---|---|---|---|
| **Early preterm birth->IP10** | | | | |
| MR Egger (bootstrap) | 69 | -0.039 | [-0.096,0.018] | 0.091 |
| Simple median | 69 | -0.025 | [-0.063,0.013] | 0.203 |
| Weighted median | 69 | -0.024 | [-0.064,0.015] | 0.226 |
| Penalised weighted median | 69 | -0.025 | [-0.063,0.014] | 0.207 |
| IVW radial | 69 | -0.024 | [-0.048,-0.000] | 0.05 |
| Inverse variance weighted (multiplicative random effects) | 69 | -0.024 | [-0.048,-0.000] | 0.049* |
| Inverse variance weighted (fixed effects) | 69 | -0.024 | [-0.051,0.003] | 0.082 |
| Simple mode | 69 | -0.03 | [-0.102,0.042] | 0.42 |
| Weighted mode | 69 | -0.022 | [-0.091,0.047] | 0.537 |
| Weighted mode (NOME) | 69 | -0.022 | [-0.090,0.046] | 0.532 |
| Simple mode (NOME) | 69 | -0.03 | [-0.101,0.041] | 0.413 |
| Robust adjusted profile score (RAPS) | 69 | -0.022 | [-0.051,0.007] | 0.14 |
| MRPRESSO | 69 | -0.024 | [-0.048,-0.000] | 0.054 |
| **Preterm birth->IL12p70** | | | | |
| MR Egger (bootstrap) | 68 | 0.047 | [-0.036,0.130] | 0.146 |
| Simple median | 68 | 0.079 | [0.017,0.142] | 0.013* |
| Weighted median | 68 | 0.079 | [0.019,0.140] | 0.01* |
| Penalised weighted median | 68 | 0.08 | [0.017,0.142] | 0.013* |
| IVW radial | 68 | 0.054 | [0.010,0.098] | 0.017* |
| Inverse variance weighted (multiplicative random effects) | 68 | 0.054 | [0.010,0.098] | 0.017* |
| Inverse variance weighted (fixed effects) | 68 | 0.054 | [0.010,0.098] | 0.015* |
| Simple mode | 68 | 0.096 | [-0.031,0.222] | 0.143 |
| Weighted mode | 68 | 0.096 | [-0.019,0.210] | 0.105 |
| Weighted mode (NOME) | 68 | 0.099 | [-0.014,0.212] | 0.09 |
| Simple mode (NOME) | 68 | 0.096 | [-0.039,0.230] | 0.169 |
| Robust adjusted profile score (RAPS) | 68 | 0.06 | [0.013,0.108] | 0.013* |
| MRPRESSO | 68 | 0.054 | [0.010,0.098] | 0.02* |

**Fig S3 UVMR results of maternal pregnancy disorders on adult cytokine levels**

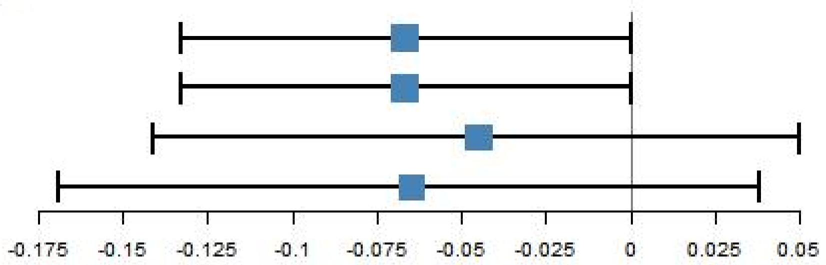

| Methods | nSNPs | BETA | CI | PVAL |
|---|---|---|---|---|
| **Preterm birth->IL6** | | | | |
| mvmr_ivw | 33 | -0.067 | [-0.133,-0.000] | 0.049* |
| mvmr_lasso | 33 | -0.067 | [-0.133,-0.000] | 0.049* |
| mvmr_median | 33 | -0.045 | [-0.141,0.050] | 0.353 |
| mvmr_egger | 33 | -0.065 | [-0.169,0.038] | 0.217 |

**Fig S4 MVMR results of maternal pregnancy disorders on adult cytokine levels adjusting for neonatal levels.**

| Methods | nSNPs | BETA | CI | PVAL |
|---|---|---|---|---|
| **CTACK->Other and unspecified visual disturbances** | | | | |
| MR Egger (bootstrap) | 65 | -0.187 | [-0.429,0.056] | 0.06 |
| Simple median | 65 | -0.18 | [-0.335,-0.025] | 0.023* |
| Weighted median | 65 | -0.112 | [-0.272,0.048] | 0.169 |
| Penalised weighted median | 65 | -0.112 | [-0.276,0.051] | 0.178 |
| IVW radial | 65 | -0.114 | [-0.202,-0.027] | 0.01* |
| Inverse variance weighted (multiplicative random effects) | 65 | -0.114 | [-0.202,-0.027] | 0.01* |
| Inverse variance weighted (fixed effects) | 65 | -0.114 | [-0.218,-0.011] | 0.031* |
| Simple mode | 65 | -0.267 | [-0.623,0.089] | 0.147 |
| Weighted mode | 65 | -0.076 | [-0.344,0.192] | 0.581 |
| Weighted mode (NOME) | 65 | -0.081 | [-0.351,0.189] | 0.557 |
| Simple mode (NOME) | 65 | -0.267 | [-0.625,0.092] | 0.15 |
| Robust adjusted profile score (RAPS) | 65 | -0.122 | [-0.235,-0.009] | 0.035* |
| MRPRESSO | 65 | -0.114 | [-0.202,-0.027] | 0.013* |
| **CTACK->Paralytic strabismus** | | | | |
| MR Egger (bootstrap) | 65 | 0.015 | [-0.238,0.268] | 0.453 |
| Simple median | 65 | -0.104 | [-0.258,0.049] | 0.183 |
| Weighted median | 65 | -0.111 | [-0.271,0.049] | 0.174 |
| Penalised weighted median | 65 | -0.11 | [-0.271,0.050] | 0.177 |
| IVW radial | 65 | -0.106 | [-0.211,-0.002] | 0.046* |
| Inverse variance weighted (multiplicative random effects) | 65 | -0.106 | [-0.210,-0.002] | 0.046* |
| Inverse variance weighted (fixed effects) | 65 | -0.106 | [-0.209,-0.003] | 0.044* |
| Simple mode | 65 | -0.132 | [-0.461,0.196] | 0.433 |
| Weighted mode | 65 | -0.114 | [-0.395,0.166] | 0.427 |
| Weighted mode (NOME) | 65 | -0.12 | [-0.393,0.152] | 0.39 |
| Simple mode (NOME) | 65 | -0.132 | [-0.479,0.214] | 0.457 |
| Robust adjusted profile score (RAPS) | 65 | -0.108 | [-0.219,0.004] | 0.059 |
| MRPRESSO | 65 | -0.106 | [-0.210,-0.002] | 0.05 |
| **GROA->Dacryoadenitis** | | | | |
| MR Egger (bootstrap) | 53 | 0.436 | [-0.128,0.999] | 0.056 |
| Simple median | 53 | 0.55 | [0.099,1.002] | 0.017* |
| Weighted median | 53 | 0.52 | [0.071,0.970] | 0.023* |
| Penalised weighted median | 53 | 0.519 | [0.084,0.955] | 0.019* |
| IVW radial | 53 | 0.466 | [0.166,0.766] | 0.002* |
| Inverse variance weighted (multiplicative random effects) | 53 | 0.466 | [0.166,0.766] | 0.002* |
| Inverse variance weighted (fixed effects) | 53 | 0.466 | [0.169,0.763] | 0.002* |
| Simple mode | 53 | 0.543 | [-0.355,1.440] | 0.241 |
| Weighted mode | 53 | 0.467 | [-0.072,1.007] | 0.096 |
| Weighted mode (NOME) | 53 | 0.486 | [-0.039,1.011] | 0.075 |
| Simple mode (NOME) | 53 | 0.543 | [-0.333,1.419] | 0.23 |
| Robust adjusted profile score (RAPS) | 53 | 0.439 | [0.121,0.757] | 0.007* |
| MRPRESSO | 53 | 0.466 | [0.166,0.766] | 0.004* |
| **GROA->Intermittent heterotropia** | | | | |
| MR Egger (bootstrap) | 53 | -0.144 | [-0.347,0.059] | 0.08 |
| Simple median | 53 | -0.084 | [-0.246,0.078] | 0.312 |
| Weighted median | 53 | -0.153 | [-0.323,0.017] | 0.078 |
| Penalised weighted median | 53 | -0.153 | [-0.319,0.013] | 0.072 |
| IVW radial | 53 | -0.112 | [-0.210,-0.013] | 0.026* |
| Inverse variance weighted (multiplicative random effects) | 53 | -0.112 | [-0.210,-0.013] | 0.026* |
| Inverse variance weighted (fixed effects) | 53 | -0.112 | [-0.217,-0.006] | 0.038* |
| Simple mode | 53 | -0.144 | [-0.449,0.161] | 0.358 |
| Weighted mode | 53 | -0.163 | [-0.347,0.021] | 0.088 |
| Weighted mode (NOME) | 53 | -0.17 | [-0.361,0.022] | 0.089 |
| Simple mode (NOME) | 53 | -0.144 | [-0.454,0.166] | 0.366 |
| Robust adjusted profile score (RAPS) | 53 | -0.108 | [-0.221,0.006] | 0.062 |
| MRPRESSO | 53 | -0.112 | [-0.210,-0.013] | 0.03* |
| **TNFa->Convergent concomitant strabismus** | | | | |
| MR Egger (bootstrap) | 49 | 0.022 | [-0.290,0.335] | 0.464 |
| Simple median | 49 | 0.197 | [0.023,0.371] | 0.027* |
| Weighted median | 49 | 0.194 | [0.026,0.361] | 0.023* |
| Penalised weighted median | 49 | 0.2 | [0.029,0.371] | 0.022* |
| IVW radial | 49 | 0.133 | [0.018,0.248] | 0.023* |
| Inverse variance weighted (multiplicative random effects) | 49 | 0.133 | [0.018,0.248] | 0.023* |
| Inverse variance weighted (fixed effects) | 49 | 0.133 | [0.012,0.254] | 0.032* |
| Simple mode | 49 | 0.245 | [-0.149,0.639] | 0.229 |
| Weighted mode | 49 | 0.235 | [-0.125,0.595] | 0.208 |
| Weighted mode (NOME) | 49 | 0.245 | [-0.156,0.645] | 0.236 |
| Simple mode (NOME) | 49 | 0.245 | [-0.138,0.628] | 0.216 |
| Robust adjusted profile score (RAPS) | 49 | 0.159 | [0.026,0.292] | 0.019* |
| MRPRESSO | 49 | 0.133 | [0.018,0.248] | 0.028* |
| **TNFa->Intermittent heterotropia** | | | | |
| MR Egger (bootstrap) | 49 | 0.072 | [-0.269,0.414] | 0.336 |
| Simple median | 49 | 0.238 | [0.048,0.428] | 0.014* |
| Weighted median | 49 | 0.213 | [0.020,0.405] | 0.03* |
| Penalised weighted median | 49 | 0.227 | [0.034,0.420] | 0.021* |
| IVW radial | 49 | 0.134 | [0.003,0.264] | 0.045* |
| Inverse variance weighted (multiplicative random effects) | 49 | 0.133 | [0.003,0.264] | 0.045* |
| Inverse variance weighted (fixed effects) | 49 | 0.133 | [0.004,0.263] | 0.044* |
| Simple mode | 49 | 0.371 | [-0.076,0.819] | 0.11 |
| Weighted mode | 49 | 0.365 | [-0.067,0.797] | 0.104 |
| Weighted mode (NOME) | 49 | 0.371 | [-0.078,0.821] | 0.112 |
| Simple mode (NOME) | 49 | 0.371 | [-0.127,0.870] | 0.151 |
| Robust adjusted profile score (RAPS) | 49 | 0.146 | [0.000,0.292] | 0.05 |
| MRPRESSO | 49 | 0.133 | [0.003,0.264] | 0.05 |

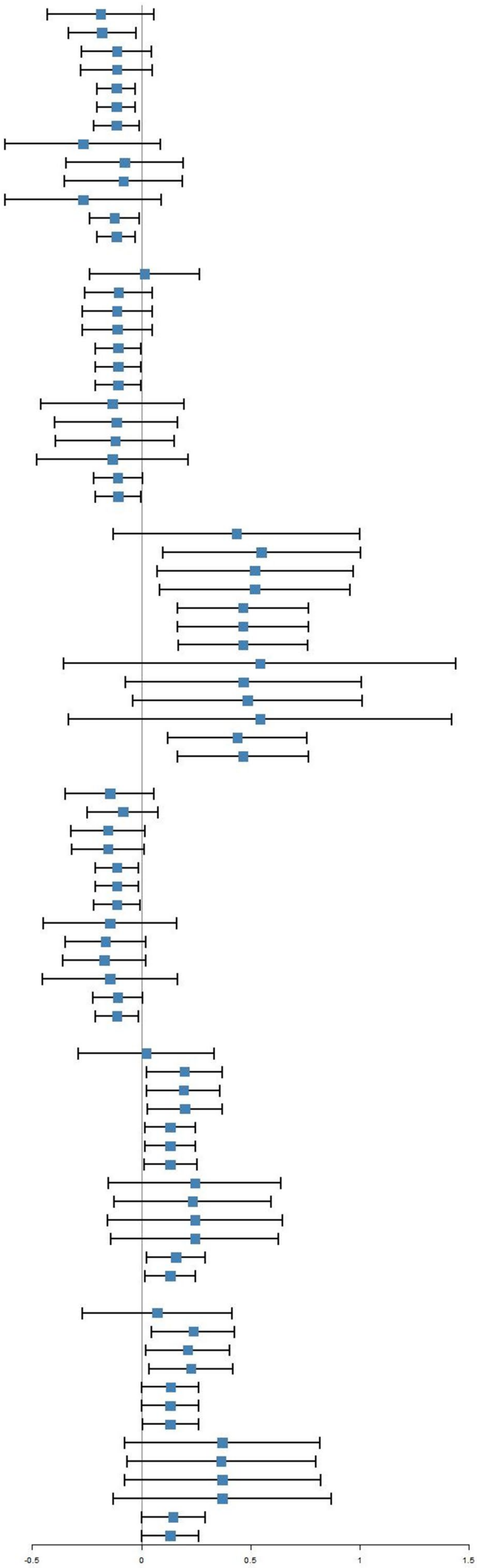

**Fig S5.1 UVMR results of adult cytokine levels (CTACK/GROA/TNF-α) on adult eye diseases.**

| Methods | nSNPs | BETA | CI | PVAL |
|---|---|---|---|---|
| **IL10->Disorders of optic nerve and visual pathways** | | | | |
| MR Egger (bootstrap) | 51 | 0.042 | [-0.245,0.329] | 0.373 |
| Simple median | 51 | 0.143 | [-0.007,0.293] | 0.062 |
| Weighted median | 51 | 0.114 | [-0.039,0.266] | 0.144 |
| Penalised weighted median | 51 | 0.114 | [-0.036,0.265] | 0.135 |
| IVW radial | 51 | 0.123 | [0.003,0.243] | 0.045* |
| Inverse variance weighted (multiplicative random effects) | 51 | 0.123 | [0.003,0.243] | 0.045* |
| Inverse variance weighted (fixed effects) | 51 | 0.123 | [0.021,0.225] | 0.018* |
| Simple mode | 51 | 0.265 | [-0.066,0.595] | 0.123 |
| Weighted mode | 51 | 0.192 | [-0.100,0.485] | 0.204 |
| Weighted mode (NOME) | 51 | 0.212 | [-0.082,0.506] | 0.163 |
| Simple mode (NOME) | 51 | 0.265 | [-0.044,0.573] | 0.099 |
| Robust adjusted profile score (RAPS) | 51 | 0.162 | [0.039,0.285] | 0.01* |
| MRPRESSO | | 0.144 | [0.029,0.258] | 0.017* |
| **IL10->Disorders of vitreous body** | | | | |
| MR Egger (bootstrap) | 51 | 0.019 | [-0.104,0.143] | 0.395 |
| Simple median | 51 | 0.095 | [0.023,0.166] | 0.01* |
| Weighted median | 51 | 0.083 | [0.016,0.149] | 0.015* |
| Penalised weighted median | 51 | 0.087 | [0.017,0.158] | 0.015* |
| IVW radial | 51 | 0.06 | [0.007,0.113] | 0.028* |
| Inverse variance weighted (multiplicative random effects) | 51 | 0.06 | [0.007,0.113] | 0.028* |
| Inverse variance weighted (fixed effects) | 51 | 0.06 | [0.014,0.106] | 0.011* |
| Simple mode | 51 | 0.113 | [-0.037,0.264] | 0.145 |
| Weighted mode | 51 | 0.095 | [-0.040,0.229] | 0.174 |
| Weighted mode (NOME) | 51 | 0.1 | [-0.043,0.243] | 0.176 |
| Simple mode (NOME) | 51 | 0.113 | [-0.052,0.279] | 0.185 |
| Robust adjusted profile score (RAPS) | 51 | 0.073 | [0.014,0.133] | 0.015* |
| MRPRESSO | 51 | 0.06 | [0.007,0.113] | 0.032* |
| **IL10->Disorders of vitreous body and globe** | | | | |
| MR Egger (bootstrap) | 51 | 0.023 | [-0.097,0.143] | 0.353 |
| Simple median | 51 | 0.084 | [0.019,0.149] | 0.012* |
| Weighted median | 51 | 0.075 | [0.012,0.139] | 0.019* |
| Penalised weighted median | 51 | 0.079 | [0.012,0.146] | 0.022* |
| IVW radial | 51 | 0.061 | [0.009,0.113] | 0.022* |
| Inverse variance weighted (multiplicative random effects) | 51 | 0.061 | [0.009,0.113] | 0.022* |
| Inverse variance weighted (fixed effects) | 51 | 0.061 | [0.016,0.106] | 0.008* |
| Simple mode | 51 | 0.089 | [-0.055,0.233] | 0.233 |
| Weighted mode | 51 | 0.079 | [-0.053,0.211] | 0.246 |
| Weighted mode (NOME) | 51 | 0.079 | [-0.056,0.214] | 0.256 |
| Simple mode (NOME) | 51 | 0.089 | [-0.053,0.231] | 0.226 |
| Robust adjusted profile score (RAPS) | 51 | 0.072 | [0.014,0.130] | 0.016* |
| MRPRESSO | 51 | 0.061 | [0.009,0.113] | 0.026* |
| **IL10->Other and unspecified disorders of vitreous body** | | | | |
| MR Egger (bootstrap) | 51 | 0.018 | [-0.119,0.155] | 0.393 |
| Simple median | 51 | 0.111 | [0.035,0.187] | 0.004* |
| Weighted median | 51 | 0.102 | [0.023,0.181] | 0.012* |
| Penalised weighted median | 51 | 0.102 | [0.027,0.177] | 0.008* |
| IVW radial | 51 | 0.088 | [0.036,0.139] | 0.001* |
| Inverse variance weighted (multiplicative random effects) | 51 | 0.088 | [0.036,0.139] | 0.001* |
| Inverse variance weighted (fixed effects) | 51 | 0.088 | [0.036,0.140] | 0.001* |
| Simple mode | 51 | 0.164 | [-0.019,0.346] | 0.085 |
| Weighted mode | 51 | 0.143 | [-0.019,0.306] | 0.089 |
| Weighted mode (NOME) | 51 | 0.149 | [-0.005,0.302] | 0.064 |
| Simple mode (NOME) | 51 | 0.164 | [-0.012,0.339] | 0.073 |
| Robust adjusted profile score (RAPS) | 51 | 0.092 | [0.035,0.149] | 0.001* |
| MRPRESSO | 51 | 0.088 | [0.036,0.139] | 0.002* |
| **IL10->Normotensive glaucoma** | | | | |
| MR Egger (bootstrap) | 51 | 0.341 | [-0.034,0.716] | 0.034* |
| Simple median | 51 | 0.248 | [0.057,0.439] | 0.011* |
| Weighted median | 51 | 0.239 | [0.047,0.432] | 0.015* |
| Penalised weighted median | 51 | 0.237 | [0.042,0.432] | 0.017* |
| IVW radial | 51 | 0.201 | [0.046,0.356] | 0.011* |
| Inverse variance weighted (multiplicative random effects) | 51 | 0.201 | [0.046,0.356] | 0.011* |
| Inverse variance weighted (fixed effects) | 51 | 0.201 | [0.076,0.326] | 0.002* |
| Simple mode | 51 | 0.306 | [-0.173,0.784] | 0.217 |
| Weighted mode | 51 | 0.298 | [-0.136,0.732] | 0.185 |
| Weighted mode (NOME) | 51 | 0.306 | [-0.167,0.778] | 0.211 |
| Simple mode (NOME) | 51 | 0.306 | [-0.146,0.757] | 0.191 |
| Robust adjusted profile score (RAPS) | 51 | 0.172 | [0.005,0.340] | 0.044* |
| MRPRESSO | 51 | 0.201 | [0.046,0.356] | 0.014* |
| **IL12p70->Corneal degeneration** | | | | |
| MR Egger (bootstrap) | 52 | -0.368 | [-1.356,0.620] | 0.225 |
| Simple median | 52 | -0.443 | [-1.158,0.273] | 0.226 |
| Weighted median | 52 | -0.732 | [-1.521,0.056] | 0.069 |
| Penalised weighted median | 52 | -0.732 | [-1.498,0.034] | 0.061 |
| IVW radial | 52 | -0.526 | [-0.971,-0.080] | 0.021* |
| Inverse variance weighted (multiplicative random effects) | 52 | -0.527 | [-0.972,-0.082] | 0.02* |
| Inverse variance weighted (fixed effects) | 52 | -0.527 | [-1.006,-0.049] | 0.031* |
| Simple mode | 52 | -0.767 | [-2.261,0.727] | 0.319 |
| Weighted mode | 52 | -0.851 | [-1.857,0.154] | 0.103 |
| Weighted mode (NOME) | 52 | -0.872 | [-1.982,0.237] | 0.129 |
| Simple mode (NOME) | 52 | -0.767 | [-2.329,0.796] | 0.341 |
| Robust adjusted profile score (RAPS) | 52 | -0.59 | [-1.130,-0.049] | 0.032* |
| MRPRESSO | 52 | -0.527 | [-0.972,-0.082] | 0.024* |
| **IL6->Disorders of orbit** | | | | |
| MR Egger (bootstrap) | 52 | -0.261 | [-0.800,0.278] | 0.166 |
| Simple median | 52 | -0.336 | [-0.629,-0.042] | 0.025* |
| Weighted median | 52 | -0.28 | [-0.586,0.027] | 0.073 |
| Penalised weighted median | 52 | -0.285 | [-0.591,0.020] | 0.067 |
| IVW radial | 52 | -0.209 | [-0.409,-0.009] | 0.041* |
| Inverse variance weighted (multiplicative random effects) | 52 | -0.209 | [-0.409,-0.009] | 0.041* |
| Inverse variance weighted (fixed effects) | 52 | -0.209 | [-0.421,0.003] | 0.053 |
| Simple mode | 52 | -0.547 | [-1.243,0.149] | 0.13 |
| Weighted mode | 52 | -0.465 | [-1.043,0.114] | 0.122 |
| Weighted mode (NOME) | 52 | -0.476 | [-1.087,0.134] | 0.133 |
| Simple mode (NOME) | 52 | -0.547 | [-1.188,0.094] | 0.101 |
| Robust adjusted profile score (RAPS) | 52 | -0.238 | [-0.470,-0.006] | 0.044* |
| MRPRESSO | 52 | -0.209 | [-0.409,-0.009] | 0.046* |
| **IL6->Intermittent heterotropia** | | | | |
| MR Egger (bootstrap) | 52 | -0.234 | [-0.684,0.216] | 0.143 |
| Simple median | 52 | -0.33 | [-0.595,-0.064] | 0.015* |
| Weighted median | 52 | -0.291 | [-0.567,-0.016] | 0.038* |
| Penalised weighted median | 52 | -0.292 | [-0.577,-0.008] | 0.044* |
| IVW radial | 52 | -0.231 | [-0.431,-0.032] | 0.023* |
| Inverse variance weighted (multiplicative random effects) | 52 | -0.232 | [-0.431,-0.032] | 0.023* |
| Inverse variance weighted (fixed effects) | 52 | -0.232 | [-0.417,-0.046] | 0.014* |
| Simple mode | 52 | -0.426 | [-1.067,0.215] | 0.199 |
| Weighted mode | 52 | -0.393 | [-0.998,0.212] | 0.208 |
| Weighted mode (NOME) | 52 | -0.404 | [-1.064,0.256] | 0.236 |
| Simple mode (NOME) | 52 | -0.426 | [-1.060,0.208] | 0.194 |
| Robust adjusted profile score (RAPS) | 52 | -0.227 | [-0.442,-0.011] | 0.039* |
| MRPRESSO | 52 | -0.232 | [-0.431,-0.032] | 0.027* |

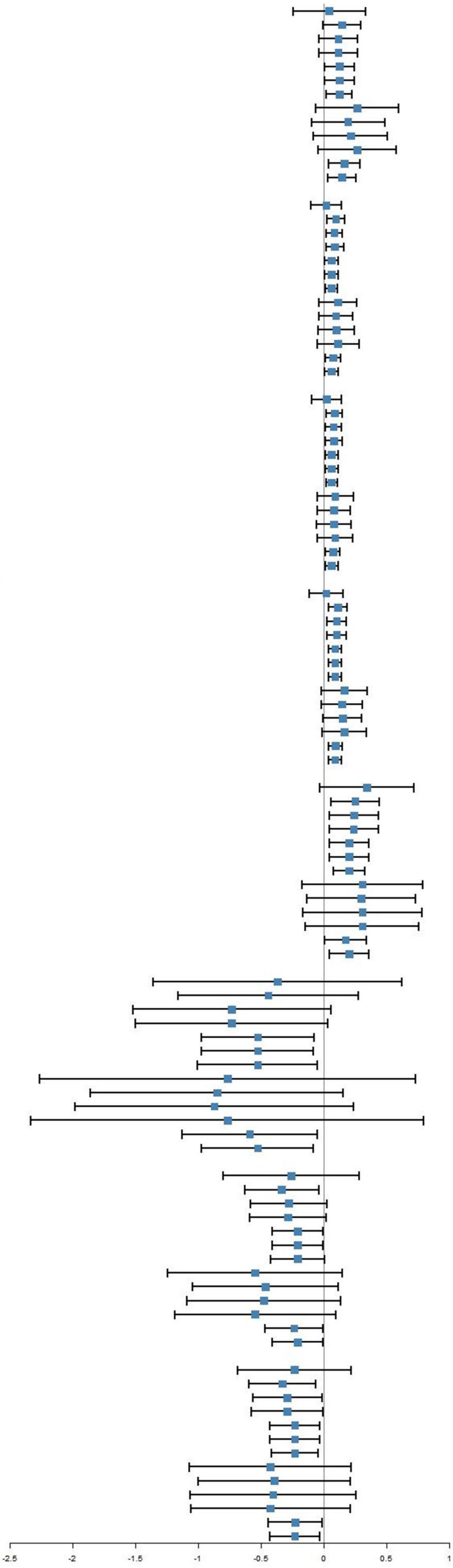

**Fig S5.2 UVMR results of adult cytokine levels (IL10/IL12p70/IL6) on adult eye diseases.**

| Methods | nSNPs | BETA | CI | PVAL |
|---|---|---|---|---|
| **CTACK->Age-related macular degeneration (whether dry or wet)** | | | | |
| MR Egger (bootstrap) | 16 | 0.2 | [-0.148,0.548] | 0.132 |
| Simple median | 16 | 0.295 | [0.088,0.503] | 0.005* |
| Weighted median | 16 | 0.301 | [0.095,0.508] | 0.004* |
| Penalised weighted median | 16 | 0.301 | [0.093,0.510] | 0.005* |
| IVW radial | 16 | 0.317 | [0.220,0.414] | 0* |
| Inverse variance weighted (multiplicative random effects) | 16 | 0.317 | [0.221,0.414] | 0* |
| Inverse variance weighted (fixed effects) | 16 | 0.317 | [0.167,0.467] | 0* |
| Simple mode | 16 | 0.285 | [-0.048,0.618] | 0.114 |
| Weighted mode | 16 | 0.29 | [-0.005,0.584] | 0.073 |
| Weighted mode (NOME) | 16 | 0.292 | [-0.014,0.599] | 0.081 |
| Simple mode (NOME) | 16 | 0.285 | [-0.048,0.618] | 0.114 |
| Robust adjusted profile score (RAPS) | 16 | 0.324 | [0.152,0.495] | 0* |
| MRPRESSO | 16 | 0.317 | [0.221,0.414] | 0* |
| **CTACK->Papilloedema, unspecified** | | | | |
| MR Egger (bootstrap) | 16 | 0.836 | [-0.355,2.027] | 0.085 |
| Simple median | 16 | 0.467 | [-0.302,1.237] | 0.234 |
| Weighted median | 16 | 0.485 | [-0.273,1.242] | 0.21 |
| Penalised weighted median | 16 | 0.485 | [-0.250,1.220] | 0.196 |
| IVW radial | 16 | 0.415 | [0.088,0.742] | 0.013* |
| Inverse variance weighted (multiplicative random effects) | 16 | 0.415 | [0.088,0.742] | 0.013* |
| Inverse variance weighted (fixed effects) | 16 | 0.415 | [-0.141,0.971] | 0.144 |
| Simple mode | 16 | 0.717 | [-0.453,1.888] | 0.248 |
| Weighted mode | 16 | 0.68 | [-0.366,1.726] | 0.222 |
| Weighted mode (NOME) | 16 | 0.689 | [-0.371,1.749] | 0.222 |
| Simple mode (NOME) | 16 | 0.717 | [-0.374,1.808] | 0.217 |
| Robust adjusted profile score (RAPS) | 16 | 0.426 | [-0.193,1.046] | 0.177 |
| MRPRESSO | 16 | 0.415 | [0.088,0.742] | 0.025* |
| **CTACK->Ectropion of eyelid** | | | | |
| MR Egger (bootstrap) | 16 | -0.121 | [-1.071,0.829] | 0.393 |
| Simple median | 16 | -0.333 | [-0.917,0.251] | 0.264 |
| Weighted median | 16 | -0.231 | [-0.799,0.336] | 0.425 |
| Penalised weighted median | 16 | -0.231 | [-0.774,0.311] | 0.404 |
| IVW radial | 16 | -0.402 | [-0.770,-0.034] | 0.032* |
| Inverse variance weighted (multiplicative random effects) | 16 | -0.401 | [-0.769,-0.034] | 0.032* |
| Inverse variance weighted (fixed effects) | 16 | -0.401 | [-0.809,0.006] | 0.053 |
| Simple mode | 16 | -0.221 | [-1.132,0.691] | 0.641 |
| Weighted mode | 16 | -0.186 | [-0.951,0.579] | 0.641 |
| Weighted mode (NOME) | 16 | -0.198 | [-1.044,0.649] | 0.654 |
| Simple mode (NOME) | 16 | -0.221 | [-1.161,0.719] | 0.652 |
| Robust adjusted profile score (RAPS) | 16 | -0.421 | [-0.874,0.033] | 0.069 |
| MRPRESSO | 16 | -0.401 | [-0.769,-0.034] | 0.049* |
| **CTACK->Other and unspecified disorders of vitreous body** | | | | |
| MR Egger (bootstrap) | 16 | 0.158 | [-0.108,0.424] | 0.118 |
| Simple median | 16 | 0.12 | [-0.044,0.284] | 0.153 |
| Weighted median | 16 | 0.152 | [-0.003,0.308] | 0.055 |
| Penalised weighted median | 16 | 0.153 | [-0.007,0.312] | 0.06 |
| IVW radial | 16 | 0.118 | [0.023,0.213] | 0.015* |
| Inverse variance weighted (multiplicative random effects) | 16 | 0.119 | [0.024,0.213] | 0.014* |
| Inverse variance weighted (fixed effects) | 16 | 0.119 | [-0.003,0.240] | 0.055 |
| Simple mode | 16 | 0.117 | [-0.135,0.369] | 0.376 |
| Weighted mode | 16 | 0.18 | [-0.043,0.404] | 0.135 |
| Weighted mode (NOME) | 16 | 0.187 | [-0.027,0.401] | 0.108 |
| Simple mode (NOME) | 16 | 0.117 | [-0.106,0.341] | 0.32 |
| Robust adjusted profile score (RAPS) | 16 | 0.129 | [-0.006,0.264] | 0.061 |
| MRPRESSO | 16 | 0.119 | [0.024,0.213] | 0.027* |
| **CTACK->Optic neuritis** | | | | |
| MR Egger (bootstrap) | 16 | 0.163 | [-0.648,0.974] | 0.345 |
| Simple median | 16 | 0.218 | [-0.286,0.721] | 0.398 |
| Weighted median | 16 | 0.121 | [-0.368,0.610] | 0.628 |
| Penalised weighted median | 16 | 0.121 | [-0.357,0.599] | 0.62 |
| IVW radial | 16 | 0.228 | [0.013,0.443] | 0.038* |
| Inverse variance weighted (multiplicative random effects) | 16 | 0.228 | [0.013,0.443] | 0.038* |
| Inverse variance weighted (fixed effects) | 16 | 0.228 | [-0.141,0.597] | 0.227 |
| Simple mode | 16 | 0.149 | [-0.626,0.923] | 0.712 |
| Weighted mode | 16 | 0.089 | [-0.635,0.812] | 0.814 |
| Weighted mode (NOME) | 16 | 0.094 | [-0.611,0.800] | 0.797 |
| Simple mode (NOME) | 16 | 0.149 | [-0.616,0.914] | 0.709 |
| Robust adjusted profile score (RAPS) | 16 | 0.232 | [-0.178,0.642] | 0.268 |
| MRPRESSO | 16 | 0.228 | [0.013,0.443] | 0.056 |
| **CTACK->Diabetic retinopathy (H7_RETINOPATHYDIAB)** | | | | |
| MR Egger (bootstrap) | 16 | -0.039 | [-0.389,0.310] | 0.406 |
| Simple median | 16 | -0.146 | [-0.345,0.053] | 0.15 |
| Weighted median | 16 | -0.127 | [-0.326,0.073] | 0.214 |
| Penalised weighted median | 16 | -0.127 | [-0.326,0.073] | 0.213 |
| IVW radial | 16 | -0.125 | [-0.225,-0.024] | 0.015* |
| Inverse variance weighted (multiplicative random effects) | 16 | -0.125 | [-0.225,-0.024] | 0.015* |
| Inverse variance weighted (fixed effects) | 16 | -0.125 | [-0.277,0.028] | 0.108 |
| Simple mode | 16 | -0.152 | [-0.478,0.174] | 0.375 |
| Weighted mode | 16 | -0.144 | [-0.453,0.165] | 0.375 |
| Weighted mode (NOME) | 16 | -0.16 | [-0.465,0.146] | 0.322 |
| Simple mode (NOME) | 16 | -0.152 | [-0.488,0.184] | 0.39 |
| Robust adjusted profile score (RAPS) | 16 | -0.128 | [-0.297,0.042] | 0.14 |
| MRPRESSO | 16 | -0.125 | [-0.225,-0.024] | 0.028* |
| **GROA->Other and unspecified iridocyclitis** | | | | |
| MR Egger (bootstrap) | 7 | 0.245 | [-1.260,1.749] | 0.348 |
| Simple median | 7 | 0.646 | [-0.192,1.484] | 0.131 |
| Weighted median | 7 | 0.476 | [-0.328,1.280] | 0.246 |
| Penalised weighted median | 7 | 0.476 | [-0.374,1.326] | 0.272 |
| IVW radial | 7 | 0.565 | [0.246,0.883] | 0.001* |
| Inverse variance weighted (multiplicative random effects) | 7 | 0.564 | [0.246,0.883] | 0.001* |
| Inverse variance weighted (fixed effects) | 7 | 0.564 | [-0.086,1.214] | 0.089 |
| Simple mode | 7 | 0.881 | [-0.311,2.072] | 0.198 |
| Weighted mode | 7 | 0.256 | [-0.912,1.424] | 0.682 |
| Weighted mode (NOME) | 7 | 0.261 | [-0.867,1.390] | 0.666 |
| Simple mode (NOME) | 7 | 0.881 | [-0.245,2.006] | 0.176 |
| Robust adjusted profile score (RAPS) | 7 | 0.571 | [-0.157,1.299] | 0.124 |
| MRPRESSO | 7 | 0.564 | [0.246,0.883] | 0.013* |
| **GROA->Central retinal artery occlusion** | | | | |
| MR Egger (bootstrap) | 9 | -0.432 | [-2.145,1.281] | 0.325 |
| Simple median | 9 | 0.786 | [-0.187,1.759] | 0.113 |
| Weighted median | 9 | 0.721 | [-0.187,1.629] | 0.12 |
| Penalised weighted median | 9 | 0.721 | [-0.200,1.642] | 0.125 |
| IVW radial | 9 | 0.528 | [0.193,0.863] | 0.002* |
| Inverse variance weighted (multiplicative random effects) | 9 | 0.529 | [0.194,0.864] | 0.002* |
| Inverse variance weighted (fixed effects) | 9 | 0.529 | [-0.188,1.246] | 0.148 |
| Simple mode | 9 | 0.93 | [-0.343,2.203] | 0.19 |
| Weighted mode | 9 | 0.894 | [-0.312,2.099] | 0.184 |
| Weighted mode (NOME) | 9 | 0.894 | [-0.396,2.183] | 0.211 |
| Simple mode (NOME) | 9 | 0.93 | [-0.401,2.261] | 0.208 |
| Robust adjusted profile score (RAPS) | 9 | 0.534 | [-0.265,1.333] | 0.19 |
| MRPRESSO | 9 | 0.529 | [0.194,0.864] | 0.015* |
| **GROA->Visual field defects** | | | | |
| MR Egger (bootstrap) | 11 | -0.521 | [-1.220,0.178] | 0.071 |
| Simple median | 11 | -0.359 | [-0.816,0.097] | 0.123 |
| Weighted median | 11 | -0.265 | [-0.698,0.167] | 0.229 |
| Penalised weighted median | 11 | -0.265 | [-0.679,0.148] | 0.208 |
| IVW radial | 11 | -0.304 | [-0.545,-0.064] | 0.013* |
| Inverse variance weighted (multiplicative random effects) | 11 | -0.303 | [-0.543,-0.063] | 0.013* |
| Inverse variance weighted (fixed effects) | 11 | -0.303 | [-0.613,0.008] | 0.056 |
| Simple mode | 11 | -0.295 | [-0.959,0.368] | 0.403 |
| Weighted mode | 11 | -0.17 | [-0.780,0.441] | 0.598 |
| Weighted mode (NOME) | 11 | -0.176 | [-0.760,0.409] | 0.569 |
| Simple mode (NOME) | 11 | -0.295 | [-0.965,0.374] | 0.408 |
| Robust adjusted profile score (RAPS) | 11 | -0.313 | [-0.659,0.033] | 0.077 |
| MRPRESSO | 11 | -0.303 | [-0.543,-0.063] | 0.033* |
| **GROA->Disorders of vitreous body and globe** | | | | |
| MR Egger (bootstrap) | 11 | 0.171 | [-0.154,0.495] | 0.155 |
| Simple median | 11 | 0.117 | [-0.071,0.305] | 0.224 |
| Weighted median | 11 | 0.143 | [-0.034,0.319] | 0.113 |
| Penalised weighted median | 11 | 0.159 | [-0.009,0.328] | 0.064 |
| IVW radial | 11 | 0.134 | [-0.001,0.268] | 0.052 |
| Inverse variance weighted (multiplicative random effects) | 11 | 0.135 | [0.001,0.269] | 0.049* |
| Inverse variance weighted (fixed effects) | 11 | 0.135 | [0.004,0.266] | 0.044* |
| Simple mode | 11 | 0.191 | [-0.074,0.456] | 0.188 |
| Weighted mode | 11 | 0.214 | [-0.011,0.439] | 0.092 |
| Weighted mode (NOME) | 11 | 0.224 | [-0.008,0.457] | 0.088 |
| Simple mode (NOME) | 11 | 0.191 | [-0.054,0.436] | 0.158 |
| Robust adjusted profile score (RAPS) | 11 | 0.157 | [0.013,0.301] | 0.033* |
| MRPRESSO | 11 | 0.135 | [0.001,0.269] | 0.077 |

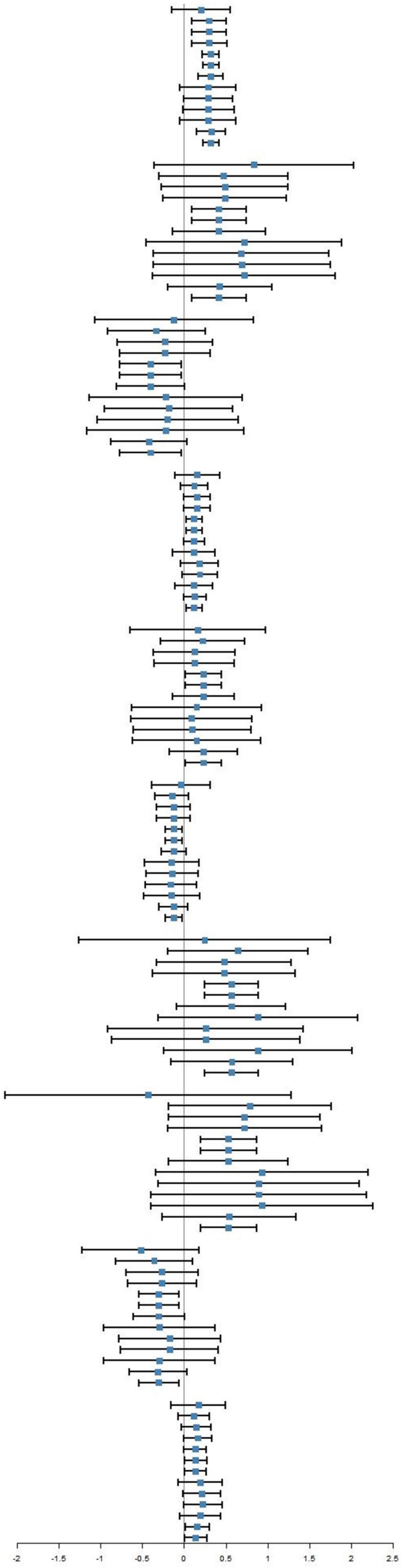

**Fig S6.1 UVMR results of neonatal cytokine levels (CTACK/GROA) on adult eye diseases.**

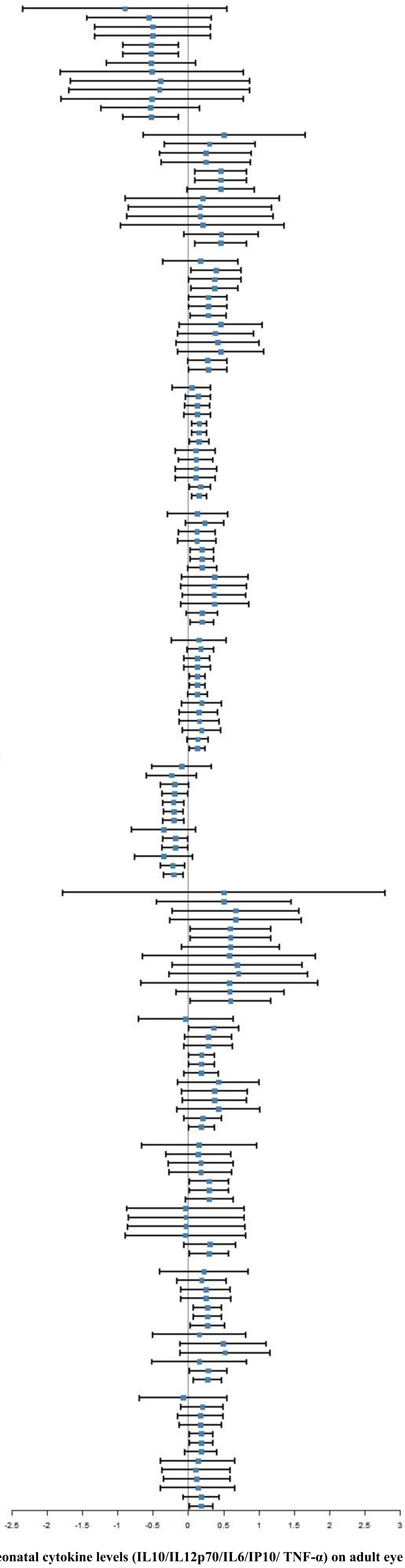

| Methods | nSNPs | BETA | CI | PVAL |
|---|---|---|---|---|
| **IL10->Central retinal artery occlusion** | | | | |
| MR Egger (bootstrap) | 17 | -0.895 | [-2.337,0.547] | 0.118 |
| Simple median | 17 | -0.555 | [-1.436,0.326] | 0.217 |
| Weighted median | 17 | -0.503 | [-1.318,0.313] | 0.227 |
| Penalised weighted median | 17 | -0.503 | [-1.322,0.317] | 0.229 |
| IVW radial | 17 | -0.529 | [-0.923,-0.136] | 0.008* |
| Inverse variance weighted (multiplicative random effects) | 17 | -0.529 | [-0.922,-0.136] | 0.008* |
| Inverse variance weighted (fixed effects) | 17 | -0.529 | [-1.160,0.102] | 0.1 |
| Simple mode | 17 | -0.51 | [-1.806,0.785] | 0.451 |
| Weighted mode | 17 | -0.395 | [-1.661,0.872] | 0.55 |
| Weighted mode (NOME) | 17 | -0.408 | [-1.686,0.871] | 0.541 |
| Simple mode (NOME) | 17 | -0.51 | [-1.803,0.782] | 0.45 |
| Robust adjusted profile score (RAPS) | 17 | -0.538 | [-1.236,0.161] | 0.131 |
| MRPRESSO | 17 | -0.529 | [-0.922,-0.136] | 0.018* |
| **IL10->Other specified and unspecified retinal disorders** | | | | |
| MR Egger (bootstrap) | 18 | 0.509 | [-0.635,1.653] | 0.181 |
| Simple median | 18 | 0.3 | [-0.342,0.943] | 0.359 |
| Weighted median | 18 | 0.248 | [-0.399,0.894] | 0.453 |
| Penalised weighted median | 18 | 0.248 | [-0.385,0.880] | 0.443 |
| IVW radial | 18 | 0.458 | [0.091,0.824] | 0.014* |
| Inverse variance weighted (multiplicative random effects) | 18 | 0.457 | [0.092,0.823] | 0.014* |
| Inverse variance weighted (fixed effects) | 18 | 0.457 | [-0.018,0.933] | 0.06 |
| Simple mode | 18 | 0.203 | [-0.887,1.292] | 0.72 |
| Weighted mode | 18 | 0.167 | [-0.845,1.180] | 0.75 |
| Weighted mode (NOME) | 18 | 0.167 | [-0.869,1.203] | 0.755 |
| Simple mode (NOME) | 18 | 0.203 | [-0.952,1.358] | 0.735 |
| Robust adjusted profile score (RAPS) | 18 | 0.467 | [-0.057,0.992] | 0.081 |
| MRPRESSO | 18 | 0.457 | [0.092,0.823] | 0.025* |
| **IL12p70->Retinal vascular occlusion** | | | | |
| MR Egger (bootstrap) | 16 | 0.174 | [-0.358,0.706] | 0.27 |
| Simple median | 16 | 0.394 | [0.045,0.743] | 0.027* |
| Weighted median | 16 | 0.375 | [0.007,0.744] | 0.046* |
| Penalised weighted median | 16 | 0.375 | [0.043,0.708] | 0.027* |
| IVW radial | 16 | 0.28 | [0.011,0.550] | 0.041* |
| Inverse variance weighted (multiplicative random effects) | 16 | 0.28 | [0.012,0.549] | 0.041* |
| Inverse variance weighted (fixed effects) | 16 | 0.28 | [0.026,0.534] | 0.031* |
| Simple mode | 16 | 0.46 | [-0.124,1.043] | 0.143 |
| Weighted mode | 16 | 0.387 | [-0.154,0.929] | 0.181 |
| Weighted mode (NOME) | 16 | 0.415 | [-0.169,0.998] | 0.184 |
| Simple mode (NOME) | 16 | 0.46 | [-0.150,1.069] | 0.16 |
| Robust adjusted profile score (RAPS) | 16 | 0.273 | [-0.005,0.552] | 0.055 |
| MRPRESSO | 16 | 0.28 | [0.012,0.549] | 0.059 |
| **IL12p70->Degeneration of macula and posterior pole** | | | | |
| MR Egger (bootstrap) | 16 | 0.047 | [-0.228,0.321] | 0.366 |
| Simple median | 16 | 0.137 | [-0.043,0.317] | 0.135 |
| Weighted median | 16 | 0.127 | [-0.049,0.303] | 0.159 |
| Penalised weighted median | 16 | 0.127 | [-0.062,0.315] | 0.187 |
| IVW radial | 16 | 0.155 | [0.052,0.257] | 0.003* |
| Inverse variance weighted (multiplicative random effects) | 16 | 0.154 | [0.052,0.257] | 0.003* |
| Inverse variance weighted (fixed effects) | 16 | 0.154 | [0.019,0.290] | 0.026* |
| Simple mode | 16 | 0.103 | [-0.182,0.387] | 0.489 |
| Weighted mode | 16 | 0.111 | [-0.133,0.355] | 0.386 |
| Weighted mode (NOME) | 16 | 0.111 | [-0.180,0.403] | 0.467 |
| Simple mode (NOME) | 16 | 0.103 | [-0.178,0.384] | 0.484 |
| Robust adjusted profile score (RAPS) | 16 | 0.169 | [0.017,0.321] | 0.029* |
| MRPRESSO | 16 | 0.154 | [0.052,0.257] | 0.01* |
| **IL12p70->Acute and subacute iridocyclitis** | | | | |
| MR Egger (bootstrap) | 14 | 0.13 | [-0.297,0.556] | 0.27 |
| Simple median | 14 | 0.234 | [-0.039,0.507] | 0.092 |
| Weighted median | 14 | 0.124 | [-0.138,0.386] | 0.354 |
| Penalised weighted median | 14 | 0.124 | [-0.148,0.396] | 0.371 |
| IVW radial | 14 | 0.197 | [0.031,0.362] | 0.02* |
| Inverse variance weighted (multiplicative random effects) | 14 | 0.196 | [0.031,0.362] | 0.02* |
| Inverse variance weighted (fixed effects) | 14 | 0.196 | [-0.008,0.401] | 0.06 |
| Simple mode | 14 | 0.374 | [-0.098,0.847] | 0.144 |
| Weighted mode | 14 | 0.362 | [-0.105,0.829] | 0.153 |
| Weighted mode (NOME) | 14 | 0.366 | [-0.078,0.810] | 0.13 |
| Simple mode (NOME) | 14 | 0.374 | [-0.109,0.858] | 0.153 |
| Robust adjusted profile score (RAPS) | 14 | 0.195 | [-0.030,0.421] | 0.09 |
| MRPRESSO | 14 | 0.196 | [0.031,0.362] | 0.037* |
| **IL6->Strabismus** | | | | |
| MR Egger (bootstrap) | 11 | 0.151 | [-0.237,0.540] | 0.217 |
| Simple median | 11 | 0.177 | [-0.013,0.366] | 0.068 |
| Weighted median | 11 | 0.126 | [-0.058,0.311] | 0.18 |
| Penalised weighted median | 11 | 0.126 | [-0.059,0.312] | 0.182 |
| IVW radial | 11 | 0.131 | [0.022,0.240] | 0.019* |
| Inverse variance weighted (multiplicative random effects) | 11 | 0.131 | [0.022,0.240] | 0.019* |
| Inverse variance weighted (fixed effects) | 11 | 0.131 | [-0.005,0.267] | 0.06 |
| Simple mode | 11 | 0.188 | [-0.098,0.474] | 0.226 |
| Weighted mode | 11 | 0.148 | [-0.123,0.418] | 0.31 |
| Weighted mode (NOME) | 11 | 0.155 | [-0.131,0.442] | 0.313 |
| Simple mode (NOME) | 11 | 0.188 | [-0.086,0.462] | 0.208 |
| Robust adjusted profile score (RAPS) | 11 | 0.135 | [-0.016,0.285] | 0.08 |
| MRPRESSO | 11 | 0.131 | [0.022,0.240] | 0.04* |
| **IP10->Other disorders of optic [2nd] nerve and visual pathways** | | | | |
| MR Egger (bootstrap) | 12 | -0.095 | [-0.516,0.326] | 0.3 |
| Simple median | 12 | -0.235 | [-0.587,0.118] | 0.192 |
| Weighted median | 12 | -0.191 | [-0.388,0.007] | 0.059 |
| Penalised weighted median | 12 | -0.191 | [-0.371,-0.010] | 0.039* |
| IVW radial | 12 | -0.21 | [-0.355,-0.065] | 0.004* |
| Inverse variance weighted (multiplicative random effects) | 12 | -0.207 | [-0.347,-0.068] | 0.004* |
| Inverse variance weighted (fixed effects) | 12 | -0.207 | [-0.359,-0.056] | 0.007* |
| Simple mode | 12 | -0.347 | [-0.797,0.103] | 0.159 |
| Weighted mode | 12 | -0.185 | [-0.362,-0.007] | 0.066 |
| Weighted mode (NOME) | 12 | -0.185 | [-0.365,-0.004] | 0.07 |
| Simple mode (NOME) | 12 | -0.347 | [-0.756,0.062] | 0.125 |
| Robust adjusted profile score (RAPS) | 12 | -0.221 | [-0.395,-0.047] | 0.013* |
| MRPRESSO | 12 | -0.207 | [-0.347,-0.068] | 0.014* |
| **IP10->Corneal degeneration** | | | | |
| MR Egger (bootstrap) | 10 | 0.505 | [-1.774,2.784] | 0.287 |
| Simple median | 10 | 0.506 | [-0.445,1.457] | 0.297 |
| Weighted median | 10 | 0.669 | [-0.228,1.565] | 0.144 |
| Penalised weighted median | 10 | 0.669 | [-0.260,1.597] | 0.158 |
| IVW radial | 10 | 0.597 | [0.025,1.169] | 0.041* |
| Inverse variance weighted (multiplicative random effects) | 10 | 0.599 | [0.030,1.167] | 0.039* |
| Inverse variance weighted (fixed effects) | 10 | 0.599 | [-0.096,1.293] | 0.091 |
| Simple mode | 10 | 0.58 | [-0.642,1.801] | 0.377 |
| Weighted mode | 10 | 0.694 | [-0.223,1.612] | 0.172 |
| Weighted mode (NOME) | 10 | 0.709 | [-0.269,1.686] | 0.189 |
| Simple mode (NOME) | 10 | 0.58 | [-0.670,1.829] | 0.387 |
| Robust adjusted profile score (RAPS) | 10 | 0.589 | [-0.176,1.355] | 0.131 |
| MRPRESSO | 10 | 0.599 | [0.030,1.167] | 0.069 |
| **IP10->Other and unspecified visual disturbances** | | | | |
| MR Egger (bootstrap) | 12 | -0.034 | [-0.701,0.633] | 0.429 |
| Simple median | 12 | 0.362 | [0.006,0.718] | 0.046* |
| Weighted median | 12 | 0.283 | [-0.049,0.615] | 0.095 |
| Penalised weighted median | 12 | 0.283 | [-0.063,0.628] | 0.109 |
| IVW radial | 12 | 0.189 | [0.004,0.373] | 0.045* |
| Inverse variance weighted (multiplicative random effects) | 12 | 0.187 | [0.003,0.372] | 0.047* |
| Inverse variance weighted (fixed effects) | 12 | 0.187 | [-0.056,0.430] | 0.131 |
| Simple mode | 12 | 0.425 | [-0.154,1.004] | 0.178 |
| Weighted mode | 12 | 0.374 | [-0.090,0.839] | 0.142 |
| Weighted mode (NOME) | 12 | 0.374 | [-0.078,0.827] | 0.133 |
| Simple mode (NOME) | 12 | 0.425 | [-0.164,1.014] | 0.185 |
| Robust adjusted profile score (RAPS) | 12 | 0.203 | [-0.065,0.471] | 0.138 |
| MRPRESSO | 12 | 0.187 | [0.003,0.372] | 0.072 |
| **TNFa->Other, unspecified and serous retinal detachments** | | | | |
| MR Egger (bootstrap) | 22 | 0.153 | [-0.661,0.966] | 0.35 |
| Simple median | 22 | 0.142 | [-0.318,0.602] | 0.546 |
| Weighted median | 22 | 0.177 | [-0.286,0.640] | 0.454 |
| Penalised weighted median | 22 | 0.177 | [-0.268,0.621] | 0.436 |
| IVW radial | 22 | 0.296 | [0.019,0.573] | 0.036* |
| Inverse variance weighted (multiplicative random effects) | 22 | 0.296 | [0.020,0.573] | 0.036* |
| Inverse variance weighted (fixed effects) | 22 | 0.296 | [-0.041,0.633] | 0.085 |
| Simple mode | 22 | -0.04 | [-0.873,0.792] | 0.926 |
| Weighted mode | 22 | -0.031 | [-0.849,0.788] | 0.942 |
| Weighted mode (NOME) | 22 | -0.031 | [-0.860,0.799] | 0.943 |
| Simple mode (NOME) | 22 | -0.04 | [-0.891,0.811] | 0.927 |
| Robust adjusted profile score (RAPS) | 22 | 0.306 | [-0.064,0.676] | 0.105 |
| MRPRESSO | 22 | 0.296 | [0.020,0.573] | 0.048* |
| **TNFa->Normotensive glaucoma** | | | | |
| MR Egger (bootstrap) | 19 | 0.222 | [-0.401,0.845] | 0.236 |
| Simple median | 19 | 0.19 | [-0.161,0.540] | 0.29 |
| Weighted median | 19 | 0.249 | [-0.100,0.598] | 0.162 |
| Penalised weighted median | 19 | 0.249 | [-0.102,0.600] | 0.164 |
| IVW radial | 19 | 0.272 | [0.068,0.475] | 0.009* |
| Inverse variance weighted (multiplicative random effects) | 19 | 0.272 | [0.069,0.476] | 0.009* |
| Inverse variance weighted (fixed effects) | 19 | 0.272 | [0.030,0.515] | 0.028* |
| Simple mode | 19 | 0.157 | [-0.499,0.813] | 0.645 |
| Weighted mode | 19 | 0.492 | [-0.113,1.098] | 0.128 |
| Weighted mode (NOME) | 19 | 0.522 | [-0.111,1.155] | 0.123 |
| Simple mode (NOME) | 19 | 0.157 | [-0.516,0.830] | 0.654 |
| Robust adjusted profile score (RAPS) | 19 | 0.287 | [0.020,0.554] | 0.035* |
| MRPRESSO | 19 | 0.272 | [0.069,0.476] | 0.017* |
| **TNFa->Paralytic strabismus** | | | | |
| MR Egger (bootstrap) | 19 | -0.072 | [-0.689,0.545] | 0.394 |
| Simple median | 19 | 0.199 | [-0.101,0.499] | 0.193 |
| Weighted median | 19 | 0.173 | [-0.146,0.493] | 0.287 |
| Penalised weighted median | 19 | 0.173 | [-0.125,0.471] | 0.254 |
| IVW radial | 19 | 0.182 | [0.016,0.348] | 0.032* |
| Inverse variance weighted (multiplicative random effects) | 19 | 0.182 | [0.016,0.348] | 0.032* |
| Inverse variance weighted (fixed effects) | 19 | 0.182 | [-0.047,0.410] | 0.119 |
| Simple mode | 19 | 0.136 | [-0.393,0.665] | 0.62 |
| Weighted mode | 19 | 0.11 | [-0.368,0.588] | 0.657 |
| Weighted mode (NOME) | 19 | 0.12 | [-0.348,0.589] | 0.621 |
| Simple mode (NOME) | 19 | 0.136 | [-0.387,0.659] | 0.617 |
| Robust adjusted profile score (RAPS) | 19 | 0.182 | [-0.071,0.435] | 0.159 |
| MRPRESSO | 19 | 0.182 | [0.016,0.348] | 0.046* |

**Fig S6.2 UVMR results of neonatal cytokine levels (IL10/IL12p70/IL6/IP10/ TNF-α) on adult eye diseases.**

| Methods | nSNPs | OR | CI | PVAL |
|---|---|---|---|---|
| **CTACK->Disorders of vitreous body and globe** | | | | |
| mvmr_ivw | 20 | 0.894 | [0.815,0.981] | 0.018* |
| mvmr_lasso | 20 | 0.894 | [0.815,0.981] | 0.018* |
| mvmr_median | 20 | 0.95 | [0.823,1.098] | 0.485 |
| mvmr_egger | 20 | 0.811 | [0.702,0.938] | 0.005* |
| **GROA->Normotensive glaucoma** | | | | |
| mvmr_ivw | 17 | 0.617 | [0.483,0.789] | 0* |
| mvmr_lasso | 17 | 0.617 | [0.483,0.789] | 0* |
| mvmr_median | 17 | 0.598 | [0.412,0.867] | 0.007* |
| mvmr_egger | 17 | 0.613 | [0.364,1.035] | 0.067 |
| **IL10->Normotensive glaucoma** | | | | |
| mvmr_ivw | 27 | 0.706 | [0.539,0.926] | 0.012* |
| mvmr_lasso | 27 | 0.744 | [0.554,0.998] | 0.049* |
| mvmr_median | 27 | 0.71 | [0.471,1.069] | 0.101 |
| mvmr_egger | 27 | 0.572 | [0.346,0.946] | 0.03* |
| **IL12p70->Disorders of vitreous body** | | | | |
| mvmr_ivw | 23 | 1.15 | [1.013,1.306] | 0.031* |
| mvmr_lasso | 23 | 1.15 | [1.013,1.306] | 0.031* |
| mvmr_median | 23 | 1.194 | [0.991,1.438] | 0.062 |
| mvmr_egger | 23 | 1.324 | [0.943,1.861] | 0.105 |
| **IL12p70->Other and unspecified visual disturbances** | | | | |
| mvmr_ivw | 23 | 1.435 | [1.075,1.916] | 0.014* |
| mvmr_lasso | 23 | 1.435 | [1.075,1.916] | 0.014* |
| mvmr_median | 23 | 1.634 | [1.085,2.461] | 0.019* |
| mvmr_egger | 23 | 2.815 | [1.361,5.822] | 0.005* |
| **IL6->Corneal degeneration** | | | | |
| mvmr_ivw | 24 | 1.754 | [1.067,2.885] | 0.027* |
| mvmr_lasso | 24 | 1.754 | [1.067,2.885] | 0.027* |
| mvmr_median | 24 | 1.898 | [0.958,3.762] | 0.066 |
| mvmr_egger | 24 | 1.813 | [0.859,3.825] | 0.118 |
| **IL6->Normotensive glaucoma** | | | | |
| mvmr_ivw | 20 | 0.847 | [0.718,0.999] | 0.049* |
| mvmr_lasso | 20 | 0.847 | [0.718,0.999] | 0.049* |
| mvmr_median | 20 | 0.984 | [0.76,1.274] | 0.902 |
| mvmr_egger | 20 | 0.78 | [0.598,1.017] | 0.066 |
| **IP10->Disorders of vitreous body** | | | | |
| mvmr_ivw | 24 | 0.969 | [0.942,0.995] | 0.024* |
| mvmr_lasso | 24 | 0.971 | [0.948,0.996] | 0.02* |
| mvmr_median | 24 | 0.965 | [0.934,0.996] | 0.029* |
| mvmr_egger | 24 | 0.969 | [0.939,1.001] | 0.06 |
| **IP10->Intermittent heterotropia** | | | | |
| mvmr_ivw | 33 | 1.17 | [1.076,1.273] | 0* |
| mvmr_lasso | 33 | 1.17 | [1.076,1.273] | 0* |
| mvmr_median | 33 | 1.164 | [1.035,1.309] | 0.011* |
| mvmr_egger | 33 | 1.171 | [1.069,1.283] | 0.001* |

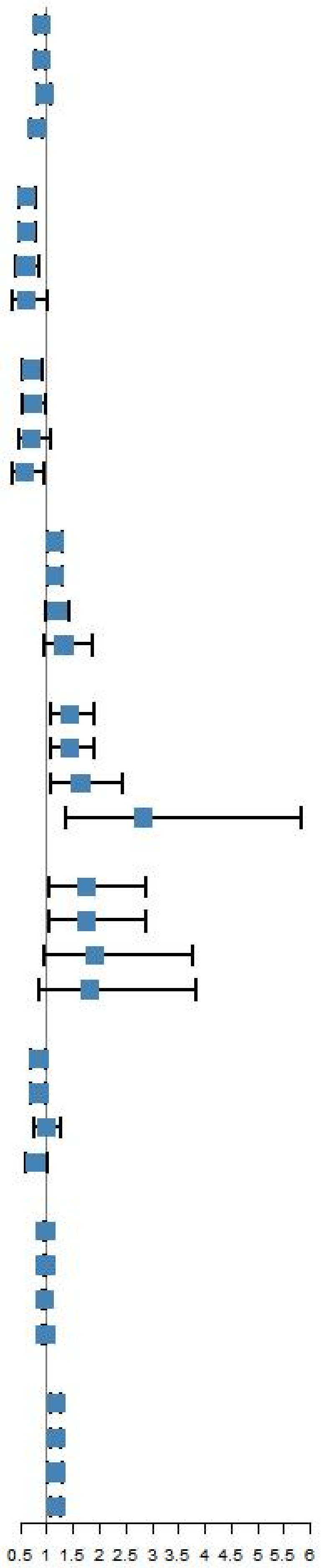

**Fig S7 MVMR results of neonatal cytokine levels on adult eye diseases adjusting for adult cytokine levels.**

**Table S4.1 Pleiotropy test of UVMR of early preterm birth on adult eye diseases**

| **Outcome** | **Intercept** | **se** | ***P*** |
|---|---|---|---|
| Retinal vascular disorders | -0.03 | 0.02 | 0.14 |
| Age-related macular degeneration (whether dry or wet) | 0.01 | 0.01 | 0.66 |
| Papilloedema, unspecified | -0.03 | 0.05 | 0.56 |
| Other and unspecified vascular occlusions | -0.04 | 0.02 | 0.12 |
| Retinal vascular occlusion | -0.03 | 0.02 | 0.18 |
| Other and unspecified iridocyclitis | -0.03 | 0.04 | 0.52 |
| Iridocyclitis | 0.00 | 0.01 | 1.00 |
| Disorders of optic nerve and visual pathways | -0.02 | 0.02 | 0.38 |
| Degeneration of macula and posterior pole | 0.00 | 0.01 | 0.90 |
| Disorders of sclera, cornea, iris and ciliary body | 0.01 | 0.01 | 0.37 |
| Other disorders of eyelid | 0.00 | 0.01 | 0.68 |
| Other disorders of optic [2nd] nerve and visual pathways | -0.03 | 0.03 | 0.26 |
| Acute and subacute iridocyclitis | -0.01 | 0.01 | 0.67 |
| Blepharochalasis | 0.00 | 0.01 | 0.87 |
| Chronic conjunctivitis | -0.02 | 0.04 | 0.68 |

**Table S4.2 Pleiotropy test of UVMR of preterm birth on adult eye diseases**

| **Outcome** | **Intercept** | **se** | ***P*** |
|---|---|---|---|
| Glaucoma, exfoliation | -0.01 | 0.02 | 0.63 |
| Disorders of ocular muscles, binocular movement, accommodation and refraction | 0.00 | 0.01 | 0.52 |
| Disorders of refraction and accommodation | 0.00 | 0.01 | 0.72 |
| Ptosis of eyelid | 0.01 | 0.02 | 0.41 |
| Normotensive glaucoma | -0.01 | 0.02 | 0.66 |
| Other and unspecified visual disturbances | -0.02 | 0.02 | 0.46 |
| Primary open-angle glaucoma | 0.00 | 0.01 | 0.68 |
| Paralytic strabismus | 0.01 | 0.02 | 0.63 |
| Disorders of sclera, cornea, iris and ciliary body | 0.00 | 0.01 | 0.87 |
| Diabetic retinopathy (H7_RETINOPATHYDIAB) | 0.00 | 0.01 | 0.91 |
| Strabismus | 0.01 | 0.01 | 0.34 |

**Table S4.3 Pleiotropy test of UVMR of postterm birth on adult eye diseases**

| Outcome | Intercept | se | *P* |
|---|---|---|---|
| Optic neuritis | -0.02 | 0.02 | 0.42 |
| Atopic conjunctivitis | -0.01 | 0.02 | 0.74 |
| Disorders of lacrimal system and orbit in diseases classified elsewhere | 0.01 | 0.03 | 0.66 |
| Other, unspecified and serous retinal detachments | 0.00 | 0.03 | 0.97 |
| Convergent concomitant strabismus | 0.00 | 0.02 | 0.92 |
| Other specified and unspecified retinal disorders | 0.01 | 0.03 | 0.74 |
| Disorders of orbit | -0.01 | 0.02 | 0.64 |
| Intermittent heterotropia | -0.02 | 0.02 | 0.42 |
| Corneal degeneration | -0.01 | 0.05 | 0.80 |
| Other retinal disorders | 0.00 | 0.01 | 0.95 |

**Table S4.4 Pleiotropy test of UVMR of gest duration on adult eye diseases**

| **Outcome** | **Intercept** | **se** | ***P*** |
|---|---|---|---|
| Disorders of vitreous body | 0.00 | 0.01 | 0.90 |
| Dacryoadenitis | -0.09 | 0.05 | 0.11 |
| Central retinal artery occlusion | 0.01 | 0.03 | 0.73 |
| Visual field defects | 0.01 | 0.02 | 0.35 |
| Ectropion of eyelid | 0.01 | 0.02 | 0.72 |
| Disorders of vitreous body and globe | 0.00 | 0.01 | 0.66 |
| Other and unspecified disorders of vitreous body | 0.00 | 0.01 | 0.79 |

**Table S5.1 Heterogeneity test of UVMR of early preterm birth on adult eye diseases**

| Outcome | Method | df | Q | *P* |
|---|---|---|---|---|
| Retinal vascular disorders | MR Egger | 75 | 63.81 | 0.82 |
| Retinal vascular disorders | Inverse variance weighted | 76 | 66.01 | 0.79 |
| Age-related macular degeneration (whether dry or wet) | MR Egger | 75 | 78.97 | 0.35 |
| Age-related macular degeneration (whether dry or wet) | Inverse variance weighted | 76 | 79.17 | 0.38 |
| Papilloedema, unspecified | MR Egger | 75 | 73.03 | 0.54 |
| Papilloedema, unspecified | Inverse variance weighted | 76 | 73.37 | 0.56 |
| Other and unspecified vascular occlusions | MR Egger | 75 | 72.97 | 0.54 |
| Other and unspecified vascular occlusions | Inverse variance weighted | 76 | 75.40 | 0.50 |
| Retinal vascular occlusion | MR Egger | 75 | 62.99 | 0.84 |
| Retinal vascular occlusion | Inverse variance weighted | 76 | 64.82 | 0.82 |
| Other and unspecified iridocyclitis | MR Egger | 75 | 50.54 | 0.99 |
| Other and unspecified iridocyclitis | Inverse variance weighted | 76 | 50.97 | 0.99 |
| Iridocyclitis | MR Egger | 75 | 56.80 | 0.94 |
| Iridocyclitis | Inverse variance weighted | 76 | 56.80 | 0.95 |
| Disorders of optic nerve and visual pathways | MR Egger | 75 | 77.42 | 0.40 |
| Disorders of optic nerve and visual pathways | Inverse variance weighted | 76 | 78.23 | 0.41 |
| Degeneration of macula and posterior pole | MR Egger | 75 | 72.86 | 0.55 |
| Degeneration of macula and posterior pole | Inverse variance weighted | 76 | 72.88 | 0.58 |
| Disorders of sclera, cornea, iris and ciliary body | MR Egger | 75 | 56.63 | 0.94 |
| Disorders of sclera, cornea, iris and ciliary body | Inverse variance weighted | 76 | 57.46 | 0.94 |
| Other disorders of eyelid | MR Egger | 75 | 61.34 | 0.87 |
| Other disorders of eyelid | Inverse variance weighted | 76 | 61.51 | 0.89 |
| Other disorders of optic [2nd] nerve and visual pathways | MR Egger | 75 | 69.98 | 0.64 |
| Other disorders of optic [2nd] nerve and visual pathways | Inverse variance weighted | 76 | 71.25 | 0.63 |
| Acute and subacute iridocyclitis | MR Egger | 75 | 69.80 | 0.65 |
| Acute and subacute iridocyclitis | Inverse variance weighted | 76 | 69.98 | 0.67 |
| Blepharochalasis | MR Egger | 75 | 58.08 | 0.93 |
| Blepharochalasis | Inverse variance weighted | 76 | 58.11 | 0.94 |
| Chronic conjunctivitis | MR Egger | 75 | 63.36 | 0.83 |
| Chronic conjunctivitis | Inverse variance weighted | 76 | 63.54 | 0.85 |

**Table S5.2 Heterogeneity test of UVMR of preterm birth on adult eye diseases**

| Outcome | Method | df | Q | *P* |
|---|---|---|---|---|
| Glaucoma, exfoliation | MR Egger | 71 | 81.22 | 0.19 |
| Glaucoma, exfoliation | Inverse variance weighted | 72 | 81.48 | 0.21 |
| Disorders of ocular muscles, binocular movement, accommodation and refraction | MR Egger | 71 | 78.43 | 0.26 |
| Disorders of ocular muscles, binocular movement, accommodation and refraction | Inverse variance weighted | 72 | 78.89 | 0.27 |
| Disorders of refraction and accommodation | MR Egger | 71 | 78.77 | 0.25 |
| Disorders of refraction and accommodation | Inverse variance weighted | 72 | 78.92 | 0.27 |
| Ptosis of eyelid | MR Egger | 71 | 81.60 | 0.18 |
| Ptosis of eyelid | Inverse variance weighted | 72 | 82.37 | 0.19 |
| Normotensive glaucoma | MR Egger | 71 | 64.48 | 0.69 |
| Normotensive glaucoma | Inverse variance weighted | 72 | 64.67 | 0.72 |
| Other and unspecified visual disturbances | MR Egger | 71 | 73.50 | 0.40 |
| Other and unspecified visual disturbances | Inverse variance weighted | 72 | 74.07 | 0.41 |
| Primary open-angle glaucoma | MR Egger | 71 | 81.61 | 0.18 |
| Primary open-angle glaucoma | Inverse variance weighted | 72 | 81.80 | 0.20 |
| Paralytic strabismus | MR Egger | 71 | 99.69 | 0.01 |
| Paralytic strabismus | Inverse variance weighted | 72 | 100.01 | 0.02 |
| Disorders of sclera, cornea, iris and ciliary body | MR Egger | 71 | 53.48 | 0.94 |
| Disorders of sclera, cornea, iris and ciliary body | Inverse variance weighted | 72 | 53.51 | 0.95 |
| Diabetic retinopathy (H7_RETINOPATHYDIAB) | MR Egger | 71 | 61.75 | 0.78 |
| Diabetic retinopathy (H7_RETINOPATHYDIAB) | Inverse variance weighted | 72 | 61.76 | 0.80 |
| Strabismus | MR Egger | 71 | 73.89 | 0.38 |
| Strabismus | Inverse variance weighted | 72 | 74.83 | 0.39 |

**Table S5.3 Heterogeneity test of UVMR of postterm birth on adult eye diseases**

| Outcome | Method | df | Q | *P* |
|---|---|---|---|---|
| Optic neuritis | MR Egger | 66 | 60.44 | 0.67 |
| Optic neuritis | Inverse variance weighted | 67 | 61.10 | 0.68 |
| Atopic conjunctivitis | MR Egger | 66 | 66.76 | 0.45 |
| Atopic conjunctivitis | Inverse variance weighted | 67 | 66.87 | 0.48 |
| Disorders of lacrimal system and orbit in diseases classified elsewhere | MR Egger | 66 | 64.99 | 0.51 |
| Disorders of lacrimal system and orbit in diseases classified elsewhere | Inverse variance weighted | 67 | 65.18 | 0.54 |
| Other, unspecified and serous retinal detachments | MR Egger | 66 | 48.77 | 0.94 |
| Other, unspecified and serous retinal detachments | Inverse variance weighted | 67 | 48.77 | 0.95 |
| Convergent concomitant strabismus | MR Egger | 66 | 44.16 | 0.98 |
| Convergent concomitant strabismus | Inverse variance weighted | 67 | 44.17 | 0.99 |
| Other specified and unspecified retinal disorders | MR Egger | 66 | 51.95 | 0.90 |
| Other specified and unspecified retinal disorders | Inverse variance weighted | 67 | 52.06 | 0.91 |
| Disorders of orbit | MR Egger | 66 | 66.67 | 0.45 |
| Disorders of orbit | Inverse variance weighted | 67 | 66.90 | 0.48 |
| Intermittent heterotropia | MR Egger | 66 | 63.37 | 0.57 |

| | | | | |
|---|---|---|---|---|
| Intermittent heterotropia | Inverse variance weighted | 67 | 64.02 | 0.58 |
| Corneal degeneration | MR Egger | 66 | 51.84 | 0.90 |
| Corneal degeneration | Inverse variance weighted | 67 | 51.91 | 0.91 |
| Other retinal disorders | MR Egger | 66 | 73.98 | 0.23 |
| Other retinal disorders | Inverse variance weighted | 67 | 73.99 | 0.26 |

**Table S5.4 Heterogeneity test of UVMR of gest duration on adult eye diseases**

| Outcome | Method | df | Q | *P* |
|---|---|---|---|---|
| Disorders of vitreous body | MR Egger | 101 | 89.68 | 0.78 |
| Disorders of vitreous body | Inverse variance weighted | 102 | 89.70 | 0.80 |
| Dacryoadenitis | MR Egger | 101 | 110.66 | 0.24 |
| Dacryoadenitis | Inverse variance weighted | 102 | 113.57 | 0.20 |
| Central retinal artery occlusion | MR Egger | 101 | 92.55 | 0.71 |
| Central retinal artery occlusion | Inverse variance weighted | 102 | 92.68 | 0.73 |
| Visual field defects | MR Egger | 101 | 105.92 | 0.35 |
| Visual field defects | Inverse variance weighted | 102 | 106.84 | 0.35 |
| Ectropion of eyelid | MR Egger | 101 | 95.24 | 0.64 |
| Ectropion of eyelid | Inverse variance weighted | 102 | 95.37 | 0.67 |
| Disorders of vitreous body and globe | MR Egger | 101 | 93.05 | 0.70 |
| Disorders of vitreous body and globe | Inverse variance weighted | 102 | 93.25 | 0.72 |
| Other and unspecified disorders of vitreous body | MR Egger | 101 | 88.12 | 0.82 |
| Other and unspecified disorders of vitreous body | Inverse variance weighted | 102 | 88.19 | 0.83 |

**Table S6 Pleiotropy test of UVMR results of maternal pregnancy disorders on neonatal cytokine levels.**

| Exposure | Outcome | Intercept | se | *P* |
|---|---|---|---|---|
| Early preterm birth | IL10 | -0.02 | 0.02 | 0.49 |
| Early preterm birth | TNF-α | -0.01 | 0.02 | 0.61 |
| Preterm birth | CTACK | -0.03 | 0.03 | 0.29 |
| Preterm birth | GROA | -0.03 | 0.03 | 0.33 |

**Table S7 Heterogeneity test of UVMR of maternal pregnancy disorders on neonatal cytokine levels**

| Exposure | Outcome | Method | df | Q | *P* |
|---|---|---|---|---|---|
| Early preterm birth | IL10 | MR Egger | 10 | 4.09 | 0.94 |
| Early preterm birth | IL10 | Inverse variance weighted | 11 | 4.60 | 0.95 |
| Early preterm birth | TNF-α | MR Egger | 10 | 4.09 | 0.94 |
| Early preterm birth | TNF-α | Inverse variance weighted | 11 | 4.36 | 0.96 |
| Preterm birth | CTACK | MR Egger | 10 | 6.95 | 0.73 |
| Preterm birth | CTACK | Inverse variance weighted | 11 | 8.20 | 0.69 |
| Preterm birth | GROA | MR Egger | 10 | 12.39 | 0.26 |
| Preterm birth | GROA | Inverse variance weighted | 11 | 13.69 | 0.25 |

**Table S8 Pleiotropy test of UVMR of maternal pregnancy disorders on adult cytokine levels**

| Exposure | Outcome | Intercept | se | *P* |
|---|---|---|---|---|
| Early preterm birth | IP10 | 0.00 | 0.02 | 0.98 |
| Preterm birth | IL12p70 | -0.01 | 0.01 | 0.55 |

**Table S9 Heterogeneity test of UVMR of maternal pregnancy disorders on adult cytokine levels**

| Exposure | Outcome | Method | df | Q | *P* |
|---|---|---|---|---|---|
| Early preterm birth | IP10 | MR Egger | 67 | 53.43 | 0.89 |
| Early preterm birth | IP10 | Inverse variance weighted | 68 | 53.43 | 0.90 |
| Preterm birth | IL12p70 | MR Egger | 66 | 68.69 | 0.39 |
| Preterm birth | IL12p70 | Inverse variance weighted | 67 | 69.06 | 0.41 |

**Table S10 Pleiotropy test of MVMR of maternal pregnancy disorders on adult cytokine levels adjusting for neonatal levels**

| Exposure | Outcome | Q | *P* |
|---|---|---|---|
| Preterm birth | IL6 | 20.34 | 0.88 |

**Table S11 Pleiotropy test of UVMR of adult cytokine levels on adult eye diseases**

| Exposure | Outcome | Intercept | se | *P* |
|---|---|---|---|---|
| CTACK | Other and unspecified visual disturbances | -0.03 | 0.02 | 0.08 |
| CTACK | Paralytic strabismus | 0.01 | 0.02 | 0.47 |
| GROA | Dacryoadenitis | 0.03 | 0.07 | 0.64 |
| GROA | Intermittent heterotropia | 0.01 | 0.02 | 0.73 |
| IL10 | Disorders of optic nerve and visual pathways | -0.01 | 0.02 | 0.62 |
| IL10 | Disorders of vitreous body | 0.00 | 0.01 | 0.87 |
| IL10 | Disorders of vitreous body and globe | 0.00 | 0.01 | 0.77 |
| IL10 | Other and unspecified disorders of vitreous body | 0.00 | 0.01 | 0.68 |
| IL10 | Normotensive glaucoma | 0.03 | 0.03 | 0.28 |
| IL12p70 | Corneal degeneration | 0.05 | 0.06 | 0.38 |
| IL6 | Disorders of orbit | -0.02 | 0.02 | 0.34 |
| IL6 | Intermittent heterotropia | 0.00 | 0.02 | 0.91 |
| TNF-α | Convergent concomitant strabismus | -0.01 | 0.02 | 0.63 |
| TNF-α | Intermittent heterotropia | -0.02 | 0.03 | 0.53 |

**Table S12 Heterogeneity test of UVMR of adult cytokine levels on adult eye diseases**

| Exposure | Outcome | Method | df | Q | *P* |
|---|---|---|---|---|---|
| CTACK | Other and unspecified visual disturbances | MR Egger | 63 | 42.18 | 0.98 |
| CTACK | Other and unspecified visual disturbances | Inverse variance weighted | 64 | 45.31 | 0.96 |
| CTACK | Paralytic strabismus | MR Egger | 63 | 64.76 | 0.42 |
| CTACK | Paralytic strabismus | Inverse variance weighted | 64 | 65.30 | 0.43 |
| GROA | Dacryoadenitis | MR Egger | 51 | 52.79 | 0.40 |
| GROA | Dacryoadenitis | Inverse variance weighted | 52 | 53.02 | 0.43 |
| GROA | Intermittent heterotropia | MR Egger | 51 | 45.17 | 0.70 |
| GROA | Intermittent heterotropia | Inverse variance weighted | 52 | 45.29 | 0.73 |
| IL10 | Disorders of optic nerve and visual pathways | MR Egger | 49 | 68.78 | 0.03 |
| IL10 | Disorders of optic nerve and visual pathways | Inverse variance weighted | 50 | 69.14 | 0.04 |
| IL10 | Disorders of vitreous body | MR Egger | 49 | 66.51 | 0.05 |
| IL10 | Disorders of vitreous body | Inverse variance weighted | 50 | 66.55 | 0.06 |
| IL10 | Disorders of vitreous body and globe | MR Egger | 49 | 67.11 | 0.04 |
| IL10 | Disorders of vitreous body and globe | Inverse variance weighted | 50 | 67.23 | 0.05 |
| IL10 | Other and unspecified disorders of vitreous body | MR Egger | 49 | 49.14 | 0.47 |
| IL10 | Other and unspecified disorders of vitreous body | Inverse variance weighted | 50 | 49.31 | 0.50 |
| IL10 | Normotensive glaucoma | MR Egger | 49 | 74.84 | 0.01 |
| IL10 | Normotensive glaucoma | Inverse variance weighted | 50 | 76.65 | 0.01 |
| IL12p70 | Corneal degeneration | MR Egger | 50 | 43.30 | 0.74 |
| IL12p70 | Corneal degeneration | Inverse variance weighted | 51 | 44.08 | 0.74 |
| IL6 | Disorders of orbit | MR Egger | 50 | 44.56 | 0.69 |
| IL6 | Disorders of orbit | Inverse variance weighted | 51 | 45.47 | 0.69 |
| IL6 | Intermittent heterotropia | MR Egger | 50 | 58.84 | 0.18 |
| IL6 | Intermittent heterotropia | Inverse variance weighted | 51 | 58.85 | 0.21 |
| TNFa | Convergent concomitant strabismus | MR Egger | 47 | 42.90 | 0.64 |
| TNFa | Convergent concomitant strabismus | Inverse variance weighted | 48 | 43.14 | 0.67 |
| TNFa | Intermittent heterotropia | MR Egger | 47 | 47.90 | 0.44 |
| TNFa | Intermittent heterotropia | Inverse variance weighted | 48 | 48.31 | 0.46 |

**Table S13 Pleiotropy test of UVMR of neonatal cytokine levels on adult eye diseases**

| Exposure | Outcome | Intercept | se | *P* |
|---|---|---|---|---|
| CTACK | Age-related macular degeneration (whether dry or wet) | -0.02 | 0.02 | 0.43 |
| CTACK | Papilloedema, unspecified | 0.11 | 0.10 | 0.29 |
| CTACK | Ectropion of eyelid | 0.00 | 0.06 | 0.97 |
| CTACK | Other and unspecified disorders of vitreous body | 0.00 | 0.02 | 0.83 |
| CTACK | Optic neuritis | 0.02 | 0.06 | 0.76 |
| CTACK | Diabetic retinopathy (H7_RETINOPATHYDIAB) | 0.01 | 0.02 | 0.64 |
| GROA | Other and unspecified iridocyclitis | -0.01 | 0.11 | 0.96 |
| GROA | Central retinal artery occlusion | -0.14 | 0.12 | 0.29 |
| GROA | Visual field defects | -0.02 | 0.05 | 0.71 |
| GROA | Disorders of vitreous body and globe | -0.01 | 0.02 | 0.52 |
| IL10 | Central retinal artery occlusion | -0.06 | 0.10 | 0.51 |
| IL10 | Other specified and unspecified retinal disorders | 0.04 | 0.08 | 0.64 |
| IL12p70 | Retinal vascular occlusion | 0.04 | 0.06 | 0.47 |
| IL12p70 | Degeneration of macula and posterior pole | -0.03 | 0.03 | 0.32 |
| IL12p70 | Acute and subacute iridocyclitis | 0.00 | 0.05 | 0.98 |
| IL6 | Strabismus | 0.02 | 0.03 | 0.41 |
| IP10 | Other disorders of optic [2nd] nerve and visual pathways | 0.03 | 0.03 | 0.24 |
| IP10 | Corneal degeneration | -0.03 | 0.10 | 0.80 |
| IP10 | Other and unspecified visual disturbances | 0.03 | 0.04 | 0.47 |
| TNF-α | Other, unspecified and serous retinal detachments | -0.03 | 0.06 | 0.61 |
| TNF-α | Normotensive glaucoma | -0.02 | 0.04 | 0.56 |
| TNF-α | Paralytic strabismus | -0.02 | 0.04 | 0.70 |

**Table S14.1 Heterogeneity test of UVMR of neonatal cytokine levels on adult eye diseases by Inverse variance weighted**

| Exposure | Outcome | df | Q | *P* |
|---|---|---|---|---|
| CTACK | Age-related macular degeneration (whether dry or wet) | 15 | 6.21 | 0.98 |
| CTACK | Papilloedema, unspecified | 15 | 5.19 | 0.99 |
| CTACK | Ectropion of eyelid | 15 | 12.19 | 0.66 |
| CTACK | Other and unspecified disorders of vitreous body | 15 | 9.15 | 0.87 |
| CTACK | Optic neuritis | 15 | 5.09 | 0.99 |
| CTACK | Diabetic retinopathy (H7_RETINOPATHYDIAB) | 15 | 6.51 | 0.97 |
| GROA | Other and unspecified iridocyclitis | 6 | 1.44 | 0.96 |
| GROA | Central retinal artery occlusion | 8 | 1.75 | 0.99 |
| GROA | Visual field defects | 10 | 5.98 | 0.82 |
| GROA | Disorders of vitreous body and globe | 10 | 10.50 | 0.40 |
| IL10 | Central retinal artery occlusion | 16 | 6.21 | 0.99 |
| IL10 | Other specified and unspecified retinal disorders | 17 | 10.05 | 0.90 |
| IL12p70 | Retinal vascular occlusion | 15 | 16.78 | 0.33 |
| IL12p70 | Degeneration of macula and posterior pole | 15 | 8.49 | 0.90 |
| IL12p70 | Acute and subacute iridocyclitis | 13 | 8.48 | 0.81 |
| IL6 | Strabismus | 10 | 6.39 | 0.78 |
| IP10 | Other disorders of optic [2nd] nerve and visual pathways | 11 | 9.26 | 0.60 |
| IP10 | Corneal degeneration | 9 | 6.02 | 0.74 |
| IP10 | Other and unspecified visual disturbances | 11 | 6.34 | 0.85 |
| TNF-α | Other, unspecified and serous retinal detachments | 21 | 14.18 | 0.86 |
| TNF-α | Normotensive glaucoma | 18 | 12.68 | 0.81 |
| TNF-α | Paralytic strabismus | 18 | 9.49 | 0.95 |

**Table S14.2 Heterogeneity test of UVMR of neonatal cytokine levels on adult eye diseases by MR Egger regression**

| Exposure | Outcome | df | Q | *P* |
|---|---|---|---|---|
| CTACK | Age-related macular degeneration (whether dry or wet) | 14 | 5.54 | 0.98 |
| CTACK | Papilloedema, unspecified | 14 | 3.96 | 1.00 |
| CTACK | Ectropion of eyelid | 14 | 12.19 | 0.59 |
| CTACK | Other and unspecified disorders of vitreous body | 14 | 9.10 | 0.82 |
| CTACK | Optic neuritis | 14 | 5.00 | 0.99 |
| CTACK | Diabetic retinopathy (H7_RETINOPATHYDIAB) | 14 | 6.28 | 0.96 |
| GROA | Other and unspecified iridocyclitis | 5 | 1.44 | 0.92 |
| GROA | Central retinal artery occlusion | 7 | 0.46 | 1.00 |
| GROA | Visual field defects | 9 | 5.83 | 0.76 |
| GROA | Disorders of vitreous body and globe | 9 | 10.00 | 0.35 |
| IL10 | Central retinal artery occlusion | 15 | 5.76 | 0.98 |
| IL10 | Other specified and unspecified retinal disorders | 16 | 9.81 | 0.88 |
| IL12p70 | Retinal vascular occlusion | 14 | 16.16 | 0.30 |
| IL12p70 | Degeneration of macula and posterior pole | 14 | 7.44 | 0.92 |
| IL12p70 | Acute and subacute iridocyclitis | 12 | 8.48 | 0.75 |
| IL6 | Strabismus | 9 | 5.65 | 0.77 |
| IP10 | Other disorders of optic [2nd] nerve and visual pathways | 10 | 7.69 | 0.66 |
| IP10 | Corneal degeneration | 8 | 5.96 | 0.65 |
| IP10 | Other and unspecified visual disturbances | 10 | 5.78 | 0.83 |
| TNF-α | Other, unspecified and serous retinal detachments | 20 | 13.91 | 0.83 |
| TNF-α | Normotensive glaucoma | 17 | 12.33 | 0.78 |
| TNF-α | Paralytic strabismus | 17 | 9.34 | 0.93 |

**Table S15 Pleiotropy test of MVMR of neonatal cytokine levels on adult eye diseases adjusting for adult cytokine levels.**

| Exposure | Outcome | Q | *P* |
|---|---|---|---|
| CTACK | Disorders of vitreous body and globe | 16.83 | 0.47 |
| GROA | Normotensive glaucoma | 7.67 | 0.91 |
| IL10 | Normotensive glaucoma | 26.36 | 0.34 |
| IL12p70 | Disorders of vitreous body | 18.72 | 0.54 |
| IL12p70 | Other and unspecified visual disturbances | 15.16 | 0.77 |
| IL6 | Corneal degeneration | 12.61 | 0.92 |
| IL6 | Normotensive glaucoma | 21.54 | 0.20 |
| IP10 | Disorders of vitreous body | 27.19 | 0.16 |
| IP10 | Intermittent heterotropia | 29.25 | 0.40 |

**Table S16 Pleiotropy test of MVMR of maternal pregnancy disorders on adult eye diseases adjusting for neonatal/adult cytokine levels.**

| Exposure | Outcome | Q | *P* |
|---|---|---|---|
| Early preterm birth | Paralytic strabismus | 41.98 | 0.02 |
| Early preterm birth | Disorder of vitreous body | 49.83 | 0.06 |
| Early preterm birth | Disorder of optic nerve and visual pathways | 55.52 | 0.05 |
| Early preterm birth | Normotensive glaucoma | 45.76 | 0.01 |
| Early preterm birth | Disorder of vitreous body and globe | 51.24 | 0.05 |
| Preterm birth | Disorder of vitreous body | 32.42 | 0.12 |
| Preterm birth | Disorder of optic nerve and visual pathways | 38.10 | 0.02 |
| Preterm birth | Normotensive glaucoma | 16.62 | 0.55 |
| Preterm birth | Corneal degeneration | 34.80 | 0.04 |
| Preterm birth | Disorder of vitreous body and globe | 35.29 | 0.36 |
| Preterm birth | Intermittent heterotropia | 31.40 | 0.05 |
| Preterm birth | Disorder of orbit | 31.78 | 0.06 |
